# Acute biomarkers of consciousness are associated with recovery after severe traumatic brain injury

**DOI:** 10.1101/2025.03.02.25322248

**Authors:** Yelena G. Bodien, Matteo Fecchio, Natalie Gilmore, Holly J. Freeman, William R. Sanders, Anogue Meydan, Phoebe K. Lawrence, Alexander S. Atalay, John Kirsch, Brian C. Healy, Brian L. Edlow

## Abstract

**Objective:** Determine whether acute behavioral, electroencephalography (EEG), and functional MRI (fMRI) biomarkers of consciousness are associated with outcome after severe traumatic brain injury (TBI).

**Methods:** Patients with acute severe TBI admitted consecutively to the intensive care unit (ICU) participated in a multimodal battery assessing behavioral level of consciousness (Coma Recovery Scale-Revised [CRS-R]), cognitive motor dissociation (CMD; task-based EEG and fMRI), covert cortical processing (CCP; stimulus-based EEG and fMRI), and default mode network connectivity (DMN; resting-state fMRI). The primary outcome was 6-month Disability Rating Scale (DRS) total scores.

**Results:** We enrolled 55 patients with acute severe TBI. Six-month outcome was available in 45 (45.2±20.7 years old, 70% male), of whom 10 died, all due to withdrawal of life-sustaining treatment (WLST). Behavioral level of consciousness and presence of command-following in the ICU were each associated with lower (i.e., better) DRS scores (p=0.003, p=0.011). EEG and fMRI biomarkers did not strengthen this relationship, but higher DMN connectivity was associated with better recovery on multiple secondary outcome measures. In a subsample of participants without command-following on the CRS-R, CMD (EEG:18%; fMRI:33%) and CCP (EEG:91%; fMRI:79%) were not associated with outcome, an unexpected result that may reflect the high rate of WLST. However, higher DMN connectivity was associated with lower DRS scores (ρ[95%CI]=-0.41[-0.707, -0.027]; p=0.046) in this group.

**Interpretation:** Standardized behavioral assessment in the ICU may improve prediction of recovery from severe TBI. Further research is required to determine whether integrating behavioral, EEG, and fMRI biomarkers of consciousness is more predictive than behavioral assessment alone.

## Introduction

Recovery from severe traumatic brain injury (TBI) occurs along multiple different trajectories. In the days following injury, clinicians have the difficult task of establishing a prognosis, which can range from death to return of all pre-injury functions. This predicted outcome is used to guide interventions, make recommendations about continuation or withdrawal of life-sustaining treatment (WLST), and determine whether rehabilitation services will be offered. Available prognostic models built on large clinical datasets are imprecise at the individual patient level because they rely on a narrow set of predictors (e.g., a single behavioral assessment, visual inspection of imaging data) and crude outcome categories (e.g., severe disability).^1,2^

Reemergence of consciousness is a critical milestone of TBI recovery,^3–6^ as it suggests reintegration of subcortical and cortical brain networks^7^ and is a necessary precursor for return of communication and independence. Yet, the typical clinical assessment of consciousness consists of a coarse bedside behavioral examination, which may be confounded by a patient’s fluctuating level of arousal, central neurological deficits (e.g., aphasia, weakness), peripheral nerve injury, pain, or an examiner’s subjective interpretation of ambiguous responses.^8^ The clinical bedside behavioral assessment results in an approximately 40% rate of misclassifying conscious patients as unconscious.^9–11^

Standardized behavior assessment with a tool such as the Coma Recovery Scale-Revised (CRS-R) improves diagnostic precision.^11^ However, up to 25% of patients with acute^12–14^ and chronic^15–19^ disorders of consciousness (DoC) who do not follow commands on behavioral assessment show signs of covert command-following, referred to as cognitive motor dissociation (CMD),^20^ when assessed with task-based electroencephalography (EEG) or functional magnetic resonance imaging (fMRI). Patients with DoC can also demonstrate covert cortical processing (CCP),^21,22^ indicated by passive association cortex responses to auditory language stimuli, despite the absence of behavioral evidence of language comprehension and expression.^23,24^ An intact default mode network (DMN)^25,26^ identified by resting-state fMRI (rs-fMRI) has also been reported in unresponsive patients. CMD,^13,14^ CCP,^24,27^ and an intact DMN^28–31^ have been associated with better outcome after severe TBI. However, most studies are unimodal,^14^ evaluate outcomes in the acute/subacute phase (e.g., hospital discharge),^29^ or do not use a standardized behavioral assessment to determine level of consciousness.^24,27,30^ Furthermore, functional outcome measures used in prior studies (e.g., dichotomized determination of favorable versus unfavorable outcome on the Glasgow Outcome Scale-Extended [GOSE]) lack granularity and are not designed to assess the full spectrum of TBI recovery.

In this prospective observational study of patients with acute severe TBI in the ICU, we identified behavioral, CMD, CCP, and DMN biomarkers that were associated with 6-month functional outcome on the Disability Rating Scale (DRS).^32^ We implemented recently proposed DoC clinical guidelines that recommend supplementing behavioral assessment with task-based EEG and fMRI^33,34^ and using the highest level of function evident on a multimodal assessment battery to establish diagnosis.^34^ Given that no single measure is ideally designed to capture outcomes in persons with DoC, we selected multiple secondary outcome measures identified recently by the Neurocritical Care Society Curing Coma Campaign as DoC Common Data Elements.^35^

## Materials and methods

### Experimental design

We prospectively screened all patients with TBI admitted to the Neurosciences ICU, Multidisciplinary ICU, and Surgical ICU at Massachusetts General Hospital (MGH) between September 2018 and December 2022. Enrollment inclusion criteria were: 1) age ≥18; 2) head trauma with Glasgow Coma Scale (GCS)^36^ score of 3-8 within 24 hours, unconfounded by sedation, paralysis, hypoxia, hypotension, hypothermia, or concurrent medical illness; and 3) clinical evidence of a DoC defined as coma, vegetative state/unresponsive wakefulness syndrome (VS/UWS),^37,38^ or minimally conscious state (MCS)^6,39^ at time of screening. Exclusion criteria were: 1) history of prior brain injury or neurological disease; and 2) non-English speaking.

Surrogate decision-makers were approached for consent ≥24 hours after injury, and informed consent was obtained in accordance with a research protocol approved by the Mass General Brigham Institutional Review Board. When approved by the clinical team, EEG was performed immediately after informed consent was obtained. fMRI was performed as soon as the patient was clinically stable for transport to the MRI scanner, as determined by the treating ICU clinicians. Whenever possible, the EEG and fMRI were scheduled within 24 hours of one another. Administration of sedative, anxiolytic, and/or analgesic medications was permitted for patient safety or comfort during the EEG and/or fMRI at the clinicians’ discretion.

A cohort of healthy participants was enrolled from the community as a comparison sample. Healthy participants had no history of neurological, psychiatric, cardiovascular, pulmonary, renal or endocrinologic disease. They provided informed consent and completed the same EEG and fMRI protocols as the patients. All patient and healthy participant assessments were performed using standard clinical EEG equipment and a clinical MRI scanner located in the MGH Neurosciences ICU.

### Neurobehavioral and outcome assessments

We acquired demographic and clinical data at the time of enrollment in accordance with the National Institute of Neurologic Disorders and Stroke (NINDS) Common Data Element guidelines for TBI. Immediately prior to EEG and fMRI, each patient participant was assessed with the Coma Recovery Scale-Revised (CRS-R),^40^ a standardized behavioral measure that is recommended by multiple professional organizations for assessment of DoC.^33,34^ The examiner assigned each CRS-R rating a Test Completion Code (Supplementary Table 1) to document whether the assessment was valid, and if not, the rationale for this determination.

We designated the DRS,^32^ assessed 6-months post-TBI, as the primary outcome measure because it was designed to evaluate a wide range of TBI outcomes, from coma to return to employment, and has been used in DoC clinical trials^41^ and observational studies.^42^ The DRS is also an NINDS^43^ and a Neurocritical Care Society Curing Coma Campaign DoC^35^ Common Data Element. However, the total score of the DRS lacks clinical significance and, unlike the GOSE,^44,45^ the DRS has not been endorsed by the US Food and Drug Administration (FDA) for TBI trials. For these reasons, we measured several secondary outcomes: 1) a dichotomous score based on the DRS (DRS_Depend_)^45^ that is a more sensitive measure of dependency than the DRS total score or the GOSE; 2) the GOSE, as an ordinal total score, accounting for TBI and any additional concurrent injuries (GOSE-All); 3) GOSE-All as a dichotomous variable (GOSE ≤3 [lower severe disability or worse] versus GOSE ≥4 [upper severe disability or better]); 4) the GOSE, as an ordinal total score, accounting for only the effects of the TBI rather than the TBI and concurrent injuries (GOSE-TBI)^46^; and 5) GOSE-TBI as a dichotomous variable. Each measure was evaluated in the full sample, a subsample of patients with a CRS-R diagnosis of coma, VS/UWS, or MCS-at the time of EEG or fMRI assessment, and a subsample of patients who survived to 6-months.

Outcome assessment was conducted either in-person or via telephone by trained study staff who were unaware of the results of prior evaluations (i.e., outcome examiner was not aware of acute CRS-R, EEG, or fMRI results). Details regarding the CRS-R, CAP, DRS, and GOSE are provided in the Supplementary Materials.

### EEG data acquisition

During the EEG assessment, participants were instructed to imagine opening and closing their right hand (i.e., active-motor-imagery paradigm) and listen to an audio recording of the Alice in Wonderland book (i.e., passive-language paradigm, see Supplementary Table 2 for task instructions). We acquired EEG data with a 19-electrode clinical XLTEK EEG system (Natus Medical Inc.; Pleasanton, CA) and analyzed data using EEGlab^47^ and customized MATLAB (vR2016b) code (see Supplementary Materials)

### EEG biomarker classification using power spectral density

EEG preprocessing steps are described in the Supplementary Materials. We used a support vector machine with a linear kernel to classify the absolute power estimated at each electrode and averaged within four frequency bands, as previously described.^12^ The output of this analysis is the accuracy of the classifier and a p-value that represents the probability that the classifier differentiated the task/stimulus and rest conditions by chance. This method was described in prior publications.^48^ We considered a patient to have a positive EEG response if p<0.05. EEG data were analyzed by trained study staff who had no knowledge of participants’ behavioral diagnosis. EEG biomarkers were the presence and accuracy of an EEG response to the active-motor-imagery and passive-language tasks.

### fMRI data acquisition

The fMRI assessment began with 10 mins of rs-fMRI (see Supplementary Table 2). Next, we administered two runs of the active-motor-imagery paradigm^12^, and two runs of the passive-language paradigm. MRI data were acquired with a 32-channel head coil on a 3 Tesla Skyra MRI scanner (Siemens Healthineers; Erlangen, Germany) located in the MGH Neurosciences ICU. The parameters of the BOLD fMRI sequence were: echo time (TE)=30 ms, repetition time (TR)=1250 ms, simultaneous multislice (SMS) factor=4, spatial resolution = 2 mm isotropic. The rs-fMRI sequence was the same as that used for task/stimulus-based fMRI. 3D T1-weighted multi-echo magnetization prepared gradient echo (MEMPRAGE) anatomical images were acquired at 1mm isotropic resolution for registration purposes.^49^ See Supplementary Materials for details about fMRI data acquisition.

### Active-motor-imagery, passive-language, and rs-fMRI biomarker analyses

The active-motor-imagery and passive-language fMRI analysis pipeline was described in a prior publication^12^ and is summarized in the Supplementary Materials Text and Supplementary Table 3. We also estimated rs-fMRI seed-based connectivity maps and ROI-to-ROI connectivity matrices to characterize patterns of functional connectivity within the DMN. Functional connectivity strength was represented by Fisher-transformed bivariate correlation coefficients from a weighted general linear model (weighted-GLM).^50,51^ Participants for whom the DMN z-statistic was within the 95% confidence interval of the healthy control participants’ average DMN z-statistic were considered to have DMN connectivity that was within normal limits. fMRI biomarkers included the presence, spatial extent (percent of suprathreshold voxels in an ROI), and magnitude (z-statistic) of a response to the active-motor-imagery and passive-language tasks and the presence (within normal limits) and strength (z-statistic) of the DMN.

### Statistical Analysis

First, we determined whether the CRS-R diagnosis was associated with 6-month DRS scores using the Kruskal-Wallis test. Then, we tested whether participants with evidence of command-following on either CRS-R, EEG, or fMRI (Composite Measure based on evidence of command-following, CM_Command_) had lower 6-month DRS total scores than those who did not have evidence of command-following on any measure using the Wilcoxon rank sum test. We estimated the DRS median difference between groups and associated 95% confidence interval using the Hodges-Lehman approach.^52^ The approach of using evidence of command-following by any measure is consistent with the European Academy of Neurology DoC clinical guidelines, which recommend that DoC diagnosis be based on the highest level of consciousness identified by any assessment within a multimodal battery.^34^ Outcomes in participants in the CM_Command_ group (CRS-R behavioral diagnosis of MCS+ *or* PTCS [on one or both CRS-R exams], *or* positive responses to active-motor-imagery EEG, *or* positive responses to active-motor-imagery fMRI) were compared to participants with a behavioral diagnosis of coma, vegetative state/unresponsive wakefulness syndrome (VS/UWS), *or* minimally conscious state minus (MCS-), *and* negative responses to active-motor-imagery EEG, *and* negative responses to active-motor-imagery fMRI. For this analysis, participants could have a maximum of 4 (i.e., 2 CRS-R, 1 EEG, and 1 fMRI) and a minimum of 2 (i.e., 1 CRS-R and 1 EEG or 1 fMRI) command-following assessments.

The presence of command-following on behavioral assessment^1,5,53^ or task-based EEG/fMRI has been associated with better outcomes after severe brain injury.^13,14,31^ However, it is less clear whether recovery of lower-level signs of consciousness (i.e., MCS-, evidenced by behaviors such as visual pursuit), is also predictive of outcome.^3,54^ We therefore also tested whether participants with evidence of conscious awareness (regardless of command-following ability) by any assessment method (Composite Measure based on evidence of consciousness, CM_Conscious_) had lower 6-month DRS total scores than those who did not have evidence of consciousness on any measure (i.e., CRS-R behavioral diagnosis of MCS-, *or* MCS plus (MCS+), *or* post-traumatic confusional state (PTCS) [on one or both CRS-R exams], *or* positive responses to active-motor-imagery EEG *or* fMRI versus participants with a behavioral diagnosis of coma *or* VS/UWS, *and* negative responses to active-motor-imagery EEG and fMRI). Participants could have a maximum of 4 (i.e., 2 CRS-R, 1 EEG, and 1 fMRI) and a minimum of 2 (i.e., 1 CRS-R and 1 EEG or 1 fMRI) assessments of command-following (CM_Command_) and consciousness (CM_Conscious_).

To determine which individual EEG and fMRI variables were associated with 6-month DRS total scores, we calculated univariate Spearman’s correlation coefficients for continuous predictors or Wilcoxon rank sum tests for dichotomous predictors separately for participants with EEG assessments and for participants with fMRI assessments. Bootstrap confidence intervals for Spearman’s correlation coefficients were calculated, and a 95% confidence interval for the Hodges-Lehman estimate of the median difference is reported along with the Wilcoxon rank sum test p-values. We scaled continuous variables and treated dichotomous variables as factors with 2 levels.

Given that multimodal assessment for outcome prediction is most relevant for patients who have not recovered command-following we repeated these analyses in a subsample of participants with a CRS-R diagnosis of coma, VS/UWS, or MCS-at the time of EEG or fMRI. We also repeated all of analyses above with the secondary outcome measures. For outcomes that were dichotomized, we used logistic regression for both dichotomous and continuous predictors, and we report odds ratios, 95% confidence intervals and p-values. Given the large number of secondary outcomes and predictor variables, we did not correct for multiple comparisons. We used R Statistical Software (v4.3.2; 2023-10-31) for all analyses.

## Results

### Data availability and visualization

To facilitate data transparency, we release an interactive, comprehensive fMRI data visualization tool hosted at [GITHUB Link TBD] that can be downloaded at [ZOTERO Link TBD, temporarily hosted <u>here</u>]. Using INVISION, active-motor-imagery, passive-language, and rs-fMRI data can be investigated interactively to enhance the interpretability of our results. We believe access to data at this level of granularity is consistent with the mission of the Open Science Framework^55^ as it facilitates transparency and reproducibility.

### Demographics and clinical characteristics

Of 770 patients with TBI admitted to one of three ICUs between 09/01/2018 and 12/29/2022, 582 were excluded. The most common exclusion criterion was absence of a GCS total score <9 within 24 hours of admission (N=430, Figure 1). Among 187 eligible patients, 133 were not enrolled. The most common reasons eligible patients were not enrolled were: death prior to approach for consent (N=57 died after WLST, N=8 died without WLST), surrogate refusal (N=27), and COVID-19 (N=14 admitted to the ICU during the COVID-19 pandemic when in-person research was not permitted, N=6 admitted after research activities resumed but tested positive for COVID-19 infection).

**Figure 1:**
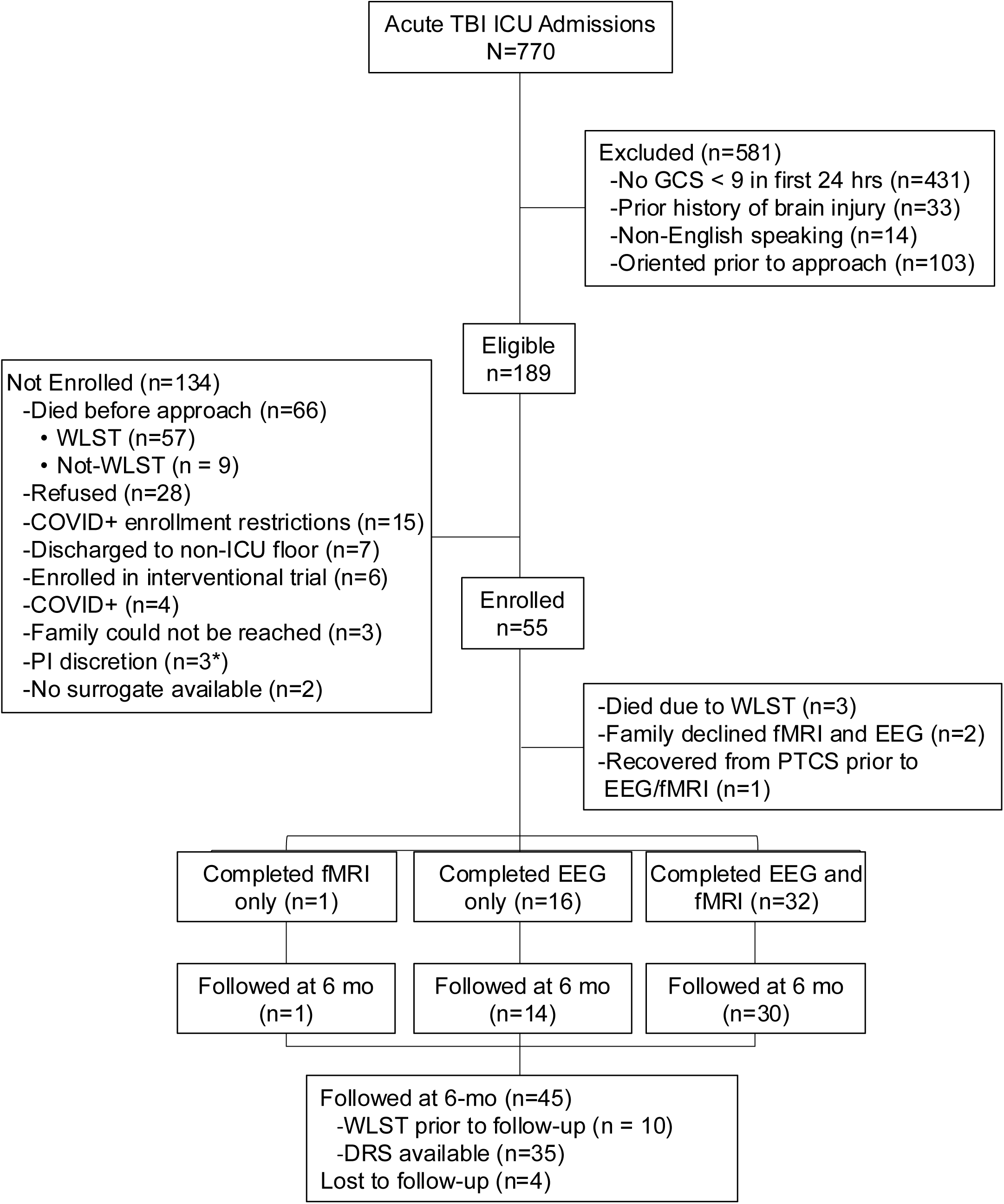
CONSORT Diagram of Participant Enrollment. Patients with traumatic brain injury (TBI) admitted to one of three intensive care units (ICU) at Massachusetts General Hospital between 09/01/2018 - 12/29/2022 were screened for eligibility. Of 770 admissions, the majority were excluded due to the absence of severe TBI (i.e., no Glasgow Coma Scale score < 9 in the first 24 hours, or recovery of orientation prior to approach for enrollment). Among eligible participants, the most common factor for non-enrollment was death due to withdrawal of life-sustaining treatment. We enrolled 55 participants, of whom 49 were assessed with either task-based electroencephalography, functional magnetic resonance imaging, or both, and 45 were followed at 6-months post-TBI. Abbreviations: DRS *Disability Rating Scale;* EEG *electroencephalography*; fMRI *functional magnetic resonance imaging*; GCS *Glasgow Coma Scale;* mo *months;* PI *principal investigator;* PTCS *post-traumatic confusional state; WLST withdrawal of life-sustaining treatment*.

We obtained written informed consent from the surrogates of 55 patients; N=3 died from WLST prior to the start of research activities, N=2 did not receive EEG or fMRI due to family refusal, N=1 recovered from PTCS and was alert and oriented prior to EEG/fMRI; and N=4 could not be reached for follow-up at 6-months post-TBI. The final sample of eligible patients who were assessed with fMRI, EEG, or both, and for whom 6-month outcome was available, was 45 (N=30 with EEG and fMRI, N=14 with EEG only, and N=1 with fMRI only; N=35 survived to 6-months, N=10 died in the ICU due to WLST). In this final sample of N=45, N=31 (68.9%) were male and the mean±SD age was 45.2±20.7 years (Table 1, Supplementary Table 4). The distribution of CRS-R and DRS scores is illustrated in Supplementary Figures 1 and 2 for the full sample and in Supplementary Figure 3 for participants who survived to 6-months.

**Table 1:** Participant EEG and fMRI responses to active-motor imagery, passive-language, and rest.

| ID | EEG Variables |  |  |  | fMRI Variables |  |  |  |  |  |  | 6 mo Outcome |  |
| --- | --- | --- | --- | --- | --- | --- | --- | --- | --- | --- | --- | --- | --- |
|  | Day of EEG | CRS-R at EEG | Active-Motor +/- (Accuracy) | Passive-Language +/- (Accuracy) | Day of fMRI | CRS-R at fMRI | Active-Motor Paradigm |  | Passive-Language Paradigm |  | Resting-state +/- DMN (Z statistic) | DRS Total | GOSE-ALL/TBI |
|  |  |  |  |  |  |  | +/- (% SMA/PMC voxels) | SMA/PMC Z statistic | +/- (% STG Voxels) | STG Z statistic |  |  |  |
| P1 | 17 | coma | - (0.47) | - (0.47) | NA | NA | NA | NA | NA | NA | NA | 30 | 1 |
| P2<sup>a</sup> | 4 | coma | + (0.54) | + (0.86) | 5 | coma | + (0.19) | -0.13 | + (34.15) | 2.39 | +(0.87) | 5 | 4/4 |
| P3<sup>a</sup> | 5 | coma | + (0.62) | + (0.61) | NA | NA | NA | NA | NA | NA | NA | 30 | 1 |
| P4<sup>a</sup> | 8 | coma | + (0.55) | + (0.57) | 9 | coma | + (0.50) | 0.28 | + (2.27) | 0.46 | -(0.66) | 30 | 1 |
| P5 | 2 | coma | - (0.50) | + (0.67) | 9 | coma | - (0) | 0.10 | + (5.58) | 0.50 | 0.52 | 12 | 3/3 |
| P6 | 3 | coma | - (0.52) | + (0.75) | NA | NA | NA | NA | NA | NA | NA | 30 | 1 |
| P7 | 26 | VS/UWS | - (0.54) | NA<sup>c</sup> | 25 | VS/UWS | - (0) | 0.26 | + (6.00) | 0.73 | +(0.86) | 30 | 1 |
| P8 | 5 | VS/UWS | - (0.51) | + (0.60) | 5 | VS/UWS | - (0) | -0.54 | + (34.74) | 2.61 | +(1.17) | 2 | 7/8 |
| P9 | 6 | VS/UWS | - (0.51) | + (0.70) | 6 | VS/UWS | - (0) | -0.19 | + (5.91) | -0.19 | +(1.13) | 6 | 3/8 |
| P10 | 4 | VS/UWS | - (0.53) | - (0.53) | 3 | VS/UWS | - (0) | -0.21 | - (0) | -0.61 | +(0.84) | 2 | 6/7 |
| P11 | 12 | VS/UWS | - (0.54) | + (0.60) | 11 | VS/UWS | - (0) | 0.28 | + (35.42) | 2.69 | 0.53 | 14 | 3/3 |
| P12<sup>a</sup> | 2 | VS/UWS | - (0.50) | + (0.58) | 9 | VS/UWS | + (2.38) | 0.46 | + (5.48) | 0.78 | +(1.45) | 16 | 3/3 |
| P13<sup>a</sup> | 2 | VS/UWS | + (0.59) | + (0.70) | 2 | VS/UWS | + (0.77) | 0.69 | + (12.40) | 0.99 | 0.25 | 9 | 4/4 |
| P14 | 14 | VS/UWS | - (0.53) | + (0.79) | NA | NA | NA | NA | NA | NA | NA | 11 | 3/3 |
| P15 | 3 | VS/UWS | - (0.51) | + (0.58) | 4 | VS/UWS | - (0) | -0.35 | + (0.88) | -0.07 | 0.28 | 30 | 1 |
| P16 | 3 | VS/UWS | - (0.54) | + (0.54) | NA | NA | NA | NA | NA | NA | NA | 0 | 8/8 |
| P17 | 1 | VS/UWS | - (0.53) | + (0.60) | NA | NA | NA | NA | NA | NA | NA | 30 | 1 |
| P18 | 2 | VS/UWS | - (0.52) | + (0.69) | NA | NA | NA | NA | NA | NA | NA | 0 | 6/8 |
| P19<sup>a</sup> | 6 | VS/UWS | - (0.51) | + (0.56) | 17 | VS/UWS | + (0.20) | -0.10 | + (0.66)<sup>a</sup> | -0.11 | 0.40 | 23 | 3/3 |
| P20 | 3 | VS/UWS | - (0.51) | + (0.55) | 4 | VS/UWS | - (0) | -0.04 | - (0) | -0.23 | 0.67 | 30 | 1 |
| P21 | 6 | VS/UWS | - (0.48) | + (0.56) | 16 | VS/UWS | - (0) | -0.21 | - (0) | -0.32 | 0.67 | 22 | 2/2 |
| P22<sup>a</sup> | 5 | VS/UWS | NA<sup>d</sup> | - (0.52) | 8 | VS/UWS | + (0.04) | -0.06 | - (0) | -0.30 | 0.72 | 1 | 8/8 |
| P23 | 5 | VS/UWS | - (0.52) | + (0.60) | 4 | VS/UWS | - (0) | -0.15 | -(0) | -0.10 | 0.53 | 23 | 2/2 |
| P24 | 34 | VS/UWS | - (0.48) | + (0.75) | 34 | VS/UWS | - (0) | -0.60 | +(20.28) | 1.27 | +(1.03) | 4 | 5/5 |
| P25 | 11 | VS/UWS | - (0.50) | + (0.62) | 11 | VS/UWS | - (0) | -0.24 | +(6.80) | 1.37 | 0.42 | 30 | 1 |
| P26<sup>a,b</sup> | 2 | MCS- | + (0.57) | + (0.56) | 2 | MCS- | + (0.16) | -0.16 | - (0) | -0.46 | 0.63 | 11 | 3/3 |
| P27<sup>b</sup> | 2 | MCS- | - (0.53) | + (0.61) | 4 | VS/UWS | - (0) | -0.46 | + (4.65) | -0.02 | 0.57 | 1 | 5/5 |
| P28<sup>a,b</sup> | 18 | MCS- | + (0.57) | + (0.63) | NA | NA | NA | NA | NA | NA | NA | 30 | 1 |
| P29<sup>b</sup> | 15 | MCS- | - (0.49) | + (0.67) | NA | NA | NA | NA | NA | NA | NA | 1 | 6/6 |
| P30<sup>a,b</sup> | 2 | MCS- | - (0.52) | + (0.60) | 6 | MCS- | + (4.16) | 0.50 | + (34.44) | 2.59 | +(0.86) | 3 | 5/6 |
| P31<sup>b,e</sup> | 3 | MCS- | - (0.50) | + (0.81) | 8 | eMCS | - (0) | 0.10 | + (18.35) | 1.44 | 0.47 | 5 | 3/3 |
| P32<sup>b</sup> | 6 | MCS- | - (0.52) | + (0.64) | 10 | MCS- | - (0) | -0.24 | + (1.97) | 0.10 | 0.59 | 3 | 4/4 |
| P33<sup>b</sup> | 15 | MCS- | - (0.50) | NA<sup>c</sup> | NA | NA | NA | NA | NA | NA | NA | 13 | 3/3 |
| P34<sup>b</sup> | 8 | MCS- | - (0.53) | + (0.78) | 23 | MCS- | - (0) | -0.83 | + (19.63) | 0.95 | +(0.82) | 14 | 3/3 |
| P35<sup>b</sup> | 10 | MCS- | - (0.45) | + (0.66) | 11 | MCS- | - (0) | 0.21 | + (14.34) | 1.24 | +(1.14) | 0 | 6/6 |
|  | Day of EEG | CRS-R at EEG | Active-Motor +/- (Accuracy) | Passive-Language +/- (Accuracy) | Day of fMRI | CRS-R at fMRI | Active-Motor Paradigm |  | Passive-Language Paradigm |  | Resting-state | DRS Total | GOSE-ALL/TBI |
|  |  |  |  |  |  |  | +/- (% SMA/PMC voxels) | SMA/PMC Z statistic | +/- (% STG Voxels) | STG Z statistic | +/- DMN (Z statistic) |  |  |
| P36<sup>a,b</sup> | 15 | MCS+ | - (0.53) | + (0.61) | 21 | MCS+ | - (0) | -0.21 | + (4.17) | 0.46 | 0.47 | 2 | 5/5 |
| P37<sup>a,b</sup> | 9 | MCS+ | - (0.51) | + (0.58) | 9 | MCS+ | - (0) | 0.16 | + (15.94) | 1.24 | 0.58 | 0 | 8/8 |
| P38<sup>a,b</sup> | 6 | MCS+ | - (0.50) | + (0.62) | NA | NA | NA | NA | NA | NA | NA | 3 | 5/5 |
| P39<sup>a,b,f</sup> | 6 | MCS+ | - (0.51) | + (0.74) | 8 | eMCS | NA | NA | NA | NA | 0.65 | 10 | 3/3 |
| P40<sup>a,b</sup> | 4 | MCS+ | + (0.56) | + (0.66) | 15 | MCS+ | - (0) | -0.09 | + (15.76) | 1.33 | 0.72 | 3 | 4/4 |
| P41<sup>a,b</sup> | 4 | MCS+ | - (0.46) | + (0.59) | NA | NA | NA | NA | NA | NA | NA | 1 | 5/7 |
| P42<sup>a,b</sup> | 5 | MCS+ | - (0.51) | + (0.72) | 5 | MCS+ | - (0) | 0.07 | + (6.85) | 0.77 | +(1.03) | 3 | 6/6 |
| P43<sup>a,b</sup> | 6 | eMCS | - (0.50) | + (0.76) | NA | NA | NA | NA | NA | NA | NA | 8 | NA<sup>g</sup> |
| P44<sup>a,b</sup> | 17 | eMCS | + (0.59) | + (0.77) | NA | NA | NA | NA | NA | NA | NA | 5 | 3/3 |
| P45<sup>a,b</sup> | NA | NA | NA | NA | 3 | eMCS | - (0) | -0.22 | + (48.95) | 3.43 | +(0.86) | 0 | 8/8 |

<sup>a</sup> participant meeting criteria for command following composite measure (CM<sub>Command</sub>; CRS-R diagnosis of MCS+, or eMCS, or positive response to active-motor fMRI, or positive response to active-motor EEG)
<sup>b</sup> participant meeting criteria for consciousness composite measure (CM<sub>Conscious</sub>; CRS-R diagnosis of MCS-, or MCS+, or eMCS, or positive response to active-motor fMRI, or positive response to active-motor EEG)
<sup>c</sup> Two or three (out of three) blocks of data were discarded due to breaks in the acquisition paradigm which resulted in a time lapse between rest and language blocks that exceeded 3 minutes, compromising the data analysis and interpretation
<sup>d</sup> Paradigm instructions were not played
<sup>e</sup> P31 had a CRS-R diagnosis of MCS- at the time of EEG
<sup>f</sup> participant crawled out of the scanner after completion of resting-state fMRI scan, thus hand and language fMRI paradigms were not acquired
<sup>g</sup> DRS was completed, for GOSE could not be obtained at the time of follow-up and participant could not be reached subsequently
Participants with a CRS-R diagnosis of coma, VS/UWS, or MCS- who have positive (+) responses to the active-motor task on EEG or fMRI are considered to have cognitive motor dissociation (CMD); Participants with a CRS-R diagnosis of coma, VS/UWS, or MCS- who have positive (+) responses to the passive-language task on EEG or fMRI are considered to have covert cortical processing (CCP)
Abbreviations: CRS-R *Coma Recovery Scale-Revised*; DMN *default mode network*; DRS *Disability Rating Scale*; EEG *electroencephalography*; fMRI *functional magnetic resonance imaging*; GOSE TBI/ALL *Glasgow Outcome Scale-Extended considering only the effects of all injuries [GOSE-All], and the effects of the TBI only [GOSE-TBI]*; mo *month*; N/A *not applicable*; SMA/PMC *supplementary motor area/premotor cortex*; STG *superior temporal gyrus*

EEG was performed on average two days prior to fMRI (EEG mean±SD=7.8±6.9 days post-injury; fMRI 10.0±7.5 days post-injury, p>0.05). EEG was performed within 24 hours of fMRI in 17 participants (Table 1). In these participants, only one CRS-R assessment was conducted; otherwise, a CRS-R assessment was conducted within 24-hours of EEG and separately within 24-hours of fMRI. Among the 45 participants who were assessed 64 times with the CRS-R, nine CRS-R assessments (in N=8 participants) had at least 1 invalid CRS-R subscale (see Supplementary Materials, Supplementary Table 5). EEG data were discarded for the active-motor-imagery paradigm of one patient participant, and the passive-language paradigm of 2 patients because two runs of EEG data were not available (Table 1). At the time of EEG, CRS-R assessment indicated coma (N=6), VS/UWS (N=19), MCS-(N=10), MCS+ (N=7) or PTCS (N=2). At the time of fMRI, CRS-R examination indicated coma (N=3), VS/UWS (N=16), MCS-(N=5), MCS+ (N=4) or PTCS (N=3).

Among the N=44 patient participants assessed with EEG, positive responses to the active-motor-imagery paradigm were observed in 8/43 (18.6%; n=1 was excluded due to data acquisition limitations, Table 1, Figure 2) and to the passive-language EEG paradigm in 39/42 (92.9%, n=2 were excluded due to data acquisition limitations). Among the N=35 subsample of participants with a CRS-R diagnosis of coma, VS/UWS, or MCS-at the time of EEG, a positive response to the active-motor-imagery EEG paradigm (i.e., CMD) was detected in 6/34 (16.2%) and to the passive-language EEG paradigm (i.e., CCP) in 30/33 (90.0%).

**Figure 2:**
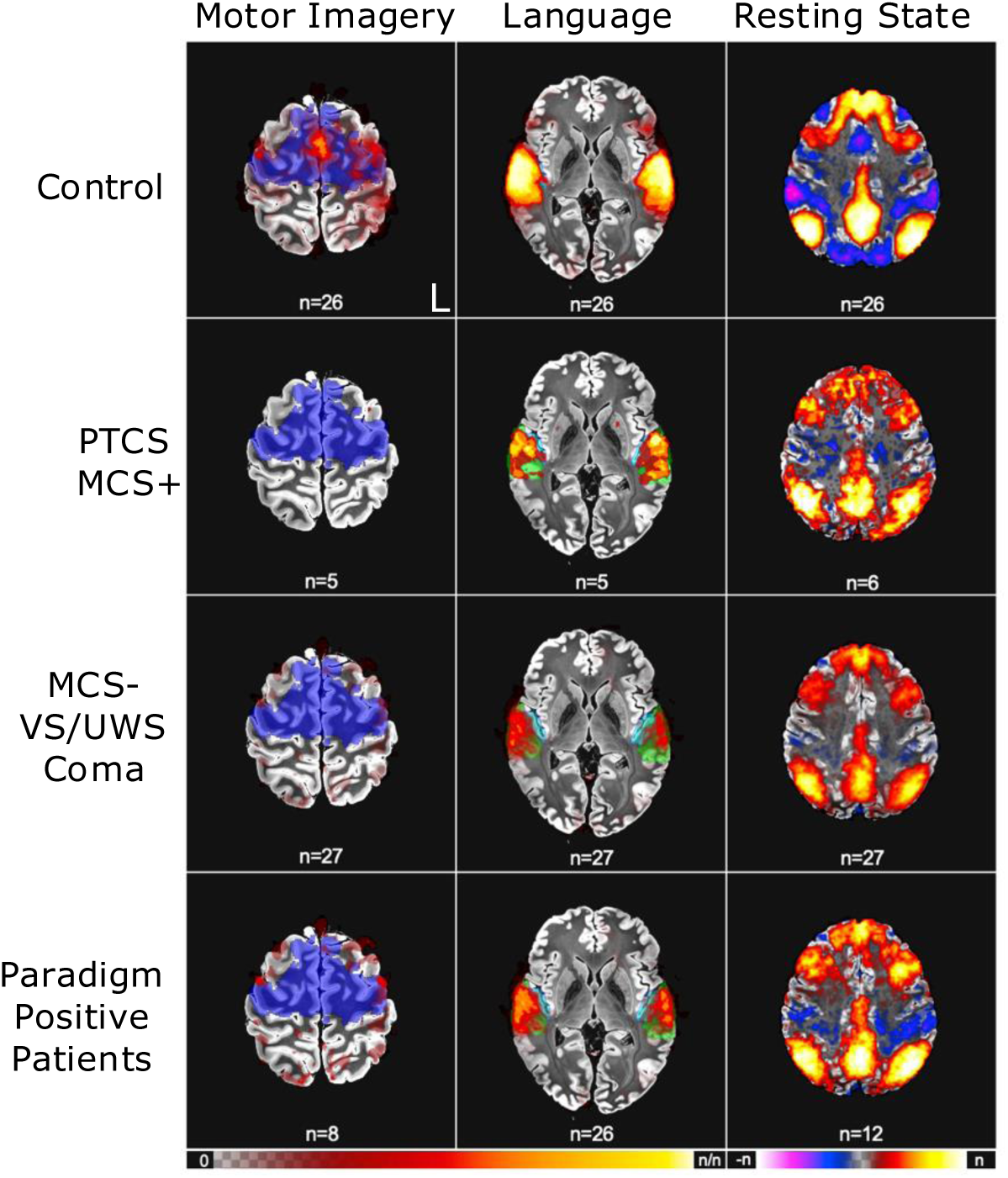
fMRI Group-level Results. Functional magnetic resonance imaging (fMRI) results at the group-level are shown for the task-based fMRI motor-action paradigm, stimulus-based passive-language paradigm, and resting-state fMRI (columns) classified across the rows by healthy control participants, patient participants with a CRS-R diagnosis of MCS+ or eMCS, and patients with a CRS-R diagnosis of MCS-, VS/UWS, or coma. The last row includes only the patient participants with a positive response to the task-based fMRI motor-action paradigm or stimulus-based passive-language paradigm, or within normal limits (i.e., within 95% confidence interval of healthy control participants) response on resting-state fMRI. The blue area in the Active-motor-imageryand Passive-Language fMRI columns illustrate the regions of interest encompassing the supplementary motor area/premotor cortex (SMA/PMC) and superior temporal gyrus (STG), respectively. Red-yellow overlays in the Active-motor-imageryand Passive-Language columns represent heatmaps of the percentage of participants (out of the total indicated in each cell) that have a positive response in the voxels shown. In the Resting State fMRI column, voxel color and intensity are modulated by the percentage of participants in each group with default mode network (DMN) correlations (red/yellow) or anticorrelations (purple/blue). No participants with a PTCS or MCS+ diagnosis had a positiveF response to active-motor-imagery fMRI. Among participants with a diagnosis of coma, VS/UWS, or MCS-, n=8 had a positive response to active-motor-imagery fMRI. Abbreviations: MCS-/MCS+ *minimally conscious state without/with language function*; PTCS *post-traumatic confusional state;* VS/UWS *vegetative state/unresponsive wakefulness syndrome*.

Among N=31 participants with fMRI data, positive responses to the active-motor-imagery fMRI paradigm were observed in 8/30 (26.7%; n=1 completed the rs-fMRI scan but became agitated and could not complete the scan, Table 1, Figure 2) and to the passive-language fMRI paradigm in 24/30 (80.0%). Compared to a sample of healthy control participants, DMN connectivity was within normal limits in 12/31 (38.7%). Among the N=24 subsample of participants with a CRS-R diagnosis of coma, VS/UWS, and MCS-, positive responses to the active-motor-imagery fMRI paradigm (i.e., CMD) was detected in 8/24 (33.3%) and to the passive-language fMRI paradigm (i.e., CCP) in 18/24 (75.0%). DMN connectivity was within normal limits in 10/24 (41.7%).

EEG and fMRI responses and connectivity can be detected even in patients with DoC receiving sedating interventions.^12,13,29,30^ Therefore, we included participants receiving these interventions and provide the types and doses of sedative, anxiolytic, and analgesic medications administered at the time of assessment in Supplementary Table 6. we provide this information for descriptive purposes. To illustrate the medical acuity of patients included in this study, we list each participant’s drains, lines, tubes, and monitors at the time of EEG and fMRI in Supplementary Table 7. Results for healthy participants are in Supplementary Materials and Supplementary Table 8, and results for patient participants who were not followed at 6-months are in Supplementary Table 9. Individual participant data on secondary outcomes is in Supplementary Table 10.

### Age and behavioral level of consciousness on the CRS-R are associated with outcome

Younger age (continuous variable) and DoC behavioral diagnosis (categorical variable representing the best CRS-R diagnosis that was available at the time of EEG or fMRI) were associated with significantly lower (i.e., better) 6-month DRS total scores (age: ρ [95%CI] 0.396 [0.070, 0.648]; p=0.007; CRS-R: p=0.014, Supplementary Table 11). Pairwise group comparisons indicated that the DRS total score at 6-months was significantly lower in patients with a CRS-R diagnosis of MCS-versus coma (DRS difference in medians [CI]= 17[0, 29], p=0.035), MCS+ versus coma (27[5,29], p=0.004), PTCS versus coma (22[2, 25]; p=0.030) and in patients with MCS+ versus VS/UWS (11.7[1,27]; p=0.029, Figure 3). Only the MCS+ versus coma comparison survived Bonferroni multiple comparison correction. Similarly, DRS total scores were significantly lower in patients who were following commands (MCS+, PTCS) versus those who were not following commands (coma, VS/UWS) (-9[-20, -1]; p=0.011) and in those who were conscious (MCS-, MCS+, PTCS) versus those who were not conscious (coma, VS/UWS) (-11[-20, -2]; p=0.003) on the CRS-R (Table 2). Age and consciousness assessed by the CRS-R were also significantly associated with secondary outcome measures, although these associations were rarely observed in the subsample of participants surviving to 6-months (Supplementary Tables 12, 13, 14, 15, and 16).

**Table 2:** Association of CRS-R, EEG, and fMRI assessment variables with 6-month DRS total score.

| Variable (n) | Wilcoxon rank sum test<br>Estimate [95% CI]; p-value |
| --- | --- |
| **CM<sub>Command</sub>** (n=45) |  |
| CRS-R<br>y (n=11)<br>n (n=34) | -9 [-20, -1]; p=0.011 |
| CRS-R or EEG<br>y (n=17)<br>n (n=28) | -2 [-12, 2]; p=0.334 |
| CRS-R or fMRI<br>y (n=19)<br>n (n=26) | -7 [-19, 0]; p=0.067 |
| CRS-R or EEG or fMRI<br>y (n=21)<br>n (n=24) | -2 [-12, 2]; p=0.313 |
| **CM<sub>Conscious</sub>** (n=45) |  |
| CRS-R<br>y (n=20)<br>n (n=25) | -11 [-20, -2]; p=0.003<sup>a</sup> |
| CRS-R or EEG<br>y (n=24)<br>n (n=21) | -9 [-19, 0]; p=0.030<sup>a</sup> |
| CRS-R or fMRI<br>y (n=26)<br>n (n=19) | -10.67 [-20, -1]; p=0.013<sup>a</sup> |
| CRS-R or EEG or fMRI<br>y (n=27)<br>n (n=18) | -8 [-19, 0]; p=0.037<sup>a</sup> |
| <sup>a</sup> Statistically significant associations with 6-month DRS total score (p<0.05)<br>Estimate reported is the Hodges-Lehman estimate of the median difference between a measurement from group 1 and a measurement from group 2.<br>Abbreviations: CI confidence interval; CM<sub>Command</sub> Composite Measure including any assessment indicating command-following; CM<sub>Conscious</sub> Composite Measure including any assessment indicating conscious awareness; CRS-R Coma Recovery Scale-Revised; DRS Disability Rating Scale; EEG electroencephalography, fMRI functional magnetic resonance imaging |  |

**Figure 3:**
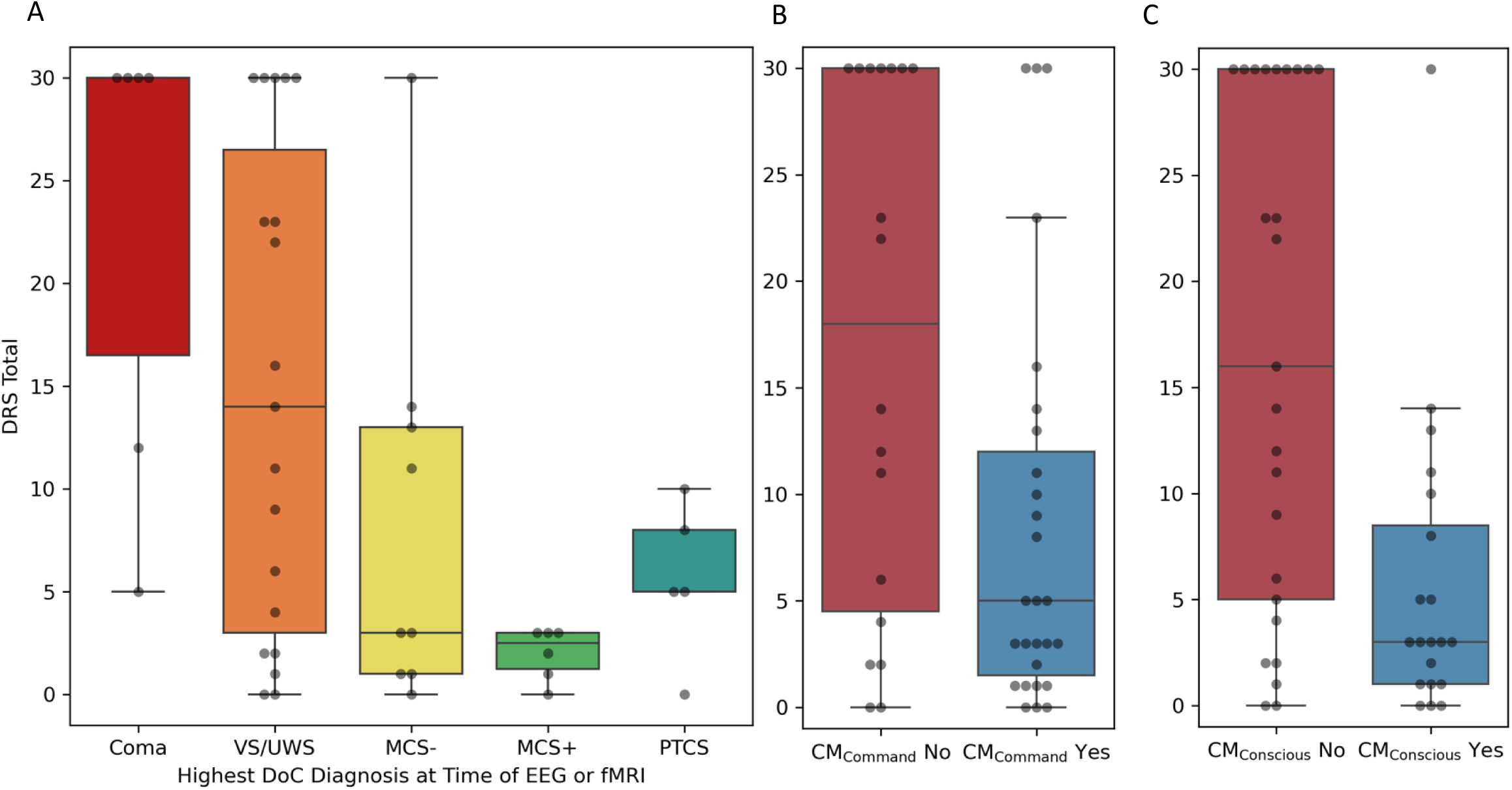
Disability Rating Scale at 6-months in Participants Across Behavioral Diagnosis and Composite Measures of Command-following and Consciousness. The 6-month DRS score is plotted for participants with each behavioral diagnosis based on the Coma Recovery Scale-Revised (CRS-R, [A]) and for participants who have evidence of command-following (CM_Command_: CRS-R diagnosis of MCS+, or PTCS, or positive response to active-motor-imagery EEG, or positive response to active-motor-imagery fMRI, [B]), or evidence of consciousness (CM_Conscious_: CRS-R diagnosis of either MCS-, or MCS+, or PTCS, or positive response to active-motor-imagery EEG, or positive response to active-motor-imagery fMRI [C]) on a composite measure. The behavioral diagnosis is based on the best CRS-R diagnosis obtained at the time of EEG or fMRI. Medians are indicated by a solid line and means are indicated by a dotted line in each box plot. Abbreviations: CM_Conscious_ *composite measure of consciousness;* CM_Command_ *composite measure of command-following;* DoC *disorders of consciousness;* MCS-/MCS+ *minimally conscious state without/with language function*; VS/UWS *vegetative state/unresponsive wakefulness syndrome;* PTCS *post-traumatic confusional state*.

**Figure 4:**
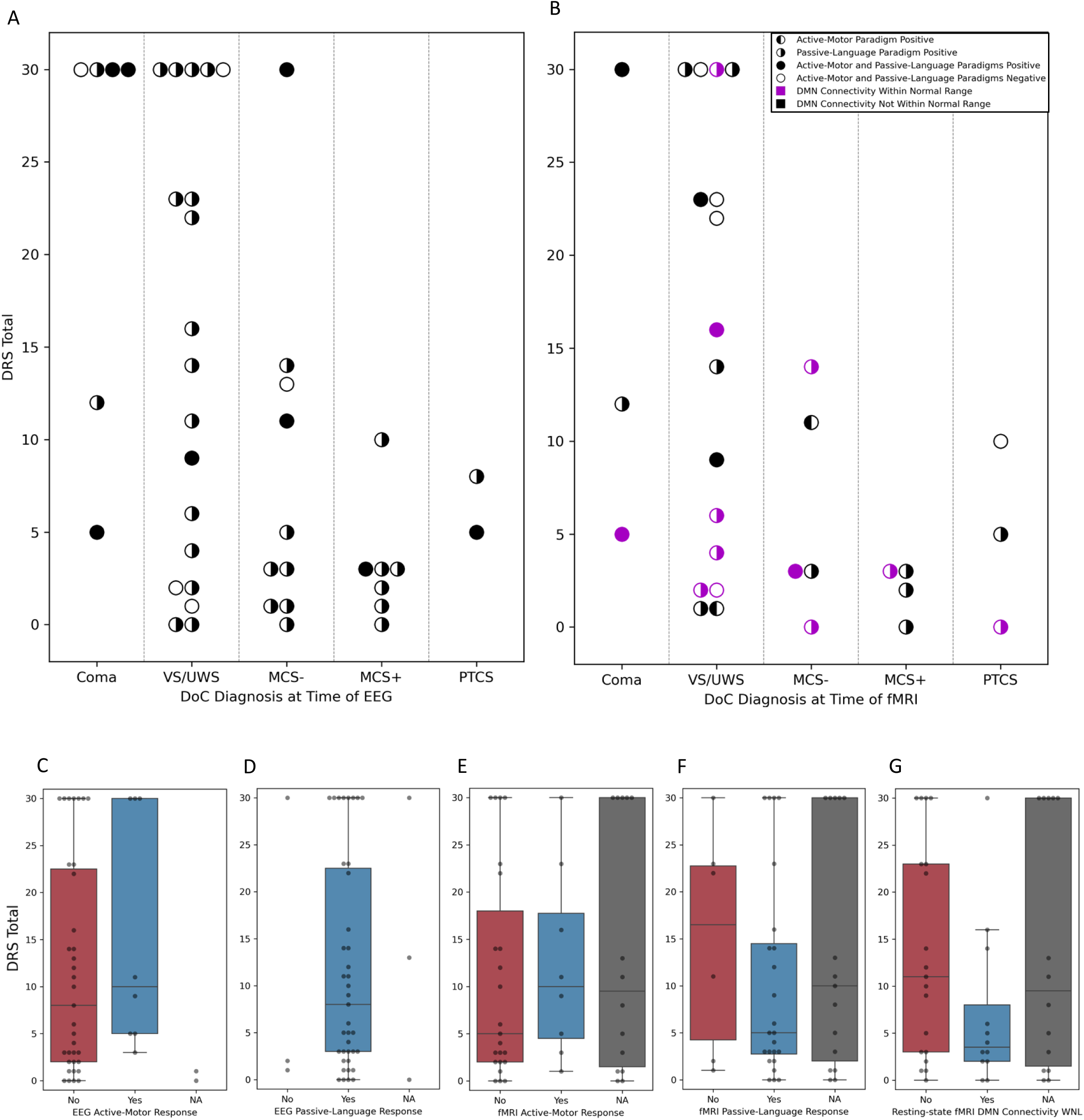
Disability Rating Scale at 6-months in Participants with and without Responses to Active-motor-imagery and Passive-language EEG and fMRI. The 6-month DRS score is plotted for each participant with the circles shaded in black to indicate whether responses to the active-motor, passive-language, or both were detected on EEG (A) and fMRI (B). In panel B, the presence of intact default mode network (DMN) connectivity is further indicated with purple shading. Participants are ordered on the x-axis based on Coma Recovery Scale-Revised (CRS-R) diagnosis at the time of the EEG (A) or fMRI (B) assessment. Panels C-G illustrate group-level results for DRS total score based on the presence of a response to active-motor-imagery EEG (C), passive-language EEG (D), active-motor-imagery fMRI (E), passive-language fMRI (F), and intact DMN connectivity (G). Box plots are omitted when there are less than 6 data points in a group (e.g., no response to passive-language EEG, [D]). NA indicates that the data were not acquired (see Table 1). Abbreviations: DoC *disorders of consciousness;* MCS-/MCS+ *minimally conscious state without/with language function*; NA *not acquired;* PTCS *post-traumatic confusional state;* VS/UWS *vegetative state/unresponsive wakefulness syndrome;* WNL *within normal limits (i.e., within 95% confidence interval of healthy control participants)*.

**Figure 5:**
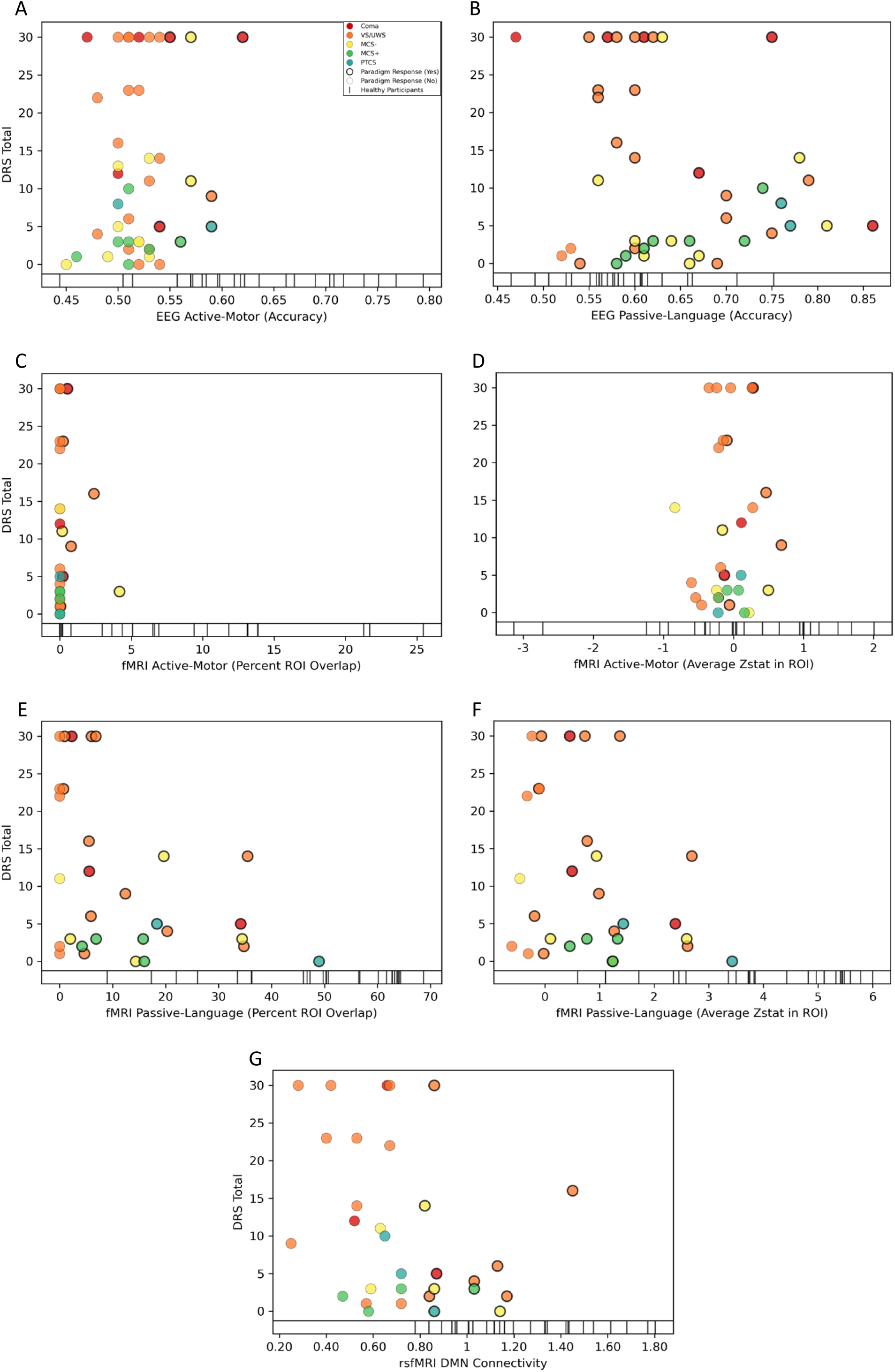
Disability Rating Scale at 6-months in Participants with Varying Degree of Responsiveness to Active-motor-imagery and Passive-language EEG and fMRI. The 6-month DRS score is plotted for each participant with the x-axis depicting the accuracy of the classifier for the active-motor-imagery (A) and passive-language (B) paradigms, percent of suprathreshold voxels within the supplementary motor area/premotor cortex (SMA/PMC, [C]) and z-statistic within the SMA/PMC (D) for the active-motor-imagery fMRI paradigm, percent of suprathreshold voxels within the superior temporal gyrus (STG, [E]) and z-statistic within the STG (F) for the passive-language fMRI paradigm, and z-statistic within the default mode network (DMN, [G]). Dot colors represent behavioral diagnosis based on the Coma Recovery Scale-Revised (CRS-R). A thick outline indicates the response to the paradigm was positive. At the bottom of each scatterplot, vertical tick-marks represent healthy control data. Abbreviations: DoC *disorders of consciousness;* MCS-/MCS+ *minimally conscious state without/with language function*; NA *not acquired;* PTCS *post-traumatic confusional state;* VS/UWS *vegetative state/unresponsive wakefulness syndrome;* WNL *within normal limits (i.e., within 95% confidence interval of healthy control participants)*.

### Multimodal composite measures of command-following and consciousness associated with outcome

There was no statistical difference in 6-month DRS total score for participants with versus without command-following when the CRS-R was combined with results of active-motor-imagery EEG, fMRI, or both (CM_Command_; Table 2, Figure 2). DRS total scores were significantly lower for participants with evidence of consciousness on either the CRS-R or active-motor-imagery EEG responses, the CRS-R or active-motor-imagery fMRI, and the CRS-R or active-motor-imagery EEG or fMRI (i.e., components of CM_Conscious_; CRS-R or EEG: -9 [-19, 0], p=0.030; CRS-R or fMRI: -10.67 [-20, -1], p=0.013; CRS-R or fMRI: -8 [-19, 0], p=0.037; Table 2). Supplementary Figure 3 illustrates results in the subsample of participants who survived to 6 months. Evidence of consciousness based on components of CM_Conscious_ was also associated with significantly better outcome on DRS_Depend_ (CRS-R or EEG: 4[1.15, 13.86]; p=0.029; CRS-R or fMRI: 4.09 [1.16, 14.43]; p=0.028), the continuous GOSE-All (CRS-R or fMRI: 2 [0, 3]; p=0.018; CRS-R or fMRI or EEG: 2 [0, 3]; p=0.049) and dichotomous GOSE-All (CRS-R or EEG: 3.89 [1.1, 13.76]; p=0.035; CRS-R or fMRI: 4.2 [1.15, 15.37]; p=0.03, Supplementary Table 12).

### EEG and fMRI biomarkers associated with 6-month DRS and secondary outcomes

Individual EEG biomarkers were not significantly associated with 6-month DRS total in the full sample, in subsamples, or with any secondary outcome measure (Table 3, Supplementary Tables 13, 15). Individual fMRI biomarkers were also not significantly associated with 6-month DRS total in the full sample (Table 3). However, in participants with a CRS-R diagnosis of coma, VS/UWS, or MCS-, a higher z-statistic within the DMN was associated with a significantly lower 6-month DRS total score (-0.411 [-0.707, - 0.027]; p=0.046, Supplementary Table 16). In the full sample, rs-fMRI DMN connectivity was associated with better outcomes on DRS_Depend_ (DMN connectivity within normal limits: 5.14 [1.03, 25.6]; p=0.046; DMN z-statistic: 32.4 [1.06, 990.42]; p=0.046) and continuous GOSE-TBI (DMN connectivity within normal limits: 2 [0, 4]; p=0.019; DMN z-statistic: 0.436 [0.140, 0.713]; p=0.014, Supplementary Table 14). Similar results were observed for the GOSE-All outcome and in the subsample of patients surviving to 6-months (Supplementary Figure 4, Supplementary Tables 14, 16). A higher percent of suprathreshold voxels in the STG ROI in response to passive-language fMRI was associated with better GOSE-All total scores (0.366 [0.01, 0.67]; p=0.047) and a similar trend was observed for the GOSE-TBI outcome (Supplementary Table 14).

**Table 3:** Association of CRS-R, EEG, and fMRI assessment variables with 6-month DRS total score.

| Variable (n) | Spearman's correlation coefficient or Wilcoxon rank sum test |
| --- | --- |
| **EEG: Full Sample** |  |
| Age | 0.363 [0.058, 0.626]; p=0.015<sup>a</sup> |
| CRS-R Conscious<sup>b</sup><br>y (n=19); n (n=25) | -11 [-20, -2]; p=0.006<sup>a</sup> |
| Days to EEG | 0.100 [-0.195, 0.424]; p=0.517 |
| Active-motor [yes/no]<br>y (n=8); n (n=35) | 3 [-4, 11]; p=0.278 |
| Active-motor [% accuracy] | 0.143 [-0.15, 0.431]; p=0.359 |
| Passive-language [yes/no]<br>y (n=39); n (n=3)<sup>c</sup> | 1 [-24, 21]; p=0.640 |
| Passive-language [% accuracy] | -0.080 [-0.386, 0.276]; p=0.614 |
| **EEG: Coma, VS/UWS, MCS- Subsample** |  |
| Age | 0.369 [0.012, 0.665]; p=0.029 |
| CRS-R Conscious<sup>b</sup><br>y (n=10); n (n=25) | -9 [-19, 0]; p=0.067 |
| Days to EEG | 0.174 [-0.189, 0.496]; p=0.318 |
| Active-motor [yes/no]<br>yes (n=6); no (n=28) | 5 [-4, 18]; p=0.272 |
| Active-motor [% accuracy] | 0.126 [-0.245, 0.455]; p=0.478 |
| Passive-language [yes/no]<br>y (n=30); n (n=3)<sup>c</sup> | 2 [-19, 28]; p=0.569 |
| Passive-language [% accuracy] | -0.162 [-0.495, 0.214]; p=0.367 |
| **fMRI: Full Sample** |  |
| Age | 0.389 [0.028, 0.705]; p=0.031<sup>a</sup> |
| CRS-R Conscious<sup>b</sup><br>y (n=12); n (n=19) | -9.07 [-20, -2]; p=0.010<sup>a</sup> |
| Days to fMRI | 0.159 [-0.217, 0.528]; p=0.392 |
| Active-motor [yes/no]<br>y (n=8); n (n=22) | 2 [-7, 10]; p=0.494 |
| Active-motor [%suprathreshold] | 0.150 [-0.2, 0.459]; p=0.428 |
| Active-motor [ROI z-stat] | 0.125 [-0.21, 0.483]; p=0.511 |
| Passive-language [yes/no]<br>y (n=24); n (n=6) | -3.80 [-19, 5]; p=0.549 |
| Passive-language [%suprathreshold] | -0.334 [-0.634, 0.032]; p=0.071 |
| Passive-language [ROI z-stat] | -0.241 [-0.562, 0.137]; p=0.20 |
| Resting-state DMN [yes/no]<br>y (n=12); n (n=19) | -6 [-16, 1]; p=0.148 |
| Resting-state DMN [z-stat] | -0.332 [-0.624, 0.025]; p=0.068 |
| **fMRI: Coma, VS, MCS- Subsample** |  |
| age | 0.306 [-0.139, 0.687]; p=0.146 |
| CRS-R Conscious<sup>b</sup><br>y (n=5); n (n=19) | -9 [-20, 2]; p=0.124 |
| Days to fMRI | 0.186 [-0.212, 0.564]; p=0.384 |
| Active-motor [yes/no]<br>y (n=8); n (n=16) | 0 [-13, 9]; p=0.902 |
| Active-motor [% suprathreshold] | -0.001 [-0.401, 0.391]; p=0.995 |
| Active-motor [ROI z-stat] | 0.155 [-0.230, 0.524]; p=0.469 |
| Passive-language [yes/no] | 0 [-17, 8]; p=0.947 |
| y (n=18); n (n=6) |  |
| Passive-language [% suprathreshold] | -0.232 [-0.606, 0.187]; p=0.275 |
| Passive-language [ROI z-stat] | -0.109 [-0.512, 0.313]; p=0.611 |
| Resting-state DMN [yes/no] |  |
| y (n=10); n (n=14) | -9 [-20, 1]; p=0.093 |
| Resting-state DMN [z-stat] | -0.411 [-0.707, -0.027]; p=0.046<sup>a</sup> |
| <sup>a</sup> Statistically significant associations with 6-month DRS total score (p<0.05)<br><sup>b</sup> MCS-, MCS+, eMCS vs Coma, VS/UWS<br><sup>c</sup> Only 3 participants did not demonstrate a positive response to passive-language with EEG assessment, therefore, the results should be interpreted with caution<br>The CRS-R Conscious variable compares participants with a behavioral diagnosis of coma or VS/UWS versus those in MCS-, MCS+, or eMCS on the CRS-R<br>*Abbreviations: CI confidence interval; CM<sub>Command</sub> Composite Measure including any assessment indicating command-following; : CM<sub>Conscious</sub> Composite Measure including any assessment indicating conscious awareness; CRS-R Coma Recovery Scale-Revised; DMN default mode network; DRS disability rating scale; EEG electroencephalography, fMRI functional magnetic resonance imaging, MCS- minimally conscious state minus (without evidence of language function); mo months; n no; ROI region of interest; SD standard deviation; VS/UWS vegetative state/unresponsive wakefulness syndrome; y yes* |  |

#### Sharing results with families and clinicians

Results of active-motor-imagery and passive-language EEG and fMRI have diagnostic and prognostic clinical relevance, as evidenced by recent clinical guidelines^33,34^ that recommend these assessments for persons with DoC. For this reason, we elected to share the results of the active-motor-imagery and passive-language fMRI assessment with families and clinicians, if the surrogate expressed interest in receiving the information. All surrogates requested the fMRI results, which were shared within 24 hours of data acquisition. The goals of care (i.e., “code status”) for all patients was the same before and after results were shared. We did not share results of EEG assessments or of rs-fMRI DMN connectivity because when we began enrollment in 2018, we did not have an approach for returning these results within 24hrs of data acquisition (Supplementary Table 17) as we do now.^56^

### Discussion

In this prospective study of critically ill patients with acute severe TBI, we demonstrate that multiple features of the CRS-R behavioral assessment, including DoC diagnosis, evidence of consciousness, and evidence of command-following, are associated with less disability at 6 months post-TBI. Task-based EEG and fMRI evidence of command-following were also associated with outcome, but this relationship did not strengthen that of the CRS-R assessment alone. Individual active-motor-imagery and passive-language EEG and fMRI biomarkers, and resting-state fMRI biomarkers, were not associated with 6-month DRS total scores, the primary outcome measure, in the full sample. However, rs-fMRI DMN connectivity was associated with 6-month DRS total scores in a subset of patients who did not follow commands behaviorally, and with multiple secondary outcome measures in the full sample, though these analyses were not corrected for multiple comparisons. Collectively, our results indicate that even a single comprehensive, standardized behavioral assessment identifies patients with acute traumatic DoC who may make functional gains in their recovery. Advanced EEG and fMRI biomarkers may provide additional information about outcome but require further study in large multi-center cohorts.

The potential value of leveraging advanced electrophysiologic and neuroimaging techniques for acute DoC prognosis has been demonstrated in multiple studies.^13,14,29,30,31,57–60^ However, these studies were conducted in samples with mixed traumatic and non-traumatic etiologies and routinely find that, compared to patients with non-TBI, patients with TBI have better outcomes. Moreover, large observational studies published over the past 5 years suggest that recovery of consciousness and in-home independence after acute severe TBI is not just possible, but common.^42,61,62^ Thus, while multiple studies, including this one, have detected a high rate of CMD on task-based EEG and fMRI assessment of patients with DoC^12,13,19^, standardized behavioral assessment may be the most accurate, and feasible, tool for TBI prognostication.

Published guidelines recommend that patients with DoC be assessed with standardized behavioral measures as well as task-based EEG and fMRI to establish diagnosis.^33,34^ We evaluated two composite measures as predictors in our prognostic models, CM_Command_ and CM_Conscious_, because recovery of command-following and consciousness have each been associated with better outcome after TBI,^3,5,6,63^ Of note, the composition of the CM_Conscious_ group and the CM_Command_ group is the same except that participants with a CRS-R diagnosis of MCS-, but no evidence of CMD (n=6), are in the CM_Conscious_ positive group but not the CM_Command_ positive group. Our observation that CM_Conscious_ was more consistently associated with outcome than CM_Command_ suggests that behavioral milestones other than command-following, which is frequently used in TBI prognostic models, may more accurately predict recovery.^63^ Command-following requires the integration of multiple cognitive processes (e.g., language comprehension, vigilance) and emerges after recovery of consciousness.^64^ Novel task- and stimulus-based paradigms are needed to determine whether advanced EEG and fMRI can also be used to detect lower levels of consciousness.^22^

In prior studies, the presence of responses to active-motor-imagery tasks^13,14^ and passive-language stimuli,^24,57,65,66^ as well as intact DMN connectivity,^31^ were associated with better functional outcome. We included additional variables in our analyses that represent not only the presence, but the magnitude of these phenomena. For example, we investigated whether a response to active-motor-imagery fMRI (i.e., dichotomous present versus absent determination) as well as the magnitude and extent of the response (continuous z-statistic and proportion) were associated with outcome. We found that while the dichotomous determination of presence or absence of responses to active-motor-imagery tasks and passive-language stimuli, and intact DMN connectivity, were not associated with outcome, a greater response to passive-language fMRI and stronger DMN connectivity may be associated with better 6-month functional outcome.

Nearly one in four participants (10/45) died due to WLST in the ICU, all of whom had a behavioral diagnosis of coma, VS/UWS, or MCS-. CMD was present in 3/10 of these participants. The absence of an association between CMD and outcome in our data may reflect our observation that only five participants who responded to the task-based EEG or fMRI paradigm survived to 6-months. Furthermore, most surviving participants had moderate, minimal, or no disability at 6-months. Given that only participants with an acute behavioral diagnosis of coma, VS/UWS, or MCS-died (all from WLST), the relationship between behavioral level of consciousness and outcome may also reflect the high rate of WLST. Patients who appear unconscious at the bedside are more likely to receive a poor prognosis, leading to WLST and the observed association between CRS-R and outcome. This phenomenon is often described as a self-fulfilling prophecy.^67,68^ Despite growing evidence that recovery is possible after severe TBI,^62,42,61^ WLST remains the leading cause of death in hospitalized patients with TBI,^62,69,70^ highlighting the importance of developing multimodal tools to improve outcome prediction for patients who have not recovered consciousness^60^ and statistical approaches for imputing outcomes for patients who die due to WLST.^62^

There is no single optimal measure for assessment of outcomes after severe TBI. For this reason, we tested the association between 6-month outcome and multiple secondary outcome measures. Consistent with the DRS total score results, the presence of responses to active-motor-imagery and passive-language EEG or fMRI were not associated with secondary outcomes. The CM_Conscious_ composite measure, passive-language fMRI, and rs-fMRI measures of DMN connectivity were associated with most of the secondary outcomes in the full sample of participants. Thus, the results of the secondary outcomes did not identify additional EEG or fMRI biomarkers associated with 6-month outcome but suggest that DRS_Depend_ and GOSE-TBI should be considered in future studies of DoC recovery.

Our study includes a larger sample of patients with severe TBI compared to most recent studies focused on acute DoC prognostication.^14,29–31^ However, our high rate of WLST, inclusion of participants who were behaviorally following commands (and therefore more likely to have a good outcome regardless of EEG or fMRI findings), and restriction of enrollment to patients with TBI may have limited our ability to detect associations between some biomarkers and outcome. In one of the first studies to establish the predictive validity of CMD in patients with acute DoC, the presence of CMD, assessed using similar EEG methods to those employed in our study, was associated with approximately the same odds of recovery as an etiology of TBI.^14^ A separate acute DoC prognostication study found that an etiology of TBI and level of consciousnesses were as, or more, predictive of 3- and 12-month^30^ outcome compared to EEG and rs-fMRI measures. These findings may explain the absence of an association between CMD and outcome in our TBI cohort, as the higher likelihood of functional recovery in patients with acute severe TBI, as compared to patients with non-traumatic etiologies of acute DoC, may make a CMD diagnosis less prognostically relevant.

### Conclusions

Establishing prognosis after severe TBI is a critical component of clinical management as it directly informs decisions about WLST and access to intensive rehabilitation. Current prediction models based on demographic information, visual evaluation of structural imaging and resting-state EEG, and bedside neurological examination are imprecise; they do not include standardized measures of consciousness or comprehensively assess brain function. We showed that a single comprehensive standardized behavioral assessment of consciousness is strongly associated with outcome 6 months after severe TBI. Serial behavioral assessment may further optimize prognostication^60^ ^71^ and reduce premature WLST.^62^ While it may be infeasible to administer the full-length CRS-R serially, an abbreviated version of the CRS-R was recently developed and validated specifically for detecting consciousness (i.e., MCS-) in the ICU.^72^ EEG and fMRI measures of brain function, while promising, require further investigation before they are implemented in prognostic models for severe TBI.^56^

## Data Availability

All data produced in the present study are available upon reasonable request to the authors

https://www.dropbox.com/scl/fi/8arsxv672lk0hob3w4rhw/RESPONSE_fmri_figures_20240927.html?rlkey=mqyu0l4c77usu2azydidj145u&dl=0

## Abbreviations

CAP: Confusion Assessment Protocol
CCP: covert cortical processing
CMD: cognitive motor dissociation
CRS-R: Coma Recovery Scale-Revised
DMN: default mode network
DoC: disorders of consciousness
DRS: Disability Rating Scale
EEG: electroencephalography
fMRI: functional MRI
GOSE: Glasgow Outcome Scale-Extended
ICU: intensive care unit
LoC: level of consciousness
MCS-: minimally conscious state without language
MCS+: minimally conscious state with language
PTCS: post-traumatic confusional state
ROI: region of interest
TBI: traumatic brain injury
VS/UWS: vegetative state/unresponsive wakefulness syndrome

## Acknowledgements

The authors thank the nursing staffs of the Massachusetts General Hospital Neurosciences ICU, Multidisciplinary ICU, and the Surgical ICU. We also thank Joseph Cohen and the EEG technologists, as well as Karen Rich, Chrissy Foster, Kellie Cahill and the MRI technologists for their assistance with data acquisition. We are grateful to the patients and families in this study for their participation and support.

## Data Availability Statement

The data that support the findings of this study are available from the corresponding author, upon reasonable request. Individual participant and group-level fMRI data visualization is available <u>here</u>.

## Funding

Dr. Bodien: National Institutes of Health

Dr. Fecchio: Massachusetts General Hospital Transformative Scholar Award

Dr. Kirsch: National Institutes of Health

Dr. Healy: National Institutes of Health

Dr. Edlow: National Institutes of Health Director’s Office (DP2HD101400), National Institute of Neurological Disorders and Stroke (R21NS109627, R01NS138257), Chen Institute MGH Research Scholar Award, and MIT/MGH Brain Arousal State Control Innovation Center (BASCIC) project

## Competing Interest

The authors report no competing interests.

## Notes

### Competing Interest Statement

The authors have declared no competing interest.

### Clinical Protocols

https://clinicaltrials.gov/study/NCT03504709?cond=Consciousness%20Disorders&term=Edlow,%20Brian&rank=6

### Funding Statement

This study was funded by National Institutes of Health, National Institutes of Health Directors Office (DP2HD101400), National Institute of Neurological Disorders and Stroke (R21NS109627, R01NS138257), Chen Institute MGH Research Scholar Award, and MIT/MGH Brain Arousal State Control Innovation Center (BASCIC) project.

### Author Declarations

IRB of Massachusetts General Brigham gave ethical approval for this work

## References

1. Steyerberg EW, Mushkudiani N, Perel P, et al. Predicting outcome after traumatic brain injury: development and international validation of prognostic scores based on admission characteristics. PLoS medicine. 2008;5(8):e165.

2. Hammond FM, Katta-Charles S, Russell MB, et al. Research needs for prognostic modeling and trajectory analysis in patients with disorders of consciousness. Neurocritical Care. 2021;35(1):55–67.

3. Giacino JT, Kalmar KT. The vegetative and minimally conscious states: a comparison of clinical features and functional outcome. Journal of Head Trauma Rehabilitation. 1997;12(4):36–51.

4. Katz DI, Polyak M, Coughlan D, Nichols M, Roche A. Natural history of recovery from brain injury after prolonged disorders of consciousness: outcome of patients admitted to inpatient rehabilitation with 1-4 year follow-up. Prog Brain Res. 2009;177:73–88.

5. Giacino JT, Sherer M, Christoforou A, et al. Behavioral recovery and early decision making in patients with prolonged disturbance in consciousness after traumatic brain injury. Journal of Neurotrauma. 2020;37(2):357–365.

6. Thibaut A, Bodien YG, Laureys S, Giacino JT. Minimally conscious state “plus”: diagnostic criteria and relation to functional recovery. Journal of Neurology. 2020;267(5):1245–1254.

7. Edlow BL, Olchanyi M, Freeman HJ, et al. Multimodal MRI reveals brainstem connections that sustain wakefulness in human consciousness. Sci Transl Med. 2024;16(745):eadj4303.

8. Bodien YG, Katz D, Schiff N, Giacino JT. Behavioral assessment of patients with disorders of consciousness. Semin Neurol. 2022;42(3):249–258.

9. Childs NL, Mercer WN, Childs HW. Accuracy of diagnosis of persistent vegetative state. Neurology. 1993;43(8):1465–1467.

10. Andrews K, Murphy L, Munday R, Littlewood C. Misdiagnosis of the vegetative state: retrospective study in a rehabilitation unit. BMJ. 1996;313(7048):13–16.

11. Schnakers C, Vanhaudenhuyse A, Giacino J, et al. Diagnostic accuracy of the vegetative and minimally conscious state: clinical consensus versus standardized neurobehavioral assessment. BMC Neurol. 2009;9:35.

12. Edlow BL, Chatelle C, Spencer CA, et al. Early detection of consciousness in patients with acute severe traumatic brain injury. Brain. 2017;140(9):2399–2414.

13. Claassen J, Doyle K, Matory A, et al. Detection of brain activation in unresponsive patients with acute brain injury. N Engl J Med. 2019;380(26):2497–2505.

14. Egbebike J, Shen Q, Doyle K, et al. Cognitive-motor dissociation and time to functional recovery in patients with acute brain injury in the USA: a prospective observational cohort study. Lancet Neurol. 2022;21(8):704–713.

15. Owen AM, Coleman MR, Boly M, Davis MH, Laureys S, Pickard JD. Detecting awareness in the vegetative state. Science. 2006;313(5792):1402.

16. Monti MM, Vanhaudenhuyse A, Coleman MR, et al. Willful modulation of brain activity in disorders of consciousness. N Engl J Med. 2010;362(7):579–589.

17. Kondziella D, Friberg CK, Frokjaer VG, Fabricius M, Møller K. Preserved consciousness in vegetative and minimal conscious states: systematic review and meta-analysis. *Journal of Neurology*, Neurosurgery & Psychiatry. 2016;87(5):485–492.

18. Schnakers C, Hirsch M, Noé E, et al. Covert cognition in disorders of consciousness: a meta-analysis. Brain Sci. 2020;10(12).

19. Bodien YG, Allanson J, Cardone P, et al. Cognitive motor dissociation in disorders of consciousness. N Engl J Med. 2024;391(7):598–608.

20. Schiff ND. Cognitive motor dissociation following severe brain injuries. JAMA Neurol. 2015;72(12):1413–1415.

21. Edlow BL, Claassen J, Schiff ND, Greer DM. Recovery from disorders of consciousness: mechanisms, prognosis and emerging therapies. Nat Rev Neurol. 2021;17(3):135–156.

22. Young MJ, Fecchio M, Bodien YG, Edlow BL. Covert cortical processing: a diagnosis in search of a definition. Neurosci Conscious. 2024;2024(1):niad026.

23. Coleman MR, Davis MH, Rodd JM, et al. Towards the routine use of brain imaging to aid the clinical diagnosis of disorders of consciousness. Brain. 2009;132(Pt 9):2541–2552.

24. Sokoliuk R, Degano G, Banellis L, et al. Covert Speech Comprehension Predicts Recovery From Acute Unresponsive States. Ann Neurol. 2021;89(4):646–656.

25. Vanhaudenhuyse A, Noirhomme Q, Tshibanda LJ, et al. Default network connectivity reflects the level of consciousness in non-communicative brain-damaged patients. Brain. 2010;133(Pt 1):161–171.

26. Demertzi A, Tagliazucchi E, Dehaene S, et al. Human consciousness is supported by dynamic complex patterns of brain signal coordination. Science Advances. 2019;5(2):eaat7603.

27. Norton L, Kazazian K, Gofton T, et al. Functional Neuroimaging as an Assessment Tool in Critically Ill Patients. Annals of Neurology. 2023;93(1):131–141.

28. Threlkeld ZD, Bodien YG, Rosenthal ES, et al. Functional networks reemerge during recovery of consciousness after acute severe traumatic brain injury. Cortex. 2018;106:299–308.

29. Amiri M, Fisher PM, Raimondo F, et al. Multimodal prediction of residual consciousness in the intensive care unit: the CONNECT-ME study. Brain. 2023;146(1):50–64.

30. Amiri M, Raimondo F, Fisher PM, et al. Multimodal Prediction of 3- and 12-Month Outcomes in ICU Patients with Acute Disorders of Consciousness. Neurocrit Care. 2024;40(2):718–733.

31. Rohaut B, Calligaris C, Hermann B, et al. Multimodal assessment improves neuroprognosis performance in clinically unresponsive critical-care patients with brain injury. Nat Med. 2024.

32. Rappaport M, Hall KM, Hopkins K, Belleza T, Cope DN. Disability rating scale for severe head trauma: coma to community. Arch Phys Med Rehabil. 1982;63(3):118–123.

33. Giacino JT, Katz DI, Schiff ND, et al. Practice Guideline Update Recommendations Summary: Disorders of Consciousness: Report of the Guideline Development, Dissemination, and Implementation Subcommittee of the American Academy of Neurology; the American Congress of Rehabilitation Medicine; and the National Institute on Disability, Independent Living, and Rehabilitation Research. Arch Phys Med Rehabil. 2018;99(9):1699–1709.

34. Kondziella D, Bender A, Diserens K, et al. European Academy of Neurology guideline on the diagnosis of coma and other disorders of consciousness. European Journal of Neurology. 2020;27(5):741–756.

35. Bodien Y, LaRovere K, Kondziella D, Taran S, Estraneo A, Shutter L. . Common data elements for disorders of consciousness: Recommendations from the Working Group on Outcomes and Endpoints. Neurocritical Care. 2024;41(2):357–368.

36. Teasdale G, Jennett B. Assessment of coma and impaired consciousness. A practical scale. Lancet. 1974;2(7872):81–84.

37. Giacino JT, Kalmar K. Diagnostic and prognostic guidelines for the vegetative and minimally conscious states. Neuropsychol Rehabil. 2005;15(3-4):166–174.

38. Laureys S, Celesia GG, Cohadon F, et al. Unresponsive wakefulness syndrome: a new name for the vegetative state or apallic syndrome. BMC Medicine. 2010;8:68.

39. Giacino J, Ashwal S, Childs N, et al. The minimally conscious state: definition and diagnostic criteria. Neurology. 2002;58(3):349–353.

40. Giacino JT, Kalmar K, Whyte J. The JFK Coma Recovery Scale-Revised: measurement characteristics and diagnostic utility. Arch Phys Med Rehabil. 2004;85(12):2020–2029.

41. Giacino JT, Whyte J, Bagiella E, et al. Placebo-controlled trial of amantadine for severe traumatic brain injury. N Engl J Med. 2012;366(9):819–826.

42. McCrea MA, Giacino JT, Barber J, et al. Functional outcomes over the first year after moderate to severe traumatic brain injury in the prospective, longitudinal TRACK-TBI study. JAMA Neurol. 2021;78(8):982–992.

43. Hicks R, Giacino J, Harrison-Felix C, Manley G, Valadka A, Wilde EA. Progress in developing common data elements for traumatic brain injury research: version two--the end of the beginning. J Neurotrauma. 2013;30(22):1852–1861.

44. Wilson JT, Pettigrew LE, Teasdale GM. Structured interviews for the Glasgow Outcome Scale and the extended Glasgow Outcome Scale: guidelines for their use. J Neurotrauma. 1998;15(8):573–585.

45. Snider SB, Kowalski RG, Hammond FM, et al. Comparison of common outcome measures for assessing independence in patients diagnosed with disorders of consciousness: a Traumatic Brain Injury Model Systems study. J Neurotrauma. 2022;39(17-18):1222–1230.

46. TRACK-TBI Resources for Researchers. https://tracktbi.ucsf.edu/researchers. Published 2024. Accessed August 26, 2024.

47. Delorme A, Makeig S. EEGLAB: an open source toolbox for analysis of single-trial EEG dynamics including independent component analysis. J Neurosci Methods. 2004;134(1):9–21.

48. Noirhomme Q, Lesenfants D, Gomez F, et al. Biased binomial assessment of cross-validated estimation of classification accuracies illustrated in diagnosis predictions. Neuroimage Clin. 2014;4:687–694.

49. van der Kouwe AJ, Benner T, Salat DH, Fischl B. Brain morphometry with multiecho MPRAGE. Neuroimage. 2008;40(2):559–569.

50. Whitfield-Gabrieli S, Nieto-Castanon A. Conn: a functional connectivity toolbox for correlated and anticorrelated brain networks. Brain Connect. 2012;2(3):125–141.

51. Nieto-Castanon A. Handbook of functional connectivity Magnetic Resonance Imaging methods in CONN. 2020.

52. Gibbons J. Nonparametric Statistical Methods. Technometrics. 2012;16:477–478.

53. Rubin ML, Yamal JM, Chan W, Robertson CS. Prognosis of six-month Glasgow Outcome Scale in severe traumatic brain injury using hospital admission characteristics, injury severity characteristics, and physiological monitoring during the first day post-injury. J Neurotrauma. 2019;36(16):2417–2422.

54. Faugeras F, Rohaut B, Valente M, et al. Survival and consciousness recovery are better in the minimally conscious state than in the vegetative state. Brain Inj. 2018;32(1):72–77.

55. Foster ED, Deardorff A. Open Science Framework (OSF). J Med Libr Assoc. 2017;105(2):203–206.

56. Bodien YG, Fecchio, M., Freeman, H. J., Sanders, W. R., Meydan, A., Lawrence, P., Kirsch, J., Fischer D., Cohen J., Rubin E., He J., Schaefer P. W., Hochberg L. R., Rapalino O., Cash S., Young M., Edlow, B. L. Clinical Implementation of Functional MRI and EEG to Detect Cognitive Motor Dissociation: Lessons Learned in an Acute Care Hospital. Neurology: Clinical Practice. 2025;15:1.

57. Dhakal K, Rosenthal ES, Kulpanowski AM, et al. Increased task-relevant fMRI responsiveness in comatose cardiac arrest patients is associated with improved neurologic outcomes. J Cereb Blood Flow Metab. 2024;44(1):50–65.

58. Wagner F, Hänggi M, Weck A, Pastore-Wapp M, Wiest R, Kiefer C. Outcome prediction with resting-state functional connectivity after cardiac arrest. Scientific Reports. 2020;10(1):11695.

59. Comanducci A, Boly M, Claassen J, et al. Clinical and advanced neurophysiology in the prognostic and diagnostic evaluation of disorders of consciousness: review of an IFCN-endorsed expert group. Clin Neurophysiol. 2020;131(11):2736–2765.

60. Fischer D, Edlow BL. Coma prognostication after acute brain injury: A review. JAMA Neurol. 2024.

61. Kowalski RG, Hammond FM, Weintraub AH, et al. Recovery of consciousness and functional outcome in moderate and severe traumatic brain injury. JAMA Neurol. 2021;78(5):548–557.

62. Sanders WR, Barber JK, Temkin NR, et al. Recovery potential in patients who died after withdrawal of life-sustaining treatment: A TRACK-TBI propensity score analysis. J Neurotrauma. 2024;doi: 10.1089/neu.2024.0014.

63. Snider SB, Deng H, Hammond FM, et al. Quantifying the relationship between time to command-following and outcomes after TBI: the “1%” rule. medRxiv. 2024:2024.2006.2004.24308423.

64. Martens G, Bodien Y, Sheau K, Christoforou A, Giacino JT. Which behaviours are first to emerge during recovery of consciousness after severe brain injury? Ann Phys Rehabil Med. 2020;63(4):263–269.

65. Perrin F, Schnakers C, Schabus M, et al. Brain response to one’s own name in vegetative state, minimally conscious state, and locked-in syndrome. Arch Neurol. 2006;63(4):562–569.

66. Di HB, Yu SM, Weng XC, et al. Cerebral response to patient’s own name in the vegetative and minimally conscious states. Neurology. 2007;68(12):895–899.

67. Izzy S, Compton R, Carandang R, Hall W, Muehlschlegel S. Self-fulfilling prophecies through withdrawal of care: do they exist in traumatic brain injury, too? Neurocrit Care. 2013;19(3):347–363.

68. Graham M. Burying our mistakes: Dealing with prognostic uncertainty after severe brain injury. Bioethics. 2020;34(6):612–619.

69. Turgeon AF, Lauzier F, Simard JF, et al. Mortality associated with withdrawal of life-sustaining therapy for patients with severe traumatic brain injury: a Canadian multicentre cohort study. CMAJ. 2011;183(14):1581–1588.

70. van Veen E, van der Jagt M, Citerio G, et al. Occurrence and timing of withdrawal of life-sustaining measures in traumatic brain injury patients: a CENTER-TBI study. Intensive Care Medicine. 2021;47(10):1115–1129.

71. Wannez S, Heine L, Thonnard M, Gosseries O, Laureys S, Group CS. The repetition of behavioral assessments in diagnosis of disorders of consciousness. Ann Neurol. 2017;81(6):883–889.

72. Bodien YG, Vora I, Barra A, et al. Feasibility and validity of the Coma Recovery Scale-Revised for Accelerated Standardized Testing: A practical assessment tool for detecting consciousness in the intensive care unit. Ann Neurol. 2023;94(5):919–924.

