## Supplementary Materials for "Acute biomarkers of consciousness are associated with recovery after severe traumatic brain injury"

Yelena G. Bodien*,^1,2,3^ Matteo Fecchio*,^1,2^ Natalie Gilmore,^1,2^ Holly J. Freeman,^1,2^ William R. Sanders,^1,2^ Anogue Meydan,^1,2^ Phoebe K. Lawrence,^1,2^ Alexander S. Atalay,^1,2^ John Kirsch,^4,5^ Brian C. Healy,^2,6,7^ Brian L. Edlow^1,2,4^

1. Center for Neurotechnology and Neurorecovery, Massachusetts General Hospital and Harvard Medical School, Boston, MA, 02114

2. Department of Neurology, Harvard Medical School, Boston, MA, 02114

3. Department of Physical Medicine and Rehabilitation, Spaulding Rehabilitation Hospital, Harvard Medical School, Charlestown, MA, 02129

4. Athinoula A. Martinos Center for Biomedical Imaging, Massachusetts General Hospital, Charlestown, MA, 02129

5. Department of Radiology, Massachusetts General Hospital, Harvard Medical School, Boston, MA 02114

6. Ann Romney Center for Neurologic Diseases, Brigham and Women’s Hospital, Boston, MA, 02115

7. Massachusetts General Hospital Biostatistics Center, Massachusetts General Hospital, Boston, MA, 02114

*co-first authors

Correspondence to:

Yelena G. Bodien, PhD

Center for Neurotechnology and Neurorecovery

Massachusetts General Hospital

101 Merrimac Street – Suite 310, Boston, MA 02114, USA

Web: www.ComaRecoveryLab.org

| **Table of Contents** | **Page** |
| --- | --- |
| Supplementary Methods | 3 |
| Supplementary Results | 11 |
| Supplementary Table 1: Coma Recovery Scale- Revised Test Completion Codes | 12 |
| Supplementary Table 2: Instructions administered prior to and during EEG and functional MRI | 13 |
| Supplementary Table 3: Regions of interest for stimulus-based functional MRI analysis | 14 |
| Supplementary Table 4: Patient demographics and clinical characteristics | 15 |
| Supplementary Table 5: Details of CRS-R Behavioral Assessments | 16 |
| Supplementary Table 6: Sedative, anxiolytic and analgesic medications administered during functional MRI and EEG | 20 |
| Supplementary Table 7: Drains, Lines, Tubes, and Monitors at time of functional MRI and EEG | 24 |
| Supplementary Table 8: Healthy participant responses to functional MRI and EEG assessments | 27 |
| Supplementary Table 9: Results of EEG and fMRI tests for participants who were not followed at 6-months | 29 |
| Supplementary Table 10: Secondary Outcomes for Individual Participants | 30 |
| Supplementary Table 11: Association between CRS-R Diagnosis and 6mo Outcome | 33 |
| Supplementary Table 12 Association between Composite Measures of Command-following and Consciousness and 6mo Outcome | 34 |
| Supplementary Table 13 Association between EEG Biomarkers and 6mo Outcome | 36 |
| Supplementary Table 14 Association between fMRI Biomarkers and 6mo Outcome | 38 |
| Supplementary Table 15: Association between EEG Biomarkers and 6mo Outcome in Subsample of Participants with Coma, VS/UWS, and MCS- | 41 |
| Supplementary Table 16: Association between fMRI Biomarkers and 6mo Outcome in Subsample of Participants with Coma, VS/UWS, and MCS- | 43 |
| Supplementary Table 17: Clinical interventions, data sharing, and goals of care | 46 |
| Supplementary Figure 1: Distribution of CRS-R scores | 51 |
| Supplementary Figure 2: Distribution of DRS scores | 52 |
| Supplementary Figure 3: DRS Outcome Across Behavioral Diagnosis and Composite Measures of Command-following and Consciousness in Participants who Survived | 53 |
| Supplementary Figure 4: DRS Outcome Across Responses to EEG and fMRI in Participants who Survived | 54 |
| Supplementary Figure 5: DRS Outcome Across Responses to EEG and fMRI in Participants with a CRS-R Diagnosis Coma, VS/UWS, MCS- Subsample | 55 |
| References | 56 |

**Supplementary Methods**

*Measures*

Coma Recovery Scale-Revised (CRS-R)

The CRS-R^1^ is a standardized, hierarchically organized, behavioral measure that was designed to monitor recovery in persons with disorders of consciousness (DoC). The scale assesses behaviors across six domains: auditory function, visual function, motor function, oromotor/verbal function, communication, and arousal level. The CRS-R provides a total score (0-23) as well as behavioral determinations of VS/UWS, MCS without language function (MCS-; i.e., at least one of the following: visual fixation, visual pursuit, object localization, localization to noxious stimulation, object manipulation, automatic motor responses, or non-functional communication, but no command-following, object recognition, or intelligible verbalization^2-4^), MCS with language function (MCS+; i.e., at least one of the following: command-following, object recognition, or intelligible verbalization^2-4^) or emerged from MCS (eMCS, also known as the post-traumatic confusional state [PTCS]^5^). Participants who had no evidence of arousal on the CRS-R assessment and no behaviors consistent with MCS were classified with a coma diagnosis.

The examiner assigned each CRS-R rating a Test Completion Code (Supplementary Table 1) to document whether the assessment was valid, and if not, the rationale for this determination. When CRS-R subscales were not valid, we supplemented the information from the valid CRS-R subscales with clinical notes to derive a CRS-R diagnosis (See Supplementary Materials). For example, if the valid CRS-R subscales indicated a diagnosis of VS/UWS and none of the clinical team notes indicated any signs of MCS, a diagnosis of VS/UWS was assigned.

Confusion Assessment Protocol (CAP)

The CAP^6^ is a composite measure comprised of seven subscales that assess cognitive impairment, disorientation, restlessness, nighttime sleep disturbance, daytime somnolence, psychotic-type symptoms. and symptom fluctuation. Cognitive impairment items assess basic attention, language comprehension, and memory. Disorientation is assessed via the Galveston Orientation and Amnesia Test. Restlessness (determined via the Agitated Behavior Scale) and the remaining items are rated by a clinician or other observer. Evidence of four symptoms of confusion, or three if disorientation is among the symptoms, indicates PTCS.

Disability Rating Scale (DRS)

The DRS^7^ evaluates recovery from coma to return to employment in persons with TBI by combining the three Glasgow Coma Scale items with three items assessing cognitive abilities related to performing self-care activities, and two items assessing level of function and employability. DRS total scores range from 0 (no disability) to 30 (death); a score of <12 has been associated with recovery of at least partial independence. DRS_Depend_^8^ is a dichotomized derivative of the DRS that identifies individuals who are dependent (i.e., Level of function ≥4, and at least some assistance needed [score >0] on the Verbal, Feeding, Toileting, or Grooming items).

Glasgow Outcome Scale-Extended (GOSE)

The GOSE^9^ categorizes TBI outcome into eight categories: 1) death, 2) vegetative state (e.g., unresponsive), 3) lower severe disability (e.g., cannot be home alone >8 hours due to level of dependency for daily activities), 4) upper severe disability (e.g., can be home alone >8 hours but may be dependent for some daily activities), 5) lower moderate disability (e.g., unable to return to work), 6) upper moderate disability (e.g., able to return to work with accommodations), 7) lower good recovery (e.g., return to normal life despite minor deficits), and 8) upper good recovery (e.g., return to normal life without deficits). The GOSE is completed via a structured interview by participants or their surrogates. After each question is asked, a total score is determined by identifying the item indicating the lowest level of function. The GOSE can also be dichotomized into “unfavorable outcome” (i.e., GOSE ≤3) and “favorable outcome” (i.e., GOSE ≥4), although a standard approach to determining this cut-point is not available.

*EEG Data Acquisition*

During the EEG assessment, participants were instructed to imagine opening and closing their right hand (i.e., active-motor-imagery paradigm) and listen to a story (i.e., passive-language paradigm). The active-motor-imagery paradigm started with instructions for the task (see Supplementary Table 2). Then, the command to imagine opening and closing the right hand was presented in alternating 10-sec blocks of stimulus (i.e., “ON”) and rest (i.e., “OFF”) and repeated 8 times, for a total task run time of 3min 30secs. The passive-language paradigm also started with instructions for the task. Then, an audio recording of the Alice in Wonderland book was read for approximately 2min 30secs, followed by 30secs of rest. This was repeated one time for a total task run time of 6min 30secs. Three runs of the active-motor-imagery paradigm and three runs of the passive-language paradigm were presented in a pseudorandomized order to ensure that each paradigm was not presented three times in a row. After every two runs there was a 4-min rest period. A 4-min rest period, followed by administration of the Arousal Facilitation Protocol,^1,10^ and another 4mins of rest preceded the first run and followed the last run. Auditory stimuli were administered via earphones (3M™ E-A-RTONE™ 3A) connected to an Artcessories HeadAmp box.

We acquired EEG data with a 19-electrode clinical XLTEK EEG system (Natus Medical Inc.; Pleasanton, CA) at a 256-, 512-, or 2048 Hz sampling rate and analyzed data using EEGlab^11^ and customized MATLAB (vR2016b) code. We concatenated runs for each paradigm to avoid generating spurious results related to the analysis process (i.e., due to different channel and independent component analysis [ICA] component selection). We divided each passive-language run into two sections of equal duration and concatenated each section with 2mins (i.e., half) of the rest period recorded prior to or after the run itself. To proceed with data analysis, we required that a participant have at least two runs of complete data.

*EEG Preprocessing*

EEG recordings were high-pass filtered (third-order Butterworth, zero-phase shift digital filter, 1 Hz), 1-sec epoched, and visually inspected for channel and epoch selection. Data were re-referenced to the average and artifact rejection was performed with EEGlab using ICA by study staff with expertise in EEG data analysis (M.F.) who was blinded to the behavioral diagnosis. Data were low-pass filtered at 30 Hz (third-order Butterworth, zero-phase shift digital filter), down-sampled to 256 Hz (if acquired at a higher frequency), and re-referenced using the Hjorth Laplacian transform.^12^

*fMRI Data Acquisition*

The fMRI assessment began with 10mins of rs-fMRI, during which participants were asked to stay awake with their eyes open (see Supplementary Table 2). Next, we administered two runs of the active-motor-imagery paradigm, and two runs of the passive-language paradigm. The active-motor-imagery paradigm began with instructions for the task. Then, the command to imagine (or stop imagining) opening and closing the right hand^13^ was presented in four alternating 17.5-sec blocks of stimulus and rest (i.e., scanner noise only) for a run time of 2min and 20secs. The active-motor paradigm was presented in two consecutive runs for a total paradigm time of 4min and40 sec. The passive-language paradigm also began with instructions. Then, an audio recording of the Alice in Wonderland book was read in four alternating 17.5-sec blocks of stimulus and rest (i.e., scanner noise only) for a total run time of 2min and 20secs. The passive-language paradigm was also presented in two consecutive runs for a total paradigm time of 4min and 40sec. Prior to each of the four runs, 19.5secs of data were acquired to obtain a stable baseline blood-oxygen level dependent (BOLD) signal. These data were excluded from analysis. An additional 2.5secs of data were acquired to optimize spatial co-registration and were, subsequently, excluded from further analysis.

MRI data were acquired with a 32-channel head coil on a 3-Tesla Skyra MRI scanner (Siemens Healthineers; Erlangen, Germany) located in the Massachusetts General Hospital Neurosciences ICU. Auditory stimuli were presented via MRI-compatible earphones (Newmatic Medical; Caledonia, MI) connected to the scanner’s sound system. The parameters of the BOLD fMRI sequence were: echo time (TE)=30ms, repetition time (TR)=1250ms, in-plane resolution=2.0x2.0mm, slice thickness=2 mm, interslice gap=0mm, matrix=106x105, field of view=212x212mm^2^, 72 slices, simultaneous multislice (SMS) factor=4. The rs-fMRI sequence was the same as that used for task/stimulus-based fMRI, except that it was 10mins 22secs long (10mins of analyzed data). High-spatial resolution 3D T1-weighted multi-echo magnetization prepared gradient echo (MEMPRAGE) anatomical images were acquired for registration purposes, as previously described.^14^

**Active-motor-imagery and passive-language fMRI region of interest analyses**

The fMRI analysis pipeline was described in a prior publication and is summarize in the the Supplementary Materials. In a first-level analysis of the individual runs, fMRI data processing was performed using FSL 5.0.7 (FMRIB's Software Library, www.fmrib.ox.ac.uk/fsl). Active-motor-imagery and passive-language blocks were contrasted against rest blocks. Z-statistic images were thresholded (Z>3.1) and a corrected cluster significance threshold of p=0.05 was used. The statistical threshold for cluster significance (Z>3.1) was selected to decrease false positive cluster activations.^54^ Higher-level analysis was carried out using a fixed effects model (FLAME in FSL).^55^^,^^56^ We selected regions of interest (ROIs) *a priori* based on fMRI studies of motor imagery (bilateral supplementary motor areas [SMA], premotor cortices [PMC], Supplementary Table 3), and language (bilateral superior temporal gyri [STG]) in patients with traumatic DoC and healthy participants.^17^ We used FEATQuery in FSL to quantify the percentage of voxels activated within each task and stimulus-specific ROI as well as the average Z-score in the ROI. We defined a positive response by the criterion that >0% of ROI voxels met the aforementioned statistical threshold.

*Task- and Stimulus-based fMRI Preprocessing and ROI Identification*

Active-motor-imagery and passive-language fMRI preprocessing steps were conducted in FMRI Expert Analysis Tool (FEAT) version 6.00 in FSL 5.0.7 (FMRIB's Software Library, www.fmrib.ox.ac.uk/fsl) and included motion correction, spatial smoothing, grand-mean intensity normalization of the entire 4D dataset by a single multiplicative factor, and high pass temporal filtering.^15^ Rs-fMRI data were analyzed using CONN^16^, and preprocessing steps included realignment with correction of susceptibility distortion interactions, slice timing correction, outlier detection, direct segmentation and MNI-space normalization, and smoothing. fMRI data were analyzed by trained study staff who had no knowledge of participants’ behavioral diagnosis.

We selected regions of interest (ROIs) *a priori* based on fMRI studies of motor imagery (bilateral supplementary motor areas [SMA], premotor cortices [PMC], Supplementary Table 3), and language (bilateral superior temporal gyrus [STG]) in patients with traumatic DoC and healthy participants.^17^ For the language stimuli, we used the bilateral superior temporal gyrus (STG) ROI distributed by the Harvard-Oxford Cortical Structural Atlas.^18^ For the motor imagery task, the bilateral supplementary motor areas (SMA) from the Harvard-Oxford Cortical Structural Atlas and premotor cortices (PMC) from the Juelich Histological Atlas ^19^ were combined as a single ROI (see Supplementary Table 3). All ROIs were transformed from standard atlas space into patient-native fMRI space for analysis, consistent with prior fMRI studies of patients with DoC.^13,20-23^

*Rs-fMRI analysis of the Default Mode Network (DMN)*

We estimated seed-based connectivity maps and ROI-to-ROI connectivity matrices to characterize the patterns of functional connectivity within the DMN. Functional connectivity strength was represented by Fisher-transformed bivariate correlation coefficients from a weighted general linear model (weighted-GLM)^24^, defined separately for each pair of seed and target areas, modeling the association between their BOLD signal timeseries. To compensate for possible transient magnetization effects at the beginning of each run, individual scans were weighted by a step function convolved with an SPM canonical hemodynamic response function and rectified.

Group-level analyses were performed using a GLM. For each individual voxel a separate GLM was estimated, with first-level connectivity measures at this voxel as dependent variables. Voxel-level hypotheses were evaluated using multivariable parametric statistics with random-effects across subjects and sample covariance estimation across multiple measurements. Inferences were performed at the level of individual clusters (groups of contiguous voxels). Cluster-level inferences were based on parametric statistics from Gaussian Random Field theory^25^. Results were thresholded using a combination of a cluster-forming p < 0.001 voxel-level threshold, and a familywise corrected p-FDR <0.05 cluster-size threshold.

**Supplementary Results**

*Behavioral Assessment*

Among the 45 participants, who were assessed 64 times with the CRS-R, 9 CRS-R assessments (in N=8 participants) had at least 1 invalid CRS-R subscale (14%; see Supplementary Table 5). For 4 of these assessments, we used the valid CRS-R subscales to assign a DoC diagnosis (e.g., if the motor or communication subscales indicate PTCS or if the auditory subscale suggested MCS+ and both the motor and communication subscales, which could indicate PTCS, were valid). In the remaining 5 assessments, we supplemented the information from the valid CRS-R subscales with clinical notes (e.g., valid CRS-R subscales indicate a diagnosis of VS and all notes from the clinical team, including those from clinicians who routinely perform the CRS-R, do not indicate any signs of MCS).

*Healthy Participants*

The healthy participant cohort was composed of 26 participants (mean±SD age 38.4±14.3 years, 11 [42%] male). Positive responses were observed in 22/26 (85%) healthy control participants for the active-motor EEG task and 22/26 (85%) for the active-motor fMRI task. Positive responses were observed in 25/25 (100%, n=1 was excluded due to data acquisition limitations) healthy control participants for passive-language EEG and 26/26 (100%) for passive-language fMRI (Supplementary Table 8).

**Supplementary Tables**

| **Supplementary Table 1: Coma Recovery Scale- Revised Test Completion Codes** | |
| --- | --- |
|  | **Test Attempted and completed** |
| **1.0** | Test completed in full, in person- results valid |
| **1.1** | Non-standard administration – a measure normally requiring an oral response, allowed a written response, results valid |
| **1.2** | Non-standard administration –Other (specify):__________________________________ |
| **1.3** | Test Completed, valid administration done over the phone |
|  | **Test Attempted but NOT completed** |
| **2.1** | Test attempted but not completed due to cognitive/neurological reason |
| **2.2** | Test attempted but not completed due to non-neurological/physical reasons |
| **2.3** | Test attempted but not completed - participant cognitively intact enough to respond but poor effort, random responding, rote response, not cooperative, refusal, intoxication |
| **2.4** | Test attempted but not completed due to major problems with English language proficiency (and/or Spanish language proficiency if the site can also enroll Spanish speaking subjects) |
| **2.5** | Test attempted but not completed due to test interrupted by illness and test could not be completed later |
| **2.6** | Test attempted but not completed due to logistical reasons, other reasons – site specific |
|  | **Test not attempted** |
| **3.1** | Test not attempted due to severity of cognitive/neurological deficits |
| **3.2** | Test not attempted due to non-neurological/physical reasons |
| **3.3** | Test not attempted - participant can respond appropriately but poor effort, not cooperative, refusal, intoxication |
| **3.4** | Test not attempted due to major problems with English language proficiency (and/or Spanish language proficiency if the site can also enroll Spanish speaking subjects) |
| **3.5** | Test not attempted due to participant illness and test could not be completed later |
| **3.6** | Test not attempted due to logistical reasons, other reasons – site specific |
| **4.0** | Test not attempted, completed or valid due to examiner error |
| **5.0** | Other (specify): |

| **Supplementary Table 2: Instructions administered prior to and during EEG and functional MRI** | | | |
| --- | --- | --- | --- |
|  | **Language** | **Motor Imagery**^a^ | **Resting State** |
| **Instructions**  **Prior to Stimulus** | EEG: Close your eyes and relax.  fMRI: When the MRI starts, close your eyes and relax.  Part of the time you will hear spoken words and part of the time you will hear silence. Keep your eyes closed the whole time. | EEG: Keep your eyes open and relax.  fMRI: When the MRI starts, keep your eyes open and relax.  After a few seconds, you will hear an instruction to imagine opening and closing your right hand. When you hear this, try to imagine that you are opening your right hand and then closing it into a fist. Concentrate on the way your muscles would feel if you were really doing this movement. Every time you hear the instruction to “imagine opening and closing your right hand,” imagine opening and closing your right hand over and over again until you hear an instruction to stop. Remember, do not actually open and close your right hand, just imagine. When you hear “stop opening and closing your right hand,” stop imagining opening and closing your hand, and relax. Remember, keep your eyes open the entire time. | fMRI: When the MRI starts, keep your eyes open and relax. Try not to move. Stay awake and let your thoughts wander. |
| **Instructions**  **During Stimulus** | Not applicable | At the beginning of Open Close Right Hand, subject hears “imagine opening and closing your right hand” At beginning of Rest, subject hears, “stop imagining opening and closing your hand.” | Not applicable |

^a^The instructions for the motor imagery task were adapted from Cruse et al. Lancet 2011;378:2088-94.

| **Supplementary Table 3: Regions of interest for stimulus-based functional MRI analysis** | | | |
| --- | --- | --- | --- |
| **ROI** | **Atlas** | **Atlas-Based Regions Included** | **Thresholding** |
| Heschl’s Gyrus | Harvard-Oxford Cortical Structural Atlas  (Makris *et al.*, 2006) | Heschl’s Gyrus (included H1 and H2) | 5% |
| Superior Temporal Gyrus | Harvard-Oxford Cortical Structural Atlas  (Makris *et al.*, 2006) | Superior Temporal Gyrus, anterior division +  Superior Temporal Gyrus, posterior division | 5% for both  component ROIs |
| Supplementary Motor Area/ Premotor Cortex | Harvard-Oxford Cortical Structural Atlas  (Makris *et al.*, 2006) and Juelich Histological Atlas  (Eickhoff *et al.*, 2005) | Juxtapositional Lobule Cortex (formerly Supplementary Motor Cortex) +  GM Premotor cortex BA6 L +  GM Premotor Cortex BA6 R | 5% for Juxtapositional Lobule Cortex ROI and 25% for GM Premotor Cortex ROIs |

Each region of interest (ROI) was constructed using atlas-based neuroanatomic regions distributed with FSLView v3.1.8 (Smith *et al.*, 2004). For the language and the music fMRI paradigms, we used the bilateral Heschl’s gyrus and superior temporal gyrus (anterior and posterior) ROIs distributed by the Harvard-Oxford Cortical Structural Atlas, each of which was thresholded at 5% and binarized using FSLMaths. For the motor imagery fMRI paradigm, we combined regions distributed by Harvard-Oxford Cortical Structural Atlas (Makris *et al.*, 2006) and the Juelich Histological Atlas (Eickhoff *et al.*, 2005). The Brodmann area (BA) 6 ROI of the Juelich Histological Atlas is comprised of the supplemental motor area (SMA), pre-SMA, and the four zones of the premotor cortex: premotor dorsal rostral (PMDr), premotor dorsal caudal (PMDc), premotor ventral rostral (PMVr), and premotor ventral caudal (PMVc). Of note, the pre-SMA is a common site of activation in fMRI motor imagery paradigms (Boly *et al.*, 2007). We added the Juxtapositional Lobule Cortex (i.e. SMA) ROI from the Harvard-Oxford Cortical Atlas because this ROI encompasses more SMA voxels along its inferior border than the Juelich BA6 ROI, and because it has been used previously in motor imagery fMRI studies of patients with DOC (Bardin *et al.*, 2011; Monti *et al.*, 2010). The BA6 ROIs from the Juelich Histological Atlas were thresholded at 25%, and the Juxtapositional Lobule Cortex ROI from the Harvard-Oxford Cortical Structural Atlas was thresholded at 5% prior to concatenation and binarization using FSLmaths.

| **Supplementary Table 4: Patient demographics and clinical characteristics** | | | | |
| --- | --- | --- | --- | --- |
| **ID** | **Age Range (yrs)** | **Sex** | **TBI Mechanism** | **iGCS** |
| P1 | 18-25 | M | MVA | 3T-7 |
| P2 | 46-50 | M | fall | 3T-8 |
| P3 | 66-70 | M | fall | 9-14 |
| P4 | 31-35 | M | MVA | 3T |
| P5 | 71-75 | M | fall | 6T |
| P6 | 71-75 | M | Fall | 3-8 |
| P7^a^ | 18-25 | F | car vs bicycle | 3T |
| P8 | 36-40 | M | MVA | 7T-8T |
| P9 | 61-65 | M | MVA | 7-13 |
| P10 | 26-30 | M | MVA | 3T-5 |
| P11 | 71-75 | F | Car vs pedestrian | 6T |
| P12 | 71-75 | M | MVA | 3T |
| P13 | 66-70 | F | Car vs pedestrian | 9-10 |
| P14 | 18-25 | M | MVA | 3 |
| P15 | 76-80 | M | Fall down stairs | 3-7T |
| P16 | 18-25 | F | MVA | 3T-5T |
| P17 | 76-80 | F | fall | 3-4 |
| P18 | 51-55 | M | Fall | 5T |
| P19 | 36-40 | F | Car vs. pedestrian | 3 |
| P20 | 56-60 | M | MVA | 3T |
| P21 | 18-25 | M | MVA | 3 |
| P22 | 18-25 | M | MVA | 3-7T |
| P23 | 36-40 | F | Car vs. pedestrian | 3T |
| P24 | 18-25 | M | MVA | 6 |
| P25 | 71-75 | M | Fall | 10/11 |
| P26 | 46-50 | M | fall | 3-3T |
| P27 | 56-60 | F | MVA | 7 |
| P28 | 66-70 | M | Gunshot wound | 9 |
| P29 | 18-25 | M | MVA | 5T-5 |
| P30 | 31-35 | M | Car vs pedestrian | 6-7 |
| P31 | 66-70 | F | Bike vs. bike | 9-12 |
| P32 | 26-30 | F | MVA | 3 |
| P33 | 31-35 | F | MVA | 4 |
| P34 | 18-25 | M | Car vs. pedestrian | 3 |
| P35 | 18-25 | M | Sport/recreation | 3-3T |
| P36 | 26-30 | M | MVA | 3T-6 |
| P37 | 26-30 | M | fall | 4-6 |
| P38 | 41-45 | M | fall | 7T-15 |
| P39 | 61-65 | M | Fall | 7-10 |
| P40 | 76-80 | F | fall | 3T-7T |
| P41 | 41-45 | F | Pedestrian vs bike | 10-14 |
| P42 | 18-25 | M | MVA | 3T-3 |
| P43 | 51-55 | M | fall | 4T |
| P44 | 56-60 | F | Car vs pedestrian | 4T-7 |
| P45 | 18-25 | M | MVA | 3T |
| The race of the patient cohort was 84% white. The initial GCS (iGCS) is defined as the best (i.e., highest) and worst (i.e., lowest) post-resuscitation GCS score assessed by a qualified clinician who performed a reliable examination (not confounded by sedation and/or paralytics) prior to ICU admission. Abbreviations: F *female*; GCS *Glasgow Coma Scale*; LoC *Level of Consciousness*; M *male*; MVA *motor vehicle accident*; N/A *not applicable*; TBI *traumatic brain injury* | | | | |

| **Supplementary Table 5: Details of CRS-R Behavioral Assessments** | | | | | | | | |  |  |  |  |
| --- | --- | --- | --- | --- | --- | --- | --- | --- | --- | --- | --- | --- |
| **ID** | **CRS-R**  **LoC at**  **EEG** | **CRS-R**  **Subscales at EEG** | **CRS-R TCC at EEG** | **CRS-R**  **LoC at**  **MRI** | **CRS-R**  **Subscales at MRI** | **CRS-R TCC at MRI** | | |  |  |  |  |
| P1 | Coma | NT | 3.1: Not administered due to heavy sedation and paralytics which were lifted for EEG and then applied again. Coma assigned based on pupils fixed and dilated, No active movement to any extremities, No withdrawal from pain | NA | | | | |  |  |  |  |
| P2 | Coma | A0V0M0O0C0Ar0 | 1.0 | Same as EEG | | | | |  |  |  |  |
| P3 | Coma | A0V0M0O0Ar0 | 3.1: CRS-R not safe to perform due to intracranial pressure elevation during examination. Coma assigned based on all clinical notes indicating GCS 3T and coma diagnosis | NA | | | | |  |  |  |  |
| P4 | Coma | A0V0M2O0C0C0 | 1.0 | Same as EEG | | | | |  |  |  |  |
| P5 | Coma | A0V0M2O1C0Ar0 | 1.0 | Coma | A0V0M0O0C0Ar0 | | | 1.0 |  | |  | |
| P6 | Coma | A0V0M0O0C0Ar0 | 1.0 | NA | | | | |  |  |  |  |
| P7 | VS | A2V0M2O1C0Ar1 | 1.0 | Same as EEG | | | | |  |  |  |  |
| P8 | VS | A0V0M0O0C0Ar1 | 1.0 | Same as EEG | | | | |  |  |  |  |
| P9 | VS | A0V0M0O1C0Ar0 | 1.0 | Same as EEG | | | | |  |  |  |  |
| P10 | VS | A0V0M2O0C0Ar1 | 1.0 | Same as EEG | | | | |  |  |  |  |
| P11 | VS | A0V0M0O1C0Ar2 | 1.0 | Same as EEG | | | | |  |  |  |  |
| P12 | VS | A0V0M1O0C0Ar1 | 1.0 | VS | A0V0M1O0C0Ar1 | | | 1.0 |  |  |  |  |
| P13 | VS | A0V0M2O0C0Ar1 | 1.0 | Same as EEG | | | | |  |  |  |  |
| P14 | VS | A0V0M0O2C0Ar1 | 1.0 | NA | | | | |  |  |  |  |
| P15 | VS | A0V0M2O1C0Ar1 | 1.0 | Same as EEG | | | | |  |  |  |  |
| P16 | VS | A0V0M2O0C0Ar1 | 1.0 | NA | | | | |  |  |  |  |
| P17 | VS | A0V1M2O1C0Ar1 | 1.0 | NA | | | | |  |  |  |  |
| **Supplementary Table 5 Continued** | | | | | | | | |  |  |  |  |
| **ID** | **CRS-R**  **LoC at**  **EEG** | **CRS-R**  **Subscales at EEG** | **CRS-R TCC at EEG** | **CRS-R**  **LoC at**  **MRI** | **CRS-R**  **Subscales at MRI** | | **CRS-R TCC at MRI** | |  |  |  |  |
| P18 | VS | A0V0M2O0C0Ar0 | 1.0 | NA | | | | |  |  |  |  |
| P19 | VS | A0V0M2O0C0Ar1 | 1.0 | VS | A0V0M2O0C0Ar1 | | | 1.0 |  |  |  |  |
| P20 | VS | A0V0M2O0C0Ar0 | 1.0 | Same as EEG | | | | |  |  |  |  |
| P21 | VS | A0V[NT]M[NT]O0C0Ar2 | 2.1: Motor and visual subscales Not completed due to neurologic injury - craniocervical dissociation, resulting in quadriplegia. VS diagnosis assigned based on clinical notes | VS | A0V[NT]M[NT]O0C0Ar2 | | | 2.1: Motor and visual subscales not completed due to neurologic injury - craniocervical dissociation, quadriplegia. VS assigned based on clinical notes |  |  |  |  |
| P22 | VS | A0V0M2O1C0Ar1 | 1.0 | VS | A0V1M2O1C0Ar1 | | | 1.0 |  |  |  |  |
| P23 | VS | A0V0M2O1C0Ar0 | 1.0 | Same as EEG | | | | |  |  |  |  |
| P24 | VS | A0V1M2O0C0Ar2 | 1.0 | Same as EEG | | | | |  |  |  |  |
| P25 | VS | A0V0M0O1C0Ar0 | 1.0 | Same as EEG | | | | |  |  |  |  |
| P26 | MCS- | A0V0M3O0C0Ar0 | 1.0 | Same as EEG | | | | |  |  |  |  |
| P27 | MCS- | A0V0M5O0C0Ar0 | 1.0 | VS | A0V0M2O2C0Ar0 | | | 1.0 |  | |  | |
| P28 | MCS- | A0V[NT]M5O2C0Ar[NT] | 2.2: Gun shot wound to the head, eyes sutured/slits, cannot administer visual subscale. MCS- assigned based on unconfounded subscales | NA | | | | |  |  |  |  |
| P29 | MCS- | A0V1M5O0C0Ar1 | 1.0 | NA | | | | |  |  |  |  |
| P30 | MCS- | A1V1M3O0C0A1 | 1.0 | MCS- | A0V0M3O0C0A0 | 1.0 | | |  |  |  |  |
| **Supplementary Table 5 Continued** | | | | | | | | |  |  |  |  |
| **ID** | **CRS-R**  **LoC at**  **EEG** | **CRS-R**  **Subscales at EEG** | **CRS-R TCC at EEG** | **CRS-R**  **LoC at**  **MRI** | **CRS-R**  **Subscales at MRI** | **CRS-R TCC at MRI** | | |  |  |  |  |
| P31 | MCS- | A2V0M5O1C0Ar0 | 1.0 | MCS+ | A4V5M6O1C0Ar3 | 1.0 | | |  |  |  |  |
| P32 | MCS- | A0V0M3O1C0Ar1 | 1.0 | MCS- | A0V4M5O2C0Ar1 | 1.0 | | |  | | | 1.0 |
| P33 | MCS- | A0V0M5O2C0Ar1 | 1.0 | NA | | | | |  |  |  |  |
| P34 | MCS- | A0V0M5O0C0Ar1 | 1.0 | MCS- | A1V0M5O0C0Ar2 | 1.0 | | |  | | | 1.0 |
| P35 | MCS- | A1V2M5O1C0Ar2 | 1.0 | MCS- | A2V4M5O3C0Ar2 | 1.0 | | |  | | |  |
| P36 | MCS+ | A4V1M4O2C0Ar2 | 1.0 | MCS+ | A4V4M4O2C0Ar2 | 1.0 | | |  |  | | |
| P37 | MCS+ | A4V3M4O2C0Ar1 | 1.0 | Same as EEG | | | | |  |  |  |  |
| P38 | MCS+ | A3VNTMNTO2C0Ar1 | 2.2: Eye lids could not be opened due to injury. MCS+ assigned based on unconfounded subscales | NA | | | | |  |  |  |  |
| P39 | MCS+ | A3V4M5O3C1A1 | 1.0 | PTCS | A4V5M6O3C2Ar3 | 1.0 | | |  |  |  |  |
| P40 | MCS+ | A3V3M2O0C0Ar1 | 1.0 | MCS+ | A3V5M5O1C1Ar2 | 1.0 | | |  |  |  |  |
| P41 | MCS+ | A2V4M[NT]O[NT]C[NT]Ar1 | 2.3: Alert and nodded when examiner introducing themselves, subsequently agitated and disinterested. Eyes remained closed with repeated stim. Nurse requested assessment be stopped due to time constraints. MCS+ assigned based on clinical notes immediately preceding and following assessment consistently indicate command-following | NA | | | | |  |  |  |  |
| P42 | NA^e^ | A3V[NT]M5O0C0Ar[NT] | 2.2: Bilateral periorbital and lid ecchymosis, lid edema. MCS+ assigned based on unconfounded subscales | Same as EEG | | | | |  |  |  |  |

| **Supplementary Table 5 Continued** | | | | | | |
| --- | --- | --- | --- | --- | --- | --- |
| **ID** | **CRS-R**  **LoC at**  **EEG** | **CRS-R**  **Subscales at EEG** | **CRS-R TCC at EEG** | **CRS-R**  **LoC at**  **MRI** | **CRS-R**  **Subscales at MRI** | **CRS-R TCC at MRI** |
| P43 | PTCS | A4V5M6O3C2Ar3 | 1.0 | NA | | |
| P44 | PTCS | A4V5M6O2C0Ar2 | 1.0 | NA | | |
| P45 | NA | | | PTCS | A4V5M[NT]O3C2Ar1 | 2.2: Extensive burns, upper extremities bandaged, could not assess motor subscale. PTCS assigned based on Communication Subscale |
| Level of Consciousness (LoC) is assessed via behavioral evaluation with the Coma Recovery Scale-Revised (CRS-R) as coma, vegetative state (VS), minimally conscious state without language function (MCS-), minimally conscious state with language function (MCS+), or post-traumatic confusional state (PTCS; emerged from MCS but disoriented). Abbreviations: A *Auditory Subscale*; Ar *Arousal Subscale*; C *Communication Subscale*; EEG *electroencephalography*; fMRI *functional magnetic resonance imaging*; M *Motor Subscale*; N/A *not applicable*; NT *Not Testable* (i.e., CRSR scale or subscore could not be administered due to confounding factors); O *Oromotor/verbal Subscale*; TCC *Test Completion Code*; V *Visual Subscale* | | | | | | |

| **Supplementary Table 6: Sedative, anxiolytic and analgesic medications administered during functional MRI and EEG** | | | | |
| --- | --- | --- | --- | --- |
| **ID** | **Sedating Medications Administered**  **Before (within 4 hours prior) and During EEG** | | **Medications Administered**  **Before (within 4 hours prior) and During fMRI** | |
|  | **Before EEG** | **During EEG** | **Before fMRI** | **During fMRI** |
| P1 | Cisatracurium 10mcg/kg/min  Ketamine 10 mcg/min/kg  Versed 6 mcg/hr (1mg/ml)  Fentanyl 300 mcg/hr (50 mcg/ml)  Norepinephrine 40 mcg/min  Vasopressin 0.08 units/min | Cisatracurium 10mcg/kg/min  Ketamine 10 mcg/min/kg  Versed 6 mcg/hr (1mg/ml)  Fentanyl 300 mcg/hr (50 mcg/ml)  Norepinephrine 40 mcg/min  Vasopressin 0.08 units/min | NA | NA |
| P2 | Precedex 0.3 mcg/kg/hr  Nicardipine 2.5 mg/hr  Oxycodone 10mg | Precedex 0.3 mcg/kg/hr  Nicardipine 2.5 mg/hr | Precedex 0.3 mcg/kg/hr | Precedex 0.3 mcg/kg/hr |
| P3 | Propofol 83 mcg/kg/min (stopped 2.5 hours prior to EEG) | Nicardipine 10 mg/hr | NA | NA |
| P4 | Fentanyl 175 mcg/hr  Precedex 0.8 mcg/kg/hr | Fentanyl 175 mcg/hr  Precedex 0.8 mcg/kg/hr | Fentanyl 200 mcg/hr  Precedex 0.8 mcg/kg/hr | Fentanyl 200 mcg/hr  Precedex 0.8 mcg/kg/hr |
| P5 | Propofol 40 mcg/kg/min  Norepinephrine 4mcg/min | Fentanyl 50mcg bolus  Propofol 40 mcg/kg/min  Norepinephrine 4mcg/min | NA | Fentanyl 100mcg bolus |
| P6 | Propofol 83 mcg/kg/min | Propofol 50 mcg/kg/min Bolus | NA | NA |
| P7 | Dilaudid 4mg, Gabapentin 900mg | None | Dilaudid 4mg | None |
| P8 | Propofol 45 mcg/kg/min continuous | Propofol 45 mcg/kg/min continuous mg, Keppra 500mg | Propofol 45 mcg/kg/min continuous,  Dilaudid 0.5mg bolus | Propofol 45 mcg/kg/min continuous |
| P9 | Propofol 50 mcg/kg/min  Norepinephrine 2 mcg/min | Propofol 40 mcg/kg/min | Propofol 50-70 mcg/kg/min  Dilaudid 1 mg bolus | Norepinephine 80 mcg/mL  Propofol 80 mcg/kg/min |

| **Supplementary Table 6 Continued** | | | | |
| --- | --- | --- | --- | --- |
| **ID** | **Sedating Medications Administered**  **Before (within 4 hours prior) and During EEG** | | **Medications Administered**  **Before (within 4 hours prior) and During fMRI** | |
|  | **Before EEG** | **During EEG** | **Before fMRI** | **During fMRI** |
| P10 | Dilaudid 0.5 mg/hr  Ketamine 8 mcg/kg/min  Norepinephrine 1-3 mcg/min | Propofol 30 mcg/kg/min  Dilaudid 0.5 mg/hr  Ketamine 8 mcg/kg/min  Norepinephrine 3 mcg/min | Dilaudid 2 mg/hr  Dilaudid 1 mg bolus | Dilaudid 2 mg/hr  Ketamine 8 mcg/kg/min  Norepinephrine 10 mcg/min  Propofol 50 mcg/kg/min  Vassopressin 0.04 units/min |
| P11 | Nicardipine 5 mg/hr | Nicardipine 5 mg/hr | Propofol 50 mcg/kg/min  Nicardipine 5 mg/hr  Dilaudid 1mg  Norepinephrine 1 mcg/min | Propofol 50 mcg/kg/min  +bolus 20 mg  Norepinephrine 1 mcg/min |
| P12 | Propofol 40 mcg/kg/min  Dilaudid 0.7 mg/hr | Propofol 40 mcg/kg/min  Dilaudid 0.7 mg/hr | Propofol 20 mcg/kg/min  Precedex 0.4-0.6 mcg/kg/hr | Propofol 20 mcg/kg/min  Precedex 0.6-0.8 mcg/kg/hr |
| P13 | Propofol 35-50 mcg/kg/min  Fentanyl 35 mcg/hr  Fentanyl 25 mcg bolus | Propofol 30 mcg/kg/min  Fentanyl 35 mcg/hr | Propofol 50 mcg/kg/min  Fentanyl 25 mcg bolus | Propofol 55 mcg/kg/min |
| P14 | None | None | NA | NA |
| P15 | Propofol 30 mcg/kg/min  Fentanyl 25 mcg/hr | Propofol 30 mcg/kg/min  Fentanyl 25 mcg/hr | Propofol 30 mcg/kg/min  Fentanyl 75 mcg/hr | Propofol 30 mcg/kg/min  Fentanyl 75 mcg/hr |
| P16 | Propofol 40 mcg/kg/min  Dilaudid 0.5 mg/hr | Propofol 40 mcg/kg/min  Dilaudid 0.5 mg/hr | NA | NA |
| P17 | None | None | NA | NA |
| P18 | Propofol 40 mcg/kg/min  Phenobarbital 130mg | Propofol 40 mcg/kg/min | NA | NA |
| P19 | Propofol 30 mcg/kg/min  Fentanyl 50 mcg/hr | Propofol 30 mcg/kg/min  Fentanyl 50 mcg/hr | NA | Propofol 40 mcg/kg/min |
| P20 | Propofol 50 mcg/kg/min continuous | Propofol 50 mcg/kg/min continuous | Dilaudid 0.5 mg | Propofol 25 mcg/kg/min continuous |
| 21 | None | None | None | None |
| **Supplementary Table 6 Continued** | | | | |
| **ID** | **Sedating Medications Administered**  **Before (within 4 hours prior) and During EEG** | | **Medications Administered**  **Before (within 4 hours prior) and During fMRI** | |
|  | **Before EEG** | **During EEG** | **Before fMRI** | **During fMRI** |
| P22 | Propofol 20 mcg/kg/min  Precedex 0.7-0.8 mcg/kg/hr  Fentanyl 60 mcg/hr  Norepinephrine 2mcg/min | Precedex 0.7-0.8 mcg/kg/hr  Fentanyl 60 mcg/hr  Norepinephrine 2mcg/min | Propofol 60 mcg/kg/min  Precedex 0.5 mcg/kg/hr  Oxycontin 5mg bolus | Propofol 83 mcg/kg/min  Precedex 0.5 mcg/kg/hr  Precedex 20 mcg bolus |
| P23 | Propofol 15 mcg/kg/min  Precedex 0.5 mcg/kg/hr | Propofol 15 mcg/kg/min  Precedex 0.5 mcg/kg/hr | Propofol 40mcg/kg/min  Precedex 0.7 mcg/kg/hr | Propofol 83 mcg/kg/min  Propofol 10mcg bolus |
| P24 | None | None | None | None |
| P25 | None | None | None | None |
| P26 | Propofol 60 mcg/kg/min | Propofol 60 mcg/kg/min  Fentanyl 25 mcg/hr | Propofol 50 mcg/kg/min  Fentanyl 25 mcg/hr | Propofol 60 mcg/kg/min |
| P27 | Propofol 40 mcg/kg/min  Nicardipine 5 mg/hr  Fentanyl 25 mcg/hr (50 mcg/ml) | Propofol 40 mcg/kg/min (for the first ~30mins)  Nicardipine 5 mg/hr  Fentanyl 25 mcg/hr (50 mcg/ml) for the first ~30mins) | Propofol 65 mcg/kg/min  Nicardipine 5 mg/hr | Propofol 65 mcg/kg/min  Nicardipine 2.5 mg/hr |
| P28 | Seroquel 200mg | None | NA | NA |
| P29 | Precedex 0.3 mcg/kg/hr | Precedex 0.4 mcg/kg/hr | NA | NA |
| P30 | Propofol 30 mcg/kg/min  Fentanyl 50 mcg/hr + bolus 100mcg | Propofol 30 mcg/kg/min  Fentanyl 50 mcg/hr | Propofol 60 mcg/kg/min  Fentanyl 50 mcg/hr  Precedex 0.9 mcg/kg/hr | Propofol 70 mcg/kg/min, + bolus 20 mcg  Fentanyl 50 mcg bolus x 2 |
| P31 | Norepinephrine 2mcg/min  Precedex 0.5 mcg/kg/hr | Norepinephrine 2mcg/min  Precedex 0.5 mcg/kg/hr | Precedex 0.4 mcg/kg/hr | Precedex 0.5 mcg/kg/hr  Fentanyl 100mg bolus |
| P32 | Propofol 20 mcg/kg/min  Dilaudid 0.5 mg/hr | Propofol 20 mcg/kg/min and bolus 2cc  Dilaudid 0.5 mg/hr | Propofol 10-20 mcg/kg/min  Dilaudid 0.5 mg/hr continuous and 3 x 1 mg bolus | Propofol 50 mcg/kg/min Norepinephrine 8 mcg/min bolus at end of scan |
| P33 | None | None | NA | NA |
| **Supplementary Table 6 Continued** | | | | |
| **ID** | **Sedating Medications Administered**  **Before (within 4 hours prior) and During EEG** | | **Medications Administered**  **Before (within 4 hours prior) and During fMRI** | |
|  | **Before EEG** | **During EEG** | **Before fMRI** | **During fMRI** |
| P34 | Precedex 0.8 mcg/kg/hr | Precedex 0.8 mcg/kg/hr  Fentanyl 50 | None | None |
| P35 | Propofol 83 mcg/kg/min | Propofol 83 mcg/kg/min | Propofol 70-75 mcg/kg/min | Propofol 70-75 mcg/kg/min  Propofol bolus 10 mg/mL  Fentanyl bolus 70mcg |
| P36 | Norepinephrine 5mcg/min  Precedex 0.3 mcg/kg/hr  Dilaudid 2 mg/hr  Versed 2.5 mg/hr | Norepinephrine 5mcg/min  Precedex 0.3 mcg/kg/hr  Dilaudid 2 mg/hr  Versed 2.5 mg/hr | Labetelol 0.2 mcg/kg/hr | Labetelol 1.5 mg/min |
| P37 | Fentanyl 25 mcg/hr  Precedex 1.2 mcg/kg/hr  Propofol 30 mcg/kg/min | Fentanyl 25, then 37.5 mcg/hr  Fentanyl 50 mcg bolus  Precedex 1.2 then 1.3 mcg/kg/hr | Precedex 1.3 mcg/kg/hr  Fentanyl 37.5 mcg/hr  Fentanyl 100mcg bolus  Propofol 40 mcg/kg/min | Precedex 1.3 mcg/kg/hr  Propofol 50 mcg/kg/min  Propofol 20 mcg bolus |
| P38 | Propofol 20 mcg/kg/min | Propofol 10-20 mcg/kg/min, then turned off | NA | NA |
| P39 | Clonadine 0.1mg  Clonadine 0.2mg | None | None | None |
| P40 | Precedex 0.3 mcg/kg/hr | Precedex 0.3 mcg/kg/hr | None | None |
| P41 | Precedex 0.7 mcg/kg/hr  Oxycodone 10mg bolus | Precedex 0.7 mcg/kg/hr | NA | NA |
| P42 | Propofol 65mcg/kg/min | Propofol 65mcg/kg/min | Propofol 80mcg/kg/min | Propofol 80mcg/kg/min |
| P43 | None | None | NA | NA |
| P44 | 10mg | NA | NA | NA |
| P45 | NA | NA | Dilaudid 0.25mg | NA |
| Sedative, anxiolytic and analgesic medications were administered as needed during EEG and fMRI to maintain patient safety and comfort | | | | |

| **Supplementary Table 7: Drains, Lines, Tubes, and Monitors at time of EEG and fMRI** | | | | | | | | | | | | | | | | | | | | | | | |
| --- | --- | --- | --- | --- | --- | --- | --- | --- | --- | --- | --- | --- | --- | --- | --- | --- | --- | --- | --- | --- | --- | --- | --- |
| **ID** | **EEG** | | | | | | | | | | | | **fMRI** | | | | | | | | | | |
|  | EVD | ICP Monitor | ETT | Trach | OGT | NGT | PEG | CVC | A-line | Foley | RT | Other | EVD | ETT | Trach | OGT | NGT | PEG | CVC | A-line | Foley | RT | Other |
| P1 | + | - | + | - | - | + | - | + | + | + | - | - | NA^a^ | | | | | | | | | | |
| P2 | + | - | - | - | + | - | - | + | - | + | - | - | + | - | - | + | - | - | + | - | + | - | - |
| P3 | + | - | + | - | + | - | - | + | + | + | + | - | NA^b^ | | | | | | | | | | |
| P4 | - | - | + | - | + | - | - | - | + | + | + | - | - | + | + | + | - | - | + | + | + | + | - |
| P5 | - | + | + | - | + | - | - | + | + | + | - | SDD | - | + | - | + | - | - | + | + | + | + | - |
| P6 | - | + | + | - | + | - | - | + | - | - | - | - | NA^b^ | | | | | | | | | | |
| P7 | - | - | - | + | - | + | - | - | - | + | + | - | - | - | + | - | + | - | - | - | + | + | - |
| P8 | + | - | + | - | + | - | - | + | + | + | - | - | + | + | - | + | - | - | + | + | + | - | - |
| P9 | - | - | + | - | + | - | - | + | + | + | - | - | - | + | - | + | - | - | + | + | + | - | - |
| P10 | - | - | + | - | + | - | - | + | + | + | - | - | - | + | - | + | - | - | + | + | + | - | - |
| P11 | - | - | + | - | - | - | - | - | + | - | - | - | - | + | - | - | + | - | + | + | + | - | - |
| P12 | + | - | + | - | + | - | - | - | + | + | - | - | + | + | - | + | - | - | + | + | + | + | - |
| P13 | - | - | + | - | + | - | - | - | + | + | - | - | - | + | - | + | - | - | - | + | + | - | - |
| P14 | - | - | - | + | - | - | + | + | - | - | + | - | NA^c^ | | | | | | | | | | |
| P15 | - | - | + | + | + | - | - | + | + | + | - | - | - | + | + | + | - | - | + | + | + | - | - |
| P16 | - | + | + | - | + | - | - | + | + | + | - | - | NA^d^ | | | | | | | | | | |
| P17 | - | - | + | - | + | - | - | - | + | + | - | - | NA^b^ | | | | | | | | | | |
| P18 | - | - | + | - | + | - | - | + | + | + | - | - | NA^e^ | | | | | | | | | | |
| P19 | + | + | + | - | + | - | - | + | + | + | - | - | - | - | + | - | - | + | + | - | - | + | - |

| **Supplementary Table 7 Continued** | | | | | | | | | | | | | | | | | | | | | | | |
| --- | --- | --- | --- | --- | --- | --- | --- | --- | --- | --- | --- | --- | --- | --- | --- | --- | --- | --- | --- | --- | --- | --- | --- |
| **ID** | **EEG** | | | | | | | | | | | | **fMRI** | | | | | | | | | | |
|  | EVD | ICP Monitor | ETT | Trach | OGT | NGT | PEG | CVC | A-line | Foley | RT | Other | EVD | ETT | Trach | OGT | NGT | PEG | CVC | A-line | Foley | RT | Other |
| P20 | - | - | - | - | - | - | - | - | - | - | - | CT | - | + | - | + | - | - | + | + | + | - | CT |
| P21 | - | - | - | - | - | - | - | + | + | + | + | - | - | + | - | + | - | - | + | - | + | + | - |
| P22 | - | + | + | - | + | + | - | + | + | + | + | - | - | - | - | - | - | - | - | + | + | + | - |
| P23 | - | - | + | - | + | - | - | + | - | + | + | - | - | + | - | + | - | - | + | - | + | + | - |
| P24 | - | - | - | + | - | - | + | - | - | - | - | - | - | - | + | - | - | + | - | - | - | - | - |
| P25 | - | - | + | - | - | + | - | + | + | + | + | - | - | + | - | - | + | - | + | + | + | + | - |
| P26 | - | - | + | - | + | - | _ | + | + | + | - | - | - | + | - | + | - | - | + | + | + | + | - |
| P27 | - | - | + | - | + | - | - | + | + | + | - | - | - | + | - | + | - | - | + | + | + | + | - |
| P28 | - | - | - | + | - | - | + | + | - | + | - | PICC | NA^a^ | | | | | | | | | | |
| P29 | + | - | + | + | + | - | - | + | + | + | + | - | NA^c,f^ | | | | | | | | | | |
| P30 | - | + | + | - | + | - | - | + | + | + | - | - | - | + | - | + | - | - | + | + | + | - | - |
| P31 | - | - | + | - | - | + | - | + | - | + | - | - | - | + | - | - | + | - | + | - | + | + | - |
| P32 | - | + | - | + | - | - | + | + | + | + | - | - | - | + | - |  | - | + | + | - | + | - | - |
| P33 | - | - | - | + | - | - | + | - | - | - | - | - | NA^f^ | | | | | | | | | | |
| P34 | + | - | - | + | - | - | + | - | + | + | + | - | - | - | + | - | - | + | - | - | - | + | - |
| P35 | - | - | + | - | - | + | - | + | + | + | + | - | - | + | - | - | + | - | + | + | + | + | - |
| P36 | + | - | + | - | + | - | - | + | + | + | - | - | - | - | + | - | - | + | - | + | - | + | - |
| P37 | - | - | + | - | + | - | - | + | + | + | + | - | - | + | - | + | - | - | + | + | + | + | - |
| P38 | - | + | - | + | - | - | - | + | + | + | + | SDD | NA^c^ | | | | | | | | | | |
| P39 | - | - | - | - | - | + | - | - | - | - | - | - | - | - | - | - | + | - | - | - | - | - | - |
| P40 | - | + | + | - | + | - | - | + | + | + | - | - | - | + | - | + | - | - | - | - | - | - | - |

| **Supplementary Table 7 Continued** | | | | | | | | | | | | | | | | | | | | | | | |
| --- | --- | --- | --- | --- | --- | --- | --- | --- | --- | --- | --- | --- | --- | --- | --- | --- | --- | --- | --- | --- | --- | --- | --- |
| **ID** | **EEG** | | | | | | | | | | | | **fMRI** | | | | | | | | | | |
|  | EVD | ICP Monitor | ETT | Trach | OGT | NGT | PEG | CVC | A-line | Foley | RT | Other | EVD | ETT | Trach | OGT | NGT | PEG | CVC | A-line | Foley | RT | Other |
| P41 | + | - | + | - | + | - | - | - | + | + | - | - | NA^d^ | | | | | | | | | | |
| P42 | + | - | + | - | + | - | - | + | + | + | - | - | + | + | - | + | - | - | + | + | + | - | - |
| P43 | - | - | - | - | - | - | - | + | - | + | - | - | NA^c^ | | | | | | | | | | |
| P44 | - | - | - | + | - | - | + | + | - | + | - | PICC | NA^c^ | | | | | | | | | | |
| P45 | NA^g^ | | | | | | | | | | | | - | - | - | - | + | - | - | - | + | + | NC, CT |
| ^a^ Medical instability  ^b^ Code status established as Comfort Measures Only before data could be acquired.  ^c^ MRI contraindication: metallic fragment in the eye  ^d^ Unable to attain MRI due to restlessness  ^e^ Clinical MRI initially obtained prior to consent for research, subsequently too restless for research MRI  ^f^ Unable to schedule MRI due to logistical/scheduling issue and/or scanner unavailability.  ^g^ Could not place EEG leads due to scalp burns  Abbreviations: fMRI *functional magnetic resonance imaging*; EEG *electroencephalogram*; NA *Not applicable*; EVD *external ventricular drain*; ETT *endotracheal tube*; Trach *tracheostomy*; OGT *orogastric tube*; NGT *nasogastric tube*; PEG *percutaneous endoscopic gastrostomy*; CVC *central venous catheter*; A-line *arterial line*; Foley *urinary catheter*; RT *rectal tube*; ICP *Intracranial Pressure*; CT *Chest Tube*; NC *Nasal Cannula*; SDD *Subdural Drain*,+ *present*; - *not present* | | | | | | | | | | | | | | | | | | | | | | | |

| **Supplementary Table 8: Healthy participant responses to functional MRI and EEG assessments** | | | | | | | | | |
| --- | --- | --- | --- | --- | --- | --- | --- | --- | --- |
| **ID** | **Age** | **Sex** | **EEG Predictors** | | **fMRI Predictors** | | | | |
|  |  |  | **Hand Motor Imagery Paradigm +/- CMD (Accuracy)** | **Passive Language Paradigm**  **+/- CCP (Accuracy)** | **Hand Motor Imagery Paradigm** | | **Passive Language**  **Paradigm** | | **Resting-state** |
|  |  |  |  |  | **+/- CMD**  **(% SMA/PMC voxels)** | **SMA/PMC**  **Z statistic** | **+/- CCP**  **(% STG**  **Voxels)** | **STG**  **Z statistic** | **DMN**  **Z statistic** |
| C1 | 31-35 | F | +(0.59) | +(0.64) | + (4.37) | 0.030 | + (68.68) | 5.60 | 0.78 |
| C2 | 31-35 | M | +(0.72) | +(0.64) | + (10.31) | 0.41 | + (64.38) | 5.49 | 0.99 |
| C3 | 46-50 | F | +(0.61) | +(0.64) | + (9.40) | 1.30 | + (56.47) | 4.43 | 1.20 |
| C4 | 26-30 | M | +(0.71) | +(0.72) | + (3.64) | -0.40 | +(56.66) | 4.82 | 0.77 |
| C5 | 26-30 | F | +(0.68) | +(0.72) | + (13.87) | 0.95 | +(60.14) | 5.11 | 1.02 |
| C6 | 36-40 | F | +(0.63) | N/A ^a^ | + (0.78) | -1.05 | +(62.77) | 5.42 | 0.76 |
| C7 | 26-30 | F | +(0.75) | +(0.68) | + (11.83) | 0.15 | +(50.28) | 3.83 | 0.74 |
| C8 | 31-35 | F | -(0.51) | +(0.76) | - (0) | -0.02 | +(47.26) | 3.85 | 0.63 |
| C9 | 51-55 | M | -(0.51) | +(0.61) | - (0) | -1.24 | +(22.02) | 1.11 | 0.80 |
| C10 | 26-30 | F | +(0.57) | +(0.71) | + (25.44) | 2.01 | +(63.89) | 5.78 | 0.65 |
| C11 | 36-40 | F | +(0.64) | +(0.70) | + (13.86) | 1.01 | +(50.72) | 3.50 | 0.57 |
| C12 | 31-35 | M | +(0.75) | +(0.73) | + (21.27) | 1.49 | +(46.70) | 3.36 | 1.19 |
| C13 | 18-25 | F | +(0.70) | +(0.67) | + (0.18) | -3.13 | +(50.33) | 3.72 | 0.79 |
| C14 | 31-35 | M | +(0.56) | +(0.68) | - (0) | -0.34 | +(17.31) | 1.11 | 1.05 |
| C15 | 56-60 | M | +(0.66) | +(0.65) | + (13.13) | 0.66 | +(61.69) | 5.46 | 0.67 |

| **Supplementary Table 8 Continued** | | | | | | | | | |
| --- | --- | --- | --- | --- | --- | --- | --- | --- | --- |
| **ID** | **Age** | **Sex** | **EEG Predictors** | | **fMRI Predictors** | | | | |
|  |  |  | **Hand Motor Imagery Paradigm +/- CMD (Accuracy)** | **Passive Language Paradigm**  **+/- CCP (Accuracy)** | **Hand Motor Imagery Paradigm** | | **Passive Language**  **Paradigm** | | **Resting-state** |
|  |  |  |  |  | **+/- CMD**  **(% SMA/PMC voxels)** | **SMA/PMC**  **Z statistic** | **+/- CCP**  **(% STG**  **Voxels)** | **STG**  **Z statistic** | **DMN**  **Z statistic** |
| C16 | 18-25 | M | -(0.54) | +(0.66) | + (6.63) | 1.22 | +(36.25) | 2.46 | 0.50 |
| C17 | 18-25 | M | +(0.81) | +(0.67) | + (0.18) | -2.72 | +(33.56) | 2.58 | 1.17 |
| C18 | 31-35 | F | +(0.77) | +(0.64) | + (5.10) | -0.21 | +(26.01) | 1.72 | 0.57 |
| C19 | 36-40 | F | +(0.59) | +(0.75) | + (6.517) | 0.05 | +(64.10) | 5.33 | 0.89 |
| C20 | 18-25 | M | +(0.58) | +(0.70) | - (0) | -0.55 | +(36.21) | 2.35 | 0.62 |
| C21 | 26-30 | F | +(0.59) | +(0.69) | + (0.09) | -0.93 | +(62.80) | 5.44 | 0.99 |
| C22 | 61-65 | F | +(0.65) | +(0.68) | + (6.92) | 0.99 | +(63.21) | 4.97 | 0.75 |
| C23 | 61-65 | M | +(0.81) | +(0.72) | + (21.70) | 1.69 | +(46.02) | 3.73 | 0.89 |
| C24 | 66-70 | F | -(0.53) | +(0.64) | + (0.001) | -0.001 | +(8.99) | 0.60 | 0.61 |
| C25 | 31-35 | F | +(0.71) | +(0.71) | + (2.96) | -0.41 | +(63.68) | 5.59 | 0.91 |
| C26 | 66-70 | M | +(0.78) | +(0.69) | +(13.14) | 1.00 | +(49.74) | 3.75 | 0.83 |
| **Group Median**  **(95% CI)** | | | **0.65**  **(0.58, 0.72)** | **0.68**  **(0.66, 0.72)** | **5.81**  **(0.18, 12.81)** | **0.04**  **(-0.41, 1.00)** | **50.53**  **(38.69, 62.79)** | **3.84**  **(2.78, 5.39)** | **0.78 (0.65,1.06)** |
| **N (%) Positive** | | | **22/26 (85%)** | **25/25 (100%)** | **22/25 (85%)** | **NA** | **26/26 (100%)** | **NA** | **NA** |
| ^a^ Only 1 block of passive language EEG data available  The race of the control cohort was 81% white.  Abbreviations: EEG *electroencephalography;* F *female*; fMRI *functional magnetic resonance imaging*; Heschl *Heschl’s gyrus*; M *male*; SMA/PMC *supplementary motor area/premotor cortex*; STG *superior temporal gyrus* | | | | | | | | | |

| **Supplementary Table 9: Results of EEG and fMRI tests for participants who were not followed at 6-months** | | | | | | | | | | | |
| --- | --- | --- | --- | --- | --- | --- | --- | --- | --- | --- | --- |
| **ID** | **EEG Predictors** | | | | **fMRI Predictors** | | | | | | |
|  | **Day of**  **EEG** | **CRS-R LoC at**  **EEG** | **Hand Paradigm +/- CMD (Accuracy)** | **Language Paradigm**  **+/- CCP (Accuracy)** | **Day**  **of**  **fMRI** | **CRS-R LoC at fMRI** | **Hand Motor Imagery Paradigm** | | **Passive Language**  **Paradigm** | | **Resting-state** |
|  |  |  |  |  |  |  | **+/- CMD**  **(% SMA/PMC voxels)** | **SMA/PMC**  **Z statistic** | **+/- CCP**  **(% STG**  **Voxels)** | **STG**  **Z statistic** | **+/- DMN**  **(Z statistic)** |
| P46 | 19 | MCS- | - (0.49) | + (0.56) | 18 | MCS- | - (0) | -0.384 | + (1.173) | 0.168 | + (0.660) |
| P47 | 9 | MCS- | - (0.51) | + (0.59) | 8 | MCS- | - (0) | -0.247 | + (35.672) | 2.639 | + (0.736) |
| P48 | 11 | MCS- | - (0.54) | + (0.66) | NA | NA | NA | NA | NA | NA | NA |
| P49 | 3 | PTCS | - (0.54) | + (0.81) | NA | NA | NA | NA | NA | NA | NA |
| Abbreviations: CRS-R *Coma Recovery Scale-*Revised; DMN *default more network*; DRS *Disability Rating Scale*; EEG *electroencephalography*; fMRI *functional magnetic resonance imaging*; GOSE TBI/ALL *Glasgow Outcome Scale-Extended considering only the effects of all injuries [GOSE-All], and the effects of the TBI only [GOSE-TBI*]; N/A *not applicable*; SMA/PMC *supplementary motor area/premotor cortex*; STG *superior temporal gyrus* | | | | | | | | | | | |

| **Supplementary Table 10: Secondary Outcomes for Individual Participants** | | | | | | |
| --- | --- | --- | --- | --- | --- | --- |
| **ID** | **DRS**  **Total** | **DRS_Depend_**  **1=Died**  **2=Dependent**  **3=Independent** | **GOSE-ALL**  **Ordinal** | **GOSE-ALL**  **Dichotomous**  **1=Died**  **2=Unfavorable (≥3)**  **3=Unfavorable (≥4** | **GOSE-TBI**  **Ordinal** | **GOSE-TBI**  **Dichotomous**  **1=Died**  **2=Unfavorable (<4)**  **3=Unfavorable (≥4** |
| P1 | 30 | 1 | 1 | 1 | 1 | 1 |
| P2 | 5 | 3 | 4 | 3 | 4 | 3 |
| P3 | 30 | 1 | 1 | 1 | 1 | 1 |
| P4 | 30 | 1 | 1 | 1 | 1 | 1 |
| P5 | 12 | 2 | 3 | 2 | 3 | 2 |
| P6 | 30 | 1 | 1 | 1 | 1 | 1 |
| P7 | 30 | 1 | 1 | 1 | 1 | 1 |
| P8 | 2 | 3 | 7 | 3 | 8 | 3 |
| P9 | 6 | 3 | 3 | 2 | 8 | 3 |
| P10 | 2 | 3 | 6 | 3 | 7 | 3 |
| P11 | 14 | 2 | 3 | 2 | 3 | 2 |
| P12 | 16 | 2 | 3 | 2 | 3 | 2 |
| P13 | 9 | 2 | 4 | 3 | 4 | 3 |
| P14 | 11 | 2 | 3 | 2 | 3 | 2 |
| P15 | 30 | 1 | 1 | 1 | 1 | 1 |
| P16 | 0 | 3 | 8 | 3 | 8 | 3 |
| P17 | 30 | 1 | 1 | 1 | 1 | 1 |
| P18 | 0 | 3 | 6 | 3 | 8 | 3 |
| **Supplementary Table 10 Continued** | | | | | | |
| **ID** | **DRS**  **Total** | **DRS_Depend_**  **1=Died**  **2=Dependent**  **3=Independent** | **GOSE-ALL**  **Ordinal** | **GOSE-ALL**  **Dichotomous**  **1=Died**  **2=Unfavorable (≤3)**  **3=Favorable (≥4** | **GOSE-TBI**  **Ordinal** | **GOSE-TBI**  **Dichotomous**  **1=Died**  **2=Unfavorable (≤3)**  **3=Favorable (≥4** |
| P19 | 23 | 2 | 3 | 2 | 3 | 2 |
| P20 | 30 | 1 | 1 | 1 | 1 | 1 |
| P21 | 22 | 2 | 2 | 2 | 2 | 2 |
| P22 | 1 | 3 | 8 | 3 | 8 | 3 |
| P23 | 23 | 2 | 2 | 2 | 2 | 2 |
| P24 | 4 | 3 | 5 | 3 | 5 | 3 |
| P25 | 30 | 1 | 1 | 1 | 1 | 1 |
| P26 | 11 | 2 | 3 | 2 | 3 | 2 |
| P27 | 1 | 3 | 5 | 3 | 5 | 3 |
| P28 | 30 | 1 | 1 | 1 | 1 | 1 |
| P29 | 1 | 3 | 6 | 3 | 6 | 3 |
| P30 | 3 | 3 | 5 | 3 | 6 | 3 |
| P31 | 5 | 3 | 3 | 2 | 3 | 2 |
| P32 | 3 | 3 | 4 | 3 | 4 | 3 |
| P33 | 13 | 2 | 3 | 2 | 3 | 2 |
| P34 | 14 | 2 | 3 | 2 | 3 | 2 |

| **Supplementary Table 10 Continued** | | | | | | |
| --- | --- | --- | --- | --- | --- | --- |
| **ID** | **DRS**  **Total** | **DRS_Depend_**  **1=Died**  **2=Dependent**  **3=Independent** | **GOSE-ALL**  **Ordinal** | **GOSE-ALL**  **Dichotomous**  **1=Died**  **2=Unfavorable (≤3)**  **3=Favorable (≥4** | **GOSE-TBI**  **Ordinal** | **GOSE-TBI**  **Dichotomous**  **1=Died**  **2=Unfavorable (≤3)**  **3=Favorable (≥4** |
| P35 | 0 | 3 | 6 | 3 | 6 | 3 |
| P36 | 2 | 3 | 5 | 3 | 5 | 3 |
| P37 | 0 | 3 | 8 | 3 | 8 | 3 |
| P38 | 3 | 3 | 5 | 3 | 5 | 3 |
| P39 | 10 | 2 | 3 | 2 | 3 | 2 |
| P40 | 3 | 3 | 4 | 3 | 4 | 3 |
| P41 | 1 | 3 | 5 | 3 | 7 | 3 |
| P42 | 3 | 3 | 6 | 3 | 6 | 3 |
| P43 | 8 | 3 | NA | NA | NA | NA |
| P44 | 5 | 3 | 3 | 2 | 3 | 2 |
| P45 | 0 | 3 | 8 | 3 | 8 | 3 |
| Abbreviations*:* DRS *Disability Rating Scale*; GOSE *Glasgow Outcome Scale Extended* (GOSE-ALL indicated all injuries were considered in the scoring; GOSE-TBI *Glasgow Outcome Scale Extended score reflecting effects of the traumatic brain injury (TBI) only* | | | | | | |

| **Supplementary Table 11: Association Between CRS-R Diagnosis and 6mo Outcome** | | | | | | |
| --- | --- | --- | --- | --- | --- | --- |
|  | CRS-R Diagnosis – Best at Time of EEG or fMRI | | | | | p-value |
|  | Coma | VS/UWS | MCS- | MCS+ | eMCS |  |
| DRS Total 6 mo | 30 (5, 30) | 14 (0, 30) | 3 (0, 30) | 2.5 (0, 3) | 5 (0, 10) | 0.014 |
| DRS Total 6 mo alive | 8.5 (5, 12) | 7.5 (0, 23) | 3 (0, 15) | 2.5 (0, 3) | 5 (0, 10) | 0.282 |
| DRS_Depend_ Dichotomous | 1 (16.7) | 7 (36.8) | 5 (55.6) | 6 (100) | 4 (80) | 0.014 |
| DRS_Depend_ Dichotomous Alive | 1 (50) | 7 (50) | 5 (62.5) | 6 (100) | 4 (80) | 0.232 |
| GOSE-TBI | 1 (1, 4)6 | 3 (1, 8)19 | 4 (1, 6)9 | 5.5 (4, 8)6 | 3 (3, 8)4 | 0.06 |
| GOSE-TBI alive | 3.5 (3, 4)2 | 4.5 (2, 8)14 | 4.5 (3, 6)8 | 5.5 (4, 8)6 | 3 (3, 8)4 | 0.539 |
| GOSE-All | 1 (1, 4)6 | 3 (1, 8)19 | 4 (1, 6)9 | 5 (4, 8)6 | 3 (3, 8)4 | 0.04 |
| GOSE-All live | 3.5 (3, 4)2 | 3.5 (2, 8)14 | 4.5 (3, 6)8 | 5 (4, 8)6 | 3 (3, 8)4 | 0.512 |
| GOSE-TBI Dichotomous | 1 (16.7) | 8 (42.1) | 5 (55.6) | 6 (100) | 1 (25) | 0.032 |
| GOSE-TBI Alive Dichotomous | 1 (50) | 8 (57.1) | 5 (62.5) | 6 (100) | 1 (25) | 0.158 |
| GOSE-All Dichotomous | 1 (16.7) | 7 (36.8) | 5 (55.6) | 6 (100) | 1 (25) | 0.024 |
| GOSE-All Alive Dichotomous | 1 (50) | 7 (50) | 5 (62.5) | 6 (100) | 1 (25) | 0.113 |

Legend: p-value was calculated using the Kruskal-Wallis test

| **Supplementary Table 12: Association between Composite Measures of Command-following and Consciousness and 6mo Outcome** | | | | |
| --- | --- | --- | --- | --- |
|  | **DRS Total (n=45)** | **DRS Total Alive (n=35)** | **DRS_Depend_ (n=45)** | **DRS_Depend_ Alive (n=35)** |
| age | **0.396 [0.070, 0.648]; p=0.007** | **0.352 [0.023, 0.648]; p=0.038** | **0.97 [0.94, 1]; p=0.038** | **0.97 [0.94, 1.01]; p=0.186** |
| **CM_Command_ (n=45)** | | | | |
| CRS-R | -9 [-20, -1]; p=0.011 | -3 [-9, 1]; p=0.153 | **16.15 [1.85, 141.26]; p=0.012** | 8.46 [0.93, 76.86]; p=0.058 |
| CRS-R or EEG | -2 [-12, 2]; p=0.334 | -1 [-9, 2]; p=0.427 | 2.44 [0.7, 8.49]; p=0.159 | 2.75 [0.59, 12.85]; p=0.198 |
| CRS-R or fMRI | -7 [-19, 0]; p=0.067 | 0 [-6, 3]; p=0.829 | 3.47 [0.99, 12.09]; p=0.051 | 1.82 [0.44, 7.48]; p=0.406 |
| CRS-R or EEG or fMRI | -2 [-12, 2]; p=0.313 | 0 [-6, 3]; p=0.829 | 2.27 [0.69, 7.54]; p=0.179 | 1.82 [0.44, 7.48]; p=0.406 |
| **CM_Conscious_ (n=45)** | | | | |
| CRS-R | **-11 [-20, -2]; p=0.003** | -3 [-9, 1]; p=0.096 | **6.37 [1.71, 23.76]; p=0.006** | 3.75 [0.86, 16.4]; p=0.079 |
| CRS-R or EEG | **-9 [-19, 0]; p=0.030** | -3 [-11, 1]; p=0.156 | **4 [1.15, 13.86]; p=0.029** | 3.2 [0.75, 13.66]; p=0.116 |
| CRS-R or fMRI | **-10.67 [-20, -1]; p=0.013** | -1 [-9, 2]; p=0.432 | **4.09 [1.16, 14.43]; p=0.028** | 2.02 [0.46, 8.87]; p=0.35 |
| CRS-R or EEG or fMRI | **-8 [-19, 0]; p=0.037** | -1 [-9, 2]; p=0.432 | 3.4 [0.97, 11.91]; p=0.056 | 2.02 [0.46, 8.87]; p=0.35 |
|  | **GOSE-TBI (n=44)** | **GOSE-TBI Alive (n=34)** | **GOSE-All (n=44)** | **GOSE-All Alive (n=34)** |
| age | **-0.395 [-0.653, -0.098]; p=0.008** | **-0.35 [-0.626, -0.007]; p=0.042** | **-0.462 [-0.694, -0.153]; p=0.002** | **-0.483 [-0.715, -0.188]; p=0.004** |
| **CM_Command_ (n=45)** |  |  |  |  |
| CRS-R | 2 [0, 3]; p=0.064 | 0 [-1, 2]; p=0.561 | **2 [0, 3]; p=0.037** | 0.48 [-1, 2]; p=0.371 |
| CRS-R or EEG | 0 [-2, 2]; p=0.701 | 0 [-2, 1]; p=0.957 | 0 [-1, 2]; p=0.518 | 0 [-1, 2]; p=0.662 |
| CRS-R or fMRI | 2 [0, 3]; p=0.089 | 0 [-2, 1]; p=0.944 | 2 [0, 2]; p=0.049 | 0 [-1, 2]; p=0.645 |
| CRS-R or EEG or fMRI | 0 [-1, 2]; p=0.388 | 0 [-2, 1]; p=0.944 | 1 [0, 2]; p=0.259 | 0 [-1, 2]; p=0.645 |
| **CM_Conscious_ (n=45)** | | | | |
| CRS-R | 2 [0, 3]; p=0.062 | 0 [-2, 2]; p=0.874 | **2 [0, 3]; p=0.022** | 0 [-1, 2]; p=0.435 |
| CRS-R or EEG | 1 [-1, 2]; p=0.237 | 0 [-2, 1]; p=0.986 | 1 [0, 2]; p=0.098 | 0 [-1, 2]; p=0.471 |
| CRS-R or fMRI | 2 [0, 3]; p=0.067 | 0 [-2, 1]; p=0.88 | **2 [0, 3]; p=0.018** | 0 [-1, 2]; p=0.52 |
| CRS-R or EEG or fMRI | 1 [0, 3]; p=0.152 | 0 [-2, 1]; p=0.88 | **2 [0, 3]; p=0.049** | 0 [-1, 2]; p=0.52 |

| **Supplementary Table 12 Continued** | | | | |
| --- | --- | --- | --- | --- |
|  | **GOSE-TBI Dichotomous (n=44)** | **GOSE-TBI Dichotomous Alive (n=34)** | **GOSE-All Dichotomous (n=44)** | **GOSE-All Dichotomous Alive (n=34)** |
| age | **0.96 [0.93, 0.99]; p=0.019** | 0.97 [0.93, 1]; p=0.087 | **0.96 [0.93, 0.99]; p=0.011** | **0.96 [0.92, 1]; p=0.044** |
| **CM_Command_ (n=45)** | | | | |
| CRS-R | 3.33 [0.73, 15.17]; p=0.119 | 1.67 [0.34, 8.07]; p=0.526 | 3.77 [0.83, 17.22]; p=0.087 | 1.97 [0.41, 9.52]; p=0.397 |
| CRS-R or EEG | 1.71 [0.5, 5.92]; p=0.394 | 1.69 [0.39, 7.27]; p=0.483 | 1.99 [0.57, 6.9]; p=0.28 | 2.05 [0.48, 8.77]; p=0.335 |
| CRS-R or fMRI | 2.51 [0.73, 8.63]; p=0.143 | 1.28 [0.32, 5.13]; p=0.724 | 2.97 [0.85, 10.31]; p=0.087 | 1.63 [0.41, 6.46]; p=0.487 |
| CRS-R or EEG or fMRI | 1.71 [0.52, 5.67]; p=0.379 | 1.28 [0.32, 5.13]; p=0.724 | 2.04 [0.61, 6.82]; p=0.248 | 1.63 [0.41, 6.46]; p=0.487 |
| **CM_Conscious_ (n=45)** |  |  |  |  |
| CRS-R | 3.05 [0.88, 10.52]; p=0.078 | 1.56 [0.39, 6.25]; p=0.534 | **3.64 [1.04, 12.78]; p=0.043** | 2 [0.5, 8]; p=0.327 |
| CRS-R or EEG | 3.11 [0.91, 10.69]; p=0.072 | 2.33 [0.56, 9.64]; p=0.242 | **3.89 [1.1, 13.76]; p=0.035** | 3.11 [0.75, 12.96]; p=0.119 |
| CRS-R or fMRI | 3.25 [0.93, 11.4]; p=0.066 | 1.56 [0.36, 6.76]; p=0.55 | **4.2 [1.15, 15.37]; p=0.03** | 2.25 [0.52, 9.73]; p=0.278 |
| CRS-R or EEG or fMRI | 2.73 [0.78, 9.53]; p=0.116 | 1.56 [0.36, 6.76]; p=0.55 | 3.55 [0.97, 12.9]; p=0.055 | 2.25 [0.52, 9.73]; p=0.278 |
| For age, a Spearman’s correlation coefficient is reported. For the remaining, a Wilcoxon rank sum test is reported along with a difference in the medians, 95% CI, and p-value. p<0.05 values are bolded | | | | |

|  | **Supplementary Table 13: Association Between EEG Biomarkers and 6mo Outcome** | | | | | | | | |
| --- | --- | --- | --- | --- | --- | --- | --- | --- | --- |
|  |  | | **DRS Total** | | **DRS Total Alive** | | **DRS_Depend_** | | **DRS_Depend_ Alive** |
|  | **Age** | | **0.363 [0.058, 0.626]; p=0.015** | | 0.313 [-0.053, 0.623]; p=0.072 | | 0.97 [0.94, 1]; p=0.054 | | 0.98 [0.94, 1.02]; p=0.235 |
|  | **CRS-R Conscious** | | **-11 [-20, -2]; p=0.006** | | -3 [-9, 1]; p=0.136 | | 5.95 [1.59, 22.33]; p=0.008 | | 3.5 [0.8, 15.4]; p=0.097 |
|  | **Days to EEG** | | 0.100 [-0.195, 0.424]; p=0.517 | | 0.075 [-0.27, 0.418]; p=0.675 | | 0.99 [0.91, 1.08]; p=0.842 | | 1.02 [0.91, 1.15]; p=0.703 |
|  | **Active-imagery [yes/no]** | | 3 [-4, 11]; p=0.278 | | 1.53 [-7, 5]; p=0.614 | | 0.57 [0.12, 2.74]; p=0.480 | | 0.83 [0.12, 5.85]; p=0.855 |
|  | **Active-imagery**  **[% accuracy]** | | 0.143 [-0.15, 0.431]; p=0.359 | | 0.043 [-0.314, 0.402]; p=0.81 | | -- | | -- |
|  | **Passive-language [yes/no]^c^** | | 1 [-24, 21]; p=0.640 | | 3 [-1, 21]; p=0.198 | | -- | | -- |
|  | **Passive-language [% accuracy]** | | -0.080 [-0.386, 0.276]; p=0.614 | | 0.163 [-0.241, 0.562]; p=0.365 | | -- | | -- |
|  |  | | **GOSE-TBI** | | **GOSE-TBI Alive** | | **GOSE-All** | | **GOSE-All Alive** |
|  | **Age** | | -0.359 [-0.628, -0.038]; p=0.018 | | -0.303 [-0.586, 0.033]; p=0.087 | | -0.425 [-0.696, -0.084]; p=0.005 | | -0.437 [-0.716, -0.119]; p=0.011 |
|  | **CRS-R Conscious** | | 2 [0, 3]; p=0.091 | | 0 [-2, 1]; p=1 | | 2 [0, 2]; p=0.036 | | 0 [-1, 2]; p=0.577 |
|  | **Days to EEG** | | -0.123 [-0.394, 0.196]; p=0.432 | | -0.115 [-0.435, 0.201]; p=0.525 | | -0.103 [-0.403, 0.205]; p=0.51 | | -0.08 [-0.414, 0.222]; p=0.659 |
|  | **Active-imagery [yes/no]** | | -2 [-3, 0]; p=0.166 | | -1 [-3, 1]; p=0.299 | | -1 [-2, 0]; p=0.208 | | -1 [-2, 1]; p=0.406 |
|  | **Active-imagery**  **[% accuracy]** | | -0.164 [-0.438, 0.138]; p=0.298 | | -0.088 [-0.422, 0.274]; p=0.633 | | -0.15 [-0.443, 0.158]; p=0.342 | | -0.063 [-0.386, 0.29]; p=0.73 |
|  | **Passive-language [yes/no]^c^** | | -2 [-6, 3]; p=0.46141 | | -3 [-5, 1]; p=0.08732 | | -2 [-5, 2]; p=0.441 | | -3 [-5, 0]; p=0.06 |
|  | **Passive-language [% accuracy]** | | 0.036 [-0.304, 0.365]; p=0.822 | | -0.219 [-0.563, 0.189]; p=0.228 | | 0.018 [-0.309, 0.366]; p=0.911 | | -0.252 [-0.621, 0.161]; p=0.165 |
| **Supplementary Table 13 Continued** | | | | | | | | | |
|  | | **GOSE-TBI Dichotomous** | | **GOSE-TBI Dichotomous Alive** | | **GOSE-All Dichotomous** | | **GOSE-All Dichotomous Alive** | |
| **Age** | | **0.96 [0.93, 1]; p=0.028** | | 0.97 [0.93, 1.01]; p=0.115 | | **0.96 [0.93, 0.99]; p=0.016** | | 0.96 [0.92, 1]; p=0.059 | |
| **CRS-R Conscious** | | 2.79 [0.8, 9.76]; p=0.107 | | 1.43 [0.35, 5.79]; p=0.62 | | 3.34 [0.94, 11.85]; p=0.062 | | 1.83 [0.45, 7.41]; p=0.395 | |
| **Days to EEG** | | 0.97 [0.89, 1.06]; p=0.543 | | 0.99 [0.89, 1.1]; p=0.849 | | 0.98 [0.89, 1.07]; p=0.595 | | 0.99 [0.89, 1.11]; p=0.905 | |
| **Active-imagery [yes/no]** | | 0.68 [0.14, 3.28]; p=0.626 | | 1.03 [0.15, 7.23]; p=0.975 | | 0.76 [0.16, 3.7]; p=0.734 | | 1.2 [0.17, 8.38]; p=0.854 | |
| **Active-imagery**  **[% accuracy]** | | -- | | -- | | -- | | -- | |
| **Passive-language [yes/no]^c^** | | -- | | -- | | -- | | -- | |
| **Passive-language [% accuracy]** | | -- | | -- | | -- | | -- | |
| Due to small cell counts for some combinations of predictors and outcomes (i.e., all but 3 participants responded to passive-language EEG) the statistical model could not be fit for some tests. p<0.05 values are bolded | | | | | | | | | |

| **Supplementary Table 14: Association Between fMRI Biomarkers and 6mo Outcome** | | | | |
| --- | --- | --- | --- | --- |
|  | **DRS Total** | **DRS Total Alive** | **DRS_Depend_** | **DRS_Depend_ Alive** |
| **Age** | **0.389 [0.028, 0.705]; p=0.031** | **0.420 [0.027, 0.737]; p=0.033** | 0.96 [0.93, 1]; p=0.06 | 0.96 [0.92, 1.01]; p=0.089 |
| **CRS-R Conscious** | **-9.07 [-20, -2]; p=0.010** | -4 [-12, 1]; p=0.084 | **5.14 [1.03, 25.6]; p=0.046** | 3 [0.56, 16.01]; p=0.199 |
| **Days to fMRI** | 0.159 [-0.217, 0.528]; p=0.392 | 0.188 [-0.204, 0.552]; p=0.358 | 0.99 [0.9, 1.09]; p=0.792 | 0.99 [0.89, 1.1]; p=0.856 |
| **Active-imagery [yes/no]** | 2 [-7, 10]; p=0.494 | 3 [-2, 10]; p=0.236 | 0.42 [0.08, 2.19]; p=0.301 | 0.29 [0.05, 1.78]; p=0.18 |
| **Active-imagery [% suprathreshold]** | 0.150 [-0.2, 0.459]; p=0.428 | 0.272 [-0.111, 0.61]; p=0.189 | 0.98 [0.42, 2.29]; p=0.968 | 0.88 [0.37, 2.07]; p=0.765 |
| **Active-imagery [ROI z-stat]** | 0.125 [-0.21, 0.483]; p=0.511 | 0.116 [-0.255, 0.494]; p=0.580 | 0.32 [0.03, 3.19]; p=0.332 | 0.32 [0.03, 3.96]; p=0.376 |
| **Passive-language [yes/no]^c^** | -3.80 [-19, 5]; p=0.549 | -6 [-19, 3]; p=0.433 | 2.8 [0.43, 18.38]; p=0.283 | 3.5 [0.46, 26.62]; p=0.226 |
| **Passive-language [% suprathreshold]** | -0.334 [-0.634, 0.032]; p=0.071 | -0.233 [-0.622, 0.258]; p=0.262 | 1.07 [0.99, 1.15]; p=0.073 | 1.05 [0.97, 1.13]; p=0.212 |
| **Passive-language [ROI z-stat]** | -0.241 [-0.562, 0.137]; p=0.20 | -0.236 [-0.601, 0.237]; p=0.257 | 1.81 [0.83, 3.95]; p=0.135 | 1.67 [0.72, 3.87]; p=0.234 |
| **Resting-state DMN [yes/no]** | -6 [-16, 1]; p=0.148 | -3 [-9, 2]; p=0.274 | **5.14 [1.03, 25.6]; p=0.046** | 5.14 [0.82, 32.3]; p=0.081 |
| **Resting-state DMN [z-stat]** | -0.332 [-0.624, 0.025]; p=0.068 | -0.276 [-0.63, 0.149]; p=0.172 | **32.4 [1.06, 990.42]; p=0.046** | 21.36 [0.52, 880.2]; p=0.107 |

| **Supplementary Table 14 Continued** | | | | |
| --- | --- | --- | --- | --- |
|  | **GOSE-TBI** | **GOSE-TBI Alive** | **GOSE-All** | **GOSE-All Alive** |
| **Age** | -0.389 [-0.687, 0.003]; p=0.03 | -0.424 [-0.733, -0.036]; p=0.031 | -0.454 [-0.754, -0.082]; p=0.01 | -0.518 [-0.806, -0.127]; p=0.007 |
| **CRS-R Conscious** | 1.63 [0, 3]; p=0.104 | 0 [-1, 2]; p=0.618 | 2 [0, 3]; p=0.041 | 1 [-1, 2]; p=0.303 |
| **Days to fMRI** | -0.199 [-0.506, 0.14]; p=0.283 | -0.251 [-0.59, 0.108]; p=0.216 | -0.162 [-0.512, 0.19]; p=0.384 | -0.195 [-0.542, 0.17]; p=0.339 |
| **Active-imagery [yes/no]** | 0 [-2, 2]; p=0.868 | 0 [-3, 1]; p=0.6 | 0 [-2, 2]; p=0.981 | 0 [-2, 1]; p=0.734 |
| **Active-imagery [% suprathreshold]** | -0.047 [-0.382, 0.298]; p=0.804 | -0.126 [-0.482, 0.25]; p=0.548 | -0.026 [-0.338, 0.311]; p=0.89 | -0.097 [-0.444, 0.294]; p=0.645 |
| **Active-imagery [ROI z-stat]** | -0.107 [-0.463, 0.272]; p=0.573 | -0.097 [-0.489, 0.274]; p=0.644 | -0.086 [-0.411, 0.282]; p=0.65 | -0.062 [-0.431, 0.297]; p=0.769 |
| **Passive-language [yes/no]^c^** | 1 [-2, 3]; p=0.511 | 1 [-3, 3]; p=0.387 | 1 [-2, 3]; p=0.563 | 1 [-3, 3]; p=0.445 |
| **Passive-language [% suprathreshold]** | 0.357 [-0.043, 0.668]; p=0.053 | 0.277 [-0.167, 0.663]; p=0.18 | **0.366 [0.01, 0.67]; p=0.047** | 0.283 [-0.224, 0.687]; p=0.171 |
| **Passive-language [ROI z-stat]** | 0.2 [-0.186, 0.549]; p=0.29 | 0.179 [-0.249, 0.603]; p=0.393 | 0.273 [-0.101, 0.607]; p=0.145 | 0.279 [-0.178, 0.684]; p=0.177 |
| **Resting-state DMN [yes/no]** | **2 [0, 4]; p=0.019** | **2 [0, 4]; p=0.024** | 2 [0, 3]; p=0.058 | 1 [0, 3]; p=0.093 |
| **Resting-state DMN [z-stat]** | **0.436 [0.140, 0.713]; p=0.014** | **0.429 [0.085, 0.715]; p=0.029** | 0.352 [0.036, 0.64]; p=0.052 | 0.307 [-0.085, 0.616]; p=0.127 |

| **Supplementary Table 14 Continued** | | | | |
| --- | --- | --- | --- | --- |
|  | **GOSE Dichotomous TBI** | **GOSE Dichotomous TBI Alive** | **GOSE Dichotomous all** | **GOSE Dichotomous all Alive** |
| **Age** | 0.96 [0.93, 1]; p=0.058 | 0.96 [0.92, 1.01]; p=0.085 | **0.96 [0.92, 1]; p=0.032** | **0.96 [0.91, 1]; p=0.044** |
| **CRS-R Conscious** | 2.75 [0.61, 12.41]; p=0.188 | 1.5 [0.3, 7.43]; p=0.619 | 3.43 [0.75, 15.67]; p=0.112 | 2 [0.41, 9.84]; p=0.394 |
| **Days to fMRI** | 0.97 [0.88, 1.07]; p=0.581 | 0.97 [0.87, 1.08]; p=0.612 | 0.98 [0.89, 1.08]; p=0.715 | 0.98 [0.89, 1.09]; p=0.765 |
| **Active-imagery [yes/no]** | 0.83 [0.16, 4.21]; p=0.825 | 0.67 [0.11, 3.99]; p=0.657 | 1 [0.2, 5.04]; p=1 | 0.85 [0.14, 4.99]; p=0.856 |
| **Active-imagery [% suprathreshold]** | 1.14 [0.47, 2.74]; p=0.772 | 1.02 [0.42, 2.51]; p=0.962 | 1.2 [0.5, 2.91]; p=0.685 | 1.09 [0.44, 2.69]; p=0.852 |
| **Active-imagery [ROI z-stat]** | 0.68 [0.08, 6.13]; p=0.732 | 0.79 [0.07, 8.71]; p=0.846 | 0.79 [0.09, 7.03]; p=0.834 | 0.93 [0.09, 9.78]; p=0.953 |
| **Passive-language [yes/no]^c^** | 2.8 [0.43, 18.38]; p=0.283 | 3.5 [0.46, 26.62]; p=0.226 | 2.36 [0.36, 15.45]; p=0.369 | 2.79 [0.37, 20.82]; p=0.318 |
| **Passive-language [% suprathreshold]** | 1.06 [0.99, 1.14]; p=0.095 | 1.04 [0.97, 1.11]; p=0.277 | 1.07 [0.99, 1.14]; p=0.072 | 1.05 [0.98, 1.12]; p=0.203 |
| **Passive-language [ROI z-stat]** | 1.69 [0.79, 3.62]; p=0.176 | 1.54 [0.68, 3.48]; p=0.301 | 1.96 [0.89, 4.32]; p=0.096 | 1.81 [0.78, 4.21]; p=0.166 |
| **Resting-state DMN [yes/no]** | **5.14 [1.03, 25.6]; p=0.046** | 5.14 [0.82, 32.3]; p=0.081 | 3.43 [0.75, 15.67]; p=0.112 | 3.05 [0.57, 16.19]; p=0.191 |
| **Resting-state DMN [z-stat]** | 9.39 [0.48, 185.14]; p=0.141 | 5.05 [0.21, 124.34]; p=0.322 | 3.96 [0.25, 61.95]; p=0.327 | 1.96 [0.1, 37.19]; p=0.654 |
| p<0.05 values are bolded | | | | |

| **Supplementary Table 15: Association Between EEG Biomarkers and 6mo Outcome in Subsample of Participants with Coma, VS/UWS, and MCS**- | | | | |
| --- | --- | --- | --- | --- |
|  | **DRS Total** | **DRS Total Alive** | **DRS_Depend_** | **DRS_Depend_ Alive** |
| **Age** | **0.369 [0.012, 0.665]; p=0.029** | 0.31 [-0.09, 0.658]; p=0.132 | 0.97 [0.93, 1]; p=0.066 | 0.97 [0.93, 1.02]; p=0.25 |
| **CRS-R Conscious** | -9 [-19, 0]; p=0.067 | -2 [-10, 2]; p=0.335 | 3.19 [0.7, 14.56]; p=0.135 | 2 [0.37, 10.92]; p=0.423 |
| **Days to EEG** | 0.174 [-0.189, 0.496]; p=0.318 | 0.137 [-0.249, 0.497]; p=0.515 | 0.98 [0.89, 1.08]; p=0.741 | 1.01 [0.9, 1.14]; p=0.861 |
| **Active-imagery [yes/no]** | 5 [-4, 18]; p=0.272 | 2 [-11, 9]; p=0.793 | 0.27 [0.03, 2.59]; p=0.255 | 0.38 [0.03, 4.81]; p=0.451 |
| **Active-imagery**  **[% accuracy]** | 0.126 [-0.245, 0.455]; p=0.478 | -0.015 [-0.421, 0.421]; p=0.945 | 0 [0, 958.62]; p=0.153 | 0 [0, 578391.15]; p=0.259 |
| **Passive-language [yes/no]^c^** | 2 [-19, 28]; p=0.569 | 4 [-2, 21]; p=0.209 | 0.33 [0.03, 4.1]; p=0.391 | 0 [0, Inf]; p=0.995 |
| **Passive-language [% accuracy]** | -0.162 [-0.495, 0.214]; p=0.367 | 0.019 [-0.44, 0.408]; p=0.93 | 84.7 [0.03, 228555.73]; p=0.271 | 6.29 [0, 44753.81]; p=0.684 |
|  | **GOSE-TBI** | **GOSE-TBI Alive** | **GOSE-All** | **GOSE-All Alive** |
| **Age** | -0.333 [-0.646, 0.036]; p=0.05 | -0.23 [-0.606, 0.163]; p=0.268 | -0.39 [-0.693, -0.051]; p=0.021 | -0.372 [-0.702, 0.002]; p=0.067 |
| **CRS-R Conscious** | 1 [-1, 2]; p=0.341 | 0 [-2, 1]; p=0.726 | 1 [0, 2]; p=0.203 | 0 [-2, 2]; p=0.93 |
| **Days to EEG** | -0.161 [-0.456, 0.153]; p=0.357 | -0.113 [-0.437, 0.239]; p=0.59 | -0.155 [-0.476, 0.185]; p=0.375 | -0.104 [-0.435, 0.271]; p=0.621 |
| **Active-imagery [yes/no]** | -1 [-4, 1]; p=0.247 | 0 [-4, 1]; p=0.719 | -1 [-2, 1]; p=0.296 | 0 [-3, 1]; p=0.891 |
| **Active-imagery**  **[% accuracy]** | -0.11 [-0.448, 0.259]; p=0.537 | 0.053 [-0.345, 0.429]; p=0.805 | -0.104 [-0.442, 0.258]; p=0.557 | 0.066 [-0.365, 0.487]; p=0.76 |
| **Passive-language [yes/no]^c^** | -2 [-6, 2]; p=0.425 | -4 [-5, 1]; p=0.097 | -2 [-5, 2]; p=0.353 | -3 [-5, 0]; p=0.059 |
| **Passive-language [% accuracy]** | 0.112 [-0.27, 0.481]; p=0.535 | -0.105 [-0.519, 0.379]; p=0.626 | 0.07 [-0.29, 0.446]; p=0.698 | -0.188 [-0.606, 0.278]; p=0.38 |

| **Supplementary Table 15 Continued** | | | | |
| --- | --- | --- | --- | --- |
|  | **GOSE-TBI Dichotomous** | **GOSE-TBI Dichotomous Alive** | **GOSE-All Dichotomous** | **GOSE-All Dichotomous Alive** |
| **Age** | 0.97 [0.93, 1]; p=0.064 | 0.97 [0.93, 1.02]; p=0.241 | **0.96 [0.92, 1]; p=0.037** | 0.96 [0.92, 1.01]; p=0.118 |
| **CRS-R Conscious** | 1.78 [0.4, 7.84]; p=0.447 | 0.97 [0.19, 5.03]; p=0.973 | 2.12 [0.48, 9.5]; p=0.324 | 1.25 [0.24, 6.44]; p=0.79 |
| **Days to EEG** | 0.98 [0.89, 1.08]; p=0.706 | 1.01 [0.9, 1.13]; p=0.907 | 0.99 [0.89, 1.09]; p=0.763 | 1.01 [0.9, 1.13]; p=0.862 |
| **Active-imagery [yes/no]** | 0.77 [0.12, 4.96]; p=0.786 | 1.82 [0.14, 23.25]; p=0.646 | 0.9 [0.14, 5.81]; p=0.912 | 2.2 [0.17, 28.14]; p=0.544 |
| **Active-imagery**  **[% accuracy]** | -- | -- | -- | -- |
| **Passive-language [yes/no]^c^** | -- | -- | -- | -- |
| **Passive-language [% accuracy]** | -- | -- | -- | -- |
| Due to small cell counts for some combinations of predictors and outcomes (i.e., all but 3 participants responded to passive-language EEG) the statistical model could not be fit for some tests, as indicated by “--”. p<0.05 values are bolded | | | | |

| **Supplementary Table 16: Association Between fMRI Biomarkers and 6mo Outcome in Subsample of Participants with Coma, VS/UWS, and MCS-** | | | | |
| --- | --- | --- | --- | --- |
|  | **DRS Total** | **DRS Total Alive** | **DRS_Depend_** | **DRS_Depend_ Alive** |
| **Age** | 0.306 [-0.139, 0.687]; p=0.146 | 0.323 [-0.197, 0.723]; p=0.178 | 0.96 [0.91, 1.01]; p=0.09 | 0.96 [0.9, 1.01]; p=0.119 |
| **CRS-R Conscious** | -9 [-20, 2]; p=0.124 | -2 [-12, 5]; p=0.404 | 2.57 [0.34, 19.33]; p=0.359 | 1.5 [0.19, 11.93]; p=0.702 |
| **Days to fMRI** | 0.186 [-0.212, 0.564]; p=0.384 | 0.243 [-0.23, 0.675]; p=0.316 | 0.98 [0.88, 1.09]; p=0.706 | 0.98 [0.87, 1.1]; p=0.754 |
| **Active-imagery [yes/no]** | 0 [-13, 9]; p=0.902 | 1 [-7, 9]; p=0.672 | 0.77 [0.14, 4.39]; p=0.77 | 0.54 [0.08, 3.53]; p=0.517 |
| **Active-imagery [% suprathreshold]** | -0.001 [-0.401, 0.391]; p=0.995 | 0.151 [-0.324, 0.573]; p=0.538 | 1.19 [0.5, 2.82]; p=0.697 | 1.05 [0.44, 2.51]; p=0.917 |
| **Active-imagery [ROI z-stat]** | 0.155 [-0.230, 0.524]; p=0.469 | 0.129 [-0.321, 0.552]; p=0.598 | 0.25 [0.02, 2.88]; p=0.264 | 0.27 [0.02, 3.47]; p=0.316 |
| **Passive-language [yes/no]^c^** | 0 [-17, 8]; p=0.947 | -2 [-17, 5]; p=0.711 | 1.6 [0.23, 11.08]; p=0.634 | 2 [0.25, 15.99]; p=0.513 |
| **Passive-language [% suprathreshold]** | -0.232 [-0.606, 0.187]; p=0.275 | -0.172 [-0.617, 0.345]; p=0.481 | 1.06 [0.98, 1.14]; p=0.126 | 1.04 [0.96, 1.12]; p=0.314 |
| **Passive-language [ROI z-stat]** | -0.109 [-0.512, 0.313]; p=0.611 | -0.129 [-0.589, 0.328]; p=0.598 | 1.47 [0.65, 3.31]; p=0.354 | 1.36 [0.58, 3.19]; p=0.475 |
| **Resting-state DMN [yes/no]** | -9 [-20, 1]; p=0.093 | -7 [-16, 2]; p=0.205 | **8.56 [1.33, 54.95]; p=0.024** | **8.17 [1.03, 64.94]; p=0.047** |
| **Resting-state DMN [z-stat]** | **-0.411 [-0.707, -0.027]; p=0.046** | -0.394 [-0.737, 0.071]; p=0.095 | **48.45 [1.12, 2102.48]; p=0.044** | 29.57 [0.56, 1562.1]; p=0.094 |

| **Supplementary Table 16 Continued** | | | | |
| --- | --- | --- | --- | --- |
|  | **GOSE-TBI** | **GOSE-TBI Alive** | **GOSE-All** | **GOSE-All Alive** |
| **Age** | -0.249 [-0.652, 0.199]; p=0.241 | -0.232 [-0.672, 0.236]; p=0.339 | -0.327 [-0.735, 0.174]; p=0.119 | -0.363 [-0.752, 0.127]; p=0.127 |
| **CRS-R Conscious** | 1 [-2, 3]; p=0.387 | 0 [-2, 2]; p=1 | 1 [-1, 3]; p=0.261 | 0 [-2, 2]; p=0.775 |
| **Days to fMRI** | -0.176 [-0.523, 0.217]; p=0.412 | -0.228 [-0.627, 0.255]; p=0.348 | -0.146 [-0.518, 0.23]; p=0.495 | -0.18 [-0.6, 0.28]; p=0.46 |
| **Active-imagery [yes/no]** | 0 [-2, 2]; p=0.686 | 0 [-2, 2]; p=0.966 | 1 [-2, 2]; p=0.512 | 0 [-2, 2]; p=0.828 |
| **Active-imagery [% suprathreshold]** | 0.071 [-0.319, 0.459]; p=0.74 | -0.041 [-0.437, 0.409]; p=0.866 | 0.117 [-0.292, 0.478]; p=0.587 | 0.028 [-0.393, 0.457]; p=0.909 |
| **Active-imagery [ROI z-stat]** | -0.124 [-0.493, 0.275]; p=0.562 | -0.088 [-0.505, 0.324]; p=0.721 | -0.109 [-0.492, 0.299]; p=0.614 | -0.052 [-0.485, 0.418]; p=0.831 |
| **Passive-language [yes/no]^c^** | 0 [-3, 3]; p=0.787 | 1 [-3, 3]; p=0.508 | 0 [-3, 2]; p=0.892 | 1 [-3, 2]; p=0.633 |
| **Passive-language [% suprathreshold]** | 0.275 [-0.135, 0.653]; p=0.193 | 0.255 [-0.301, 0.725]; p=0.293 | 0.272 [-0.204, 0.689]; p=0.198 | 0.239 [-0.35, 0.755]; p=0.325 |
| **Passive-language [ROI z-stat]** | 0.099 [-0.356, 0.546]; p=0.644 | 0.115 [-0.426, 0.641]; p=0.64 | 0.178 [-0.291, 0.611]; p=0.405 | 0.236 [-0.296, 0.731]; p=0.331 |
| **Resting-state DMN [yes/no]** | **2 [0, 4]; p=0.028** | **2 [0, 4]; p=0.045** | 2 [0, 3]; p=0.069 | 1 [0, 3]; p=0.141 |
| **Resting-state DMN [z-stat]** | **0.483 [0.139, 0.74]; p=0.017** | **0.507 [0.085, 0.785]; p=0.027** | 0.403 [0.043, 0.695]; p=0.051 | 0.383 [-0.071, 0.717]; p=0.106 |

| **Supplementary Table 16 Continued** | | | | |
| --- | --- | --- | --- | --- |
|  | **GOSE Dichotomous TBI** | **GOSE Dichotomous TBI Alive** | **GOSE Dichotomous all** | **GOSE Dichotomous all Alive** |
| **Age** | 0.97 [0.93, 1.02]; p=0.196 | 0.97 [0.92, 1.02]; p=0.296 | 0.96 [0.92, 1.01]; p=0.108 | 0.96 [0.91, 1.01]; p=0.147 |
| **CRS-R Conscious** | 2.06 [0.28, 15.36]; p=0.48 | 1.12 [0.14, 8.99]; p=0.912 | 2.57 [0.34, 19.33]; p=0.359 | 1.5 [0.19, 11.93]; p=0.702 |
| **Days to fMRI** | 0.96 [0.86, 1.07]; p=0.44 | 0.95 [0.85, 1.08]; p=0.451 | 0.97 [0.87, 1.08]; p=0.559 | 0.97 [0.86, 1.09]; p=0.588 |
| **Active-imagery [yes/no]** | 1.29 [0.23, 7.05]; p=0.772 | 0.95 [0.14, 6.28]; p=0.96 | 1.67 [0.3, 9.27]; p=0.56 | 1.33 [0.2, 8.71]; p=0.764 |
| **Active-imagery [% suprathreshold]** | 1.29 [0.52, 3.21]; p=0.578 | 1.13 [0.45, 2.85]; p=0.793 | 1.39 [0.55, 3.5]; p=0.483 | 1.23 [0.48, 3.13]; p=0.662 |
| **Active-imagery [ROI z-stat]** | 0.73 [0.08, 6.94]; p=0.786 | 0.89 [0.08, 9.74]; p=0.921 | 0.84 [0.09, 8.14]; p=0.884 | 1.02 [0.1, 10.96]; p=0.984 |
| **Passive-language [yes/no]^c^** | 2 [0.29, 13.81]; p=0.482 | 2.7 [0.33, 21.98]; p=0.353 | 1.6 [0.23, 11.08]; p=0.634 | 2 [0.25, 15.99]; p=0.513 |
| **Passive-language [% suprathreshold]** | 1.06 [0.99, 1.14]; p=0.117 | 1.04 [0.96, 1.12]; p=0.305 | 1.07 [0.99, 1.15]; p=0.088 | 1.05 [0.97, 1.13]; p=0.225 |
| **Passive-language [ROI z-stat]** | 1.55 [0.68, 3.52]; p=0.3 | 1.45 [0.6, 3.5]; p=0.41 | 1.82 [0.77, 4.26]; p=0.171 | 1.72 [0.7, 4.26]; p=0.239 |
| **Resting-state DMN [yes/no]** | 5.83 [0.98, 34.64]; p=0.052 | 5.25 [0.7, 39.48]; p=0.107 | 3.75 [0.67, 20.86]; p=0.131 | 3 [0.46, 19.59]; p=0.251 |
| **Resting-state DMN [z-stat]** | 10.75 [0.46, 253.47]; p=0.141 | 5.48 [0.19, 158.88]; p=0.322 | 4.28 [0.23, 78.63]; p=0.327 | 2.04 [0.09, 44.62]; p=0.652 |

| **Supplementary Table 17: Clinical interventions, data sharing, and goals of care** | | | | | | | | |
| --- | --- | --- | --- | --- | --- | --- | --- | --- |
| **ID** | **Day post injury of EEG/fMRI** | **Day Trach/Peg** | **If EEG/fMRI after Trach/PEG, Reason Why** | **If research EEG/fMRI not collected at the same time as clinical EEG/MRI, Reason Why** | **Day Post-injury MRI Results Available** | **Goals of Care At MRI** | **Goals of Care Before MRI Data Shared** | **Goals of Care After MRI Data Shared** |
| P1 | 17/NA | 2/NA | Trach was placed at an outside hospital prior to transfer | NA | NA | NA | NA | NA |
| P2 | 4/5 | NA/NA | NA | Research EEG collected at the same time as clinical EEG; Clinical MRI was ordered prior to obtaining consent for Research MRI. | 6 | Full Code | Full Code | Full Code |
| P3 | 5/NA | NA | NA | NA | NA | NA | NA | NA |
| P4 | 8/9 | NA | NA | NA- collected at same time as clinical | 10 | Full Code | Full Code | CMO |
| P5 | 2/9 | 12 | NA | Research EEG collected at the same time as clinical EEG; Clinical MRI completed day before research MRI because subject was enrolled in another study requiring MRI, which was collected with the clinical MRI making the scan time too long for our research scan | 10 | Full Code | Full Code | Full Code |
| P6 | 3/NA | NA | No trach/peg | NA | NA | NA | NA | NA |
| P7 | 26/25 | NA^1^ | Trach was placed at another facility prior to transfer | Clinical EEG and MRI completed prior to enrollment | 30 | Full Code | CMO | CMO |
| P8 | 5/5 | 8 | NA | Clinical EEG completed prior to enrollment | 11 | Full Code | Full Code | Full Code |
|  | 6/6 | 7^2^ | NA | NA | 12 | Full Code | Full Code | Full Code |
| **Supplementary Table 17 continued** | | | | | | | | |
| **ID** | **Day post injury of EEG/fMRI** | **Day Trach/Peg** | **If EEG/fMRI after Trach/PEG, Reason Why** | **If research EEG/fMRI not collected at the same time as clinical EEG/MRI, Reason Why** | **Day Post-injury MRI Results Available** | **Goals of Care At MRI** | **Goals of Care Before MRI Data Shared** | **Goals of Care After MRI Data Shared** |
| P10 | 4/3 | 8 | NA | Clinical EEG completed prior to enrollment | 1 | Full Code | Full Code | Full Code |
| P11 | 12/11 | 21 | NA | Research EEG obtained at the same time as clinical EEG; Clinical MRI completed on Day 1, prior to obtaining consent | 11 (within 24 hours of MRI) | Full Code | Full Code | Full Code |
| P12 | 2/9 | 14 | NA | Research EEG collected at the same time as clinical/Clinical MRI obtained without knowledge of research team | 9 (within 24 hours of MRI) | Full Code | Full Code | Full Code |
| P13 | 2/2 | NA/10 | NA | NA | 7 | Full Code | Full Code | Full Code |
| P14 | 14/NA | Placed at outside facility | Placed at outside facility | NA | No MRI | NA | NA | NA |
| P15 | 3/4 | NA | NA | NA | 5 | Full Code | Full Code | Full Code |
| P16 | 3/NA | 14/14 | NA | NA | NA | NA | NA | NA |
| P17 | 1/NA | NA | NA | NA | NA | NA | NA | NA |
| P18 | 2/NA | NA/14 | NA | Clinical EEG and MRI completed prior to consent for research. Subsequent MRI for research was not obtained because pt was restless/agitated and clinical team said he would not tolerate. | NA | NA | NA | NA |
| P19 | 6/17 | 7 | Trach placed after EEG but before MRI due to medical instability for MRI | NA | Day of MRI | Full Code | Full Code | Full Code |
| **Supplementary Table 17 continued** | | | | | | | | |
| **ID** | **Day post injury of EEG/fMRI** | **Day Trach/Peg** | **If EEG/fMRI after Trach/PEG, Reason Why** | **If research EEG/fMRI not collected at the same time as clinical EEG/MRI, Reason Why** | **Day Post-injury MRI Results Available** | **Goals of Care At MRI** | **Goals of Care Before MRI Data Shared** | **Goals of Care After MRI Data Shared** |
| P20 | 3/4 | NA | NA | NA | 5 | Full Code | NA^a^ | NA^a^ |
| P21 | 6/16 | 35 | NA | NA | 17 | DNR | DNR | DNR |
| P22 | 5/8 | NA | No trach/peg | NA | 9 | Full Code | Full Code | Full Code |
| P23 | 5/4 | 8 | NA | NA | 5 | Full Code | Full Code | Full Code |
| P24 | 34/34 | 22/32 | Patient transferred to MGH ICU after trach/peg was placed | No clinical EEG ordered; MRI collected at same time as clinical MRI | 35 | Full Code | Full Code | Full Code |
| P25 | 11/11 | NA | No trach/peg | NA | NA- participant died the day after EEG/fMRI | NA | NA | NA |
| P26 | 2/2 | 14^3^ | NA | Research EEG collected at the same time as clinical EEG; Clinical MRI not ordered by attending physician | 3 | Full Code | Full Code | Full Code |
| P27 | 2/4 | NA/9 | N/A | Research EEG collected at the same time as clinical EEG; ICP elevation after EEG, leading to delay in MRI | 7 (called family but not available) | Full Code | DNR | DNR- switched to Full Code several months later |

| **Supplementary Table 17 continued** | | | | | | | | |
| --- | --- | --- | --- | --- | --- | --- | --- | --- |
| **ID** | **Day post injury of MRI/EEG** | **Day Trach/Peg** | **If EEG/fMRI after Trach/PEG, Reason Why** | **If research EEG/fMRI not collected at the same time as clinical EEG/MRI, Reason Why** | **Day Post-injury MRI Results Available** | **Goals of Care At MRI/EEG** | **Goals of Care Before MRI Data Shared** | **Goals of Care After MRI Data Shared** |
| P28 | 18/NA | 2/2 | Multiple surgeries and grafts prevented earlier EEG acquisition | Clinical EEG not ordered | NA | NA | NA | NA |
| P29 | 15/NA | 15/15 | NA | NA | NA | NA | NA | NA |
| P30 | 2/6 | NA/11 | NA | NA | 7 | Full Code | Full Code | Full Code |
| P31 | 3/8 | 9/15 | NA | Research EEG acquired at the same time as clinical, clinical MRI conducted over the weekend and staff unavailable to collect research data | 9 | Full Code | Full Code | Full Code |
| P32 | 6/10 | 11/2 | PEG before because undergoing abdominal surgery | NA | 7 | Full Code | Full Code | Full Code |
| P33 | 15/NA | 13/13 | No family available for several days after injury delaying consent | NA | NA | NA | NA | NA |
| P34 | 8/23 | 7/7 | Delayed acute enrollment | Research EEG acquired at the same time as clinical, clinical MRI obtained prior to enrollment | 9 | Full Code | Full Code | Full Code |
| P35 | 10/11 | NA | No Trach/Peg | NA | 12 | Full Code | Full Code | Full Code |
| P36 | 15/21 | 20/20 | EEG before trach/PEG but MRI after due to medical instability | Research EEG same time as clinical; Clinical MRI not indicated. | 22 | Full Code | Full Code | Full Code |
| P37 | 9/9 | NA | NA | NA | 7 | Full Code | Full Code | Full Code |
| **Supplementary Table 17 continued** | | | | | | | | |
| **ID** | **Day post injury of EEG/fMRI** | **Day Trach/Peg** | **If EEG/fMRI after Trach/PEG, Reason Why** | **If research EEG/fMRI not collected at the same time as clinical EEG/MRI, Reason Why** | **Day Post-injury MRI Results Available** | **Goals of Care At MRI** | **Goals of Care Before MRI Data Shared** | **Goals of Care After MRI Data Shared** |
| P38 | 6/NA | 1/7 | Trach on day 1 due to complex facial fractures | NA | NA | NA | NA | NA |
| P39 | 6/8 | NA | No trach/peg | NA | NA | NA | NA | NA |
| P40 | 4/15 | NA | No trach/peg | Research EEG conducted at the same time as clinical EEG; Clinical MRI was completed Day 8 with research MRI for another study; could not combine all MRI’s due to excessive scan time so study MRI was conducted at a later date | 16 | Full Code | Full Code | Full Code |
| P41 | 4/NA | NA | No trach/peg | NA | NA | NA | NA | NA |
| P42 | 5/5 | NA | No trach/peg | No clinical EEG ordered; MRI collected at same time as clinical MRI. | 6 | Full Code | Full Code | Full Code |
| P43 | 6/NA | NA | Extubated prior to EEG, no trach or PEG | Enrolled after clinical EEG was obtained. Clinical MRI obtained on day 4, prior to enrollment. Research MRI not obtained because pt moved to the floor | No MRI | NA | NA | NA |
| P44 | 17/NA | 15/15 | Self-extubated in the morning, had to be emergently re-intubated; significant agitation | Enrolled after clinical EEG was obtained | NA | NA | NA | NA |
| P45 | NA/3 | 3/NA | Extubated on day 3, the day before research consent was obtained | Clinical MRI was ordered prior to obtaining consent for study. Consent was obtained the same day of the clinical MRI, so both MRIs were completed on the same day (but different scanners) | 8 | Full Code | Full Code | Full Code |
| ^a^ Clinical team requested data not be shared until after family meeting; goals of care changed to Comfort Measures Only (CMO) at family meeting | | | | | | | | |

**Supplementary Figure 1**

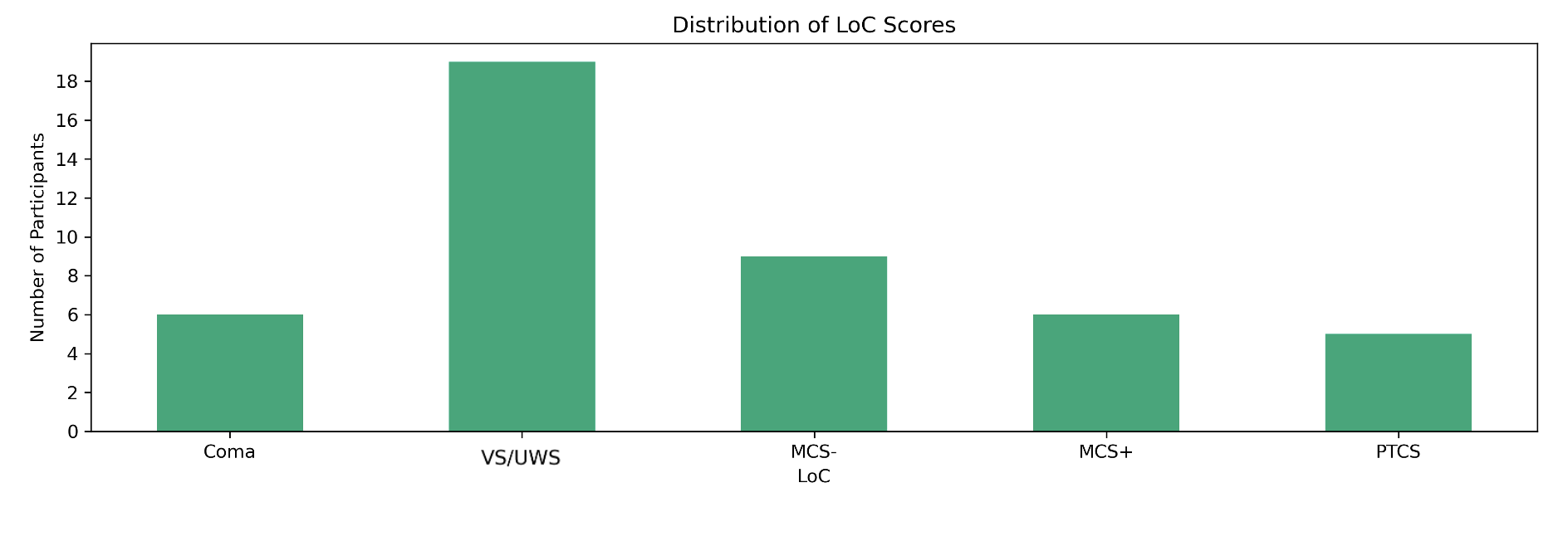

Highest DoC Diagnosis at Time of EEG or fMRI

Number of Participants

**Supplementary Figure 1: Distribution of CRS-R Diagnostic Ratings**

Abbreviations: DoC *disorders of* consciousness; EEG *electroencephalography;* fMRI *functional magnetic resonance imaging;* MCS-/MCS+ *minimally conscious state without/with language function*; PTCS *post traumatic confusional state;* VS *vegetative state*

**Supplementary Figure 2: Distribution of DRS Scores**

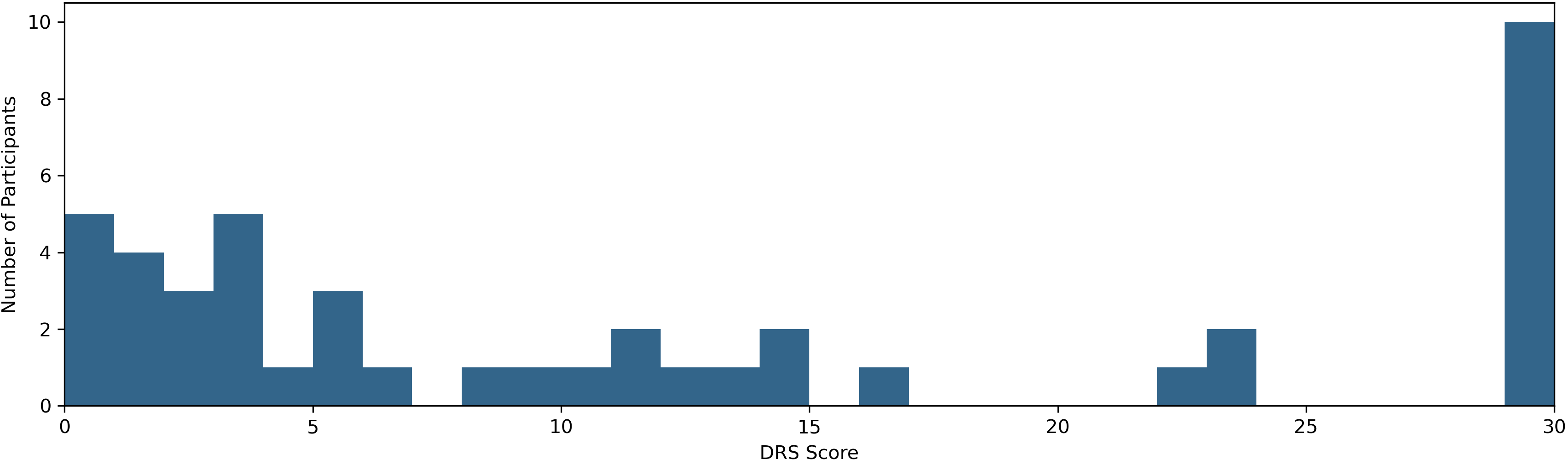

DRS Total

**Supplementary Figure 2: Distribution of DRS Scores**

Number of participants with each DRS total score at 6-months post-injury. A score of 0 indicates no disability, scores ≥12 suggest severe disability, and a score of 30 indicates death.

Abbreviations: DRS *Disability Rating Scale*

Number of Participants

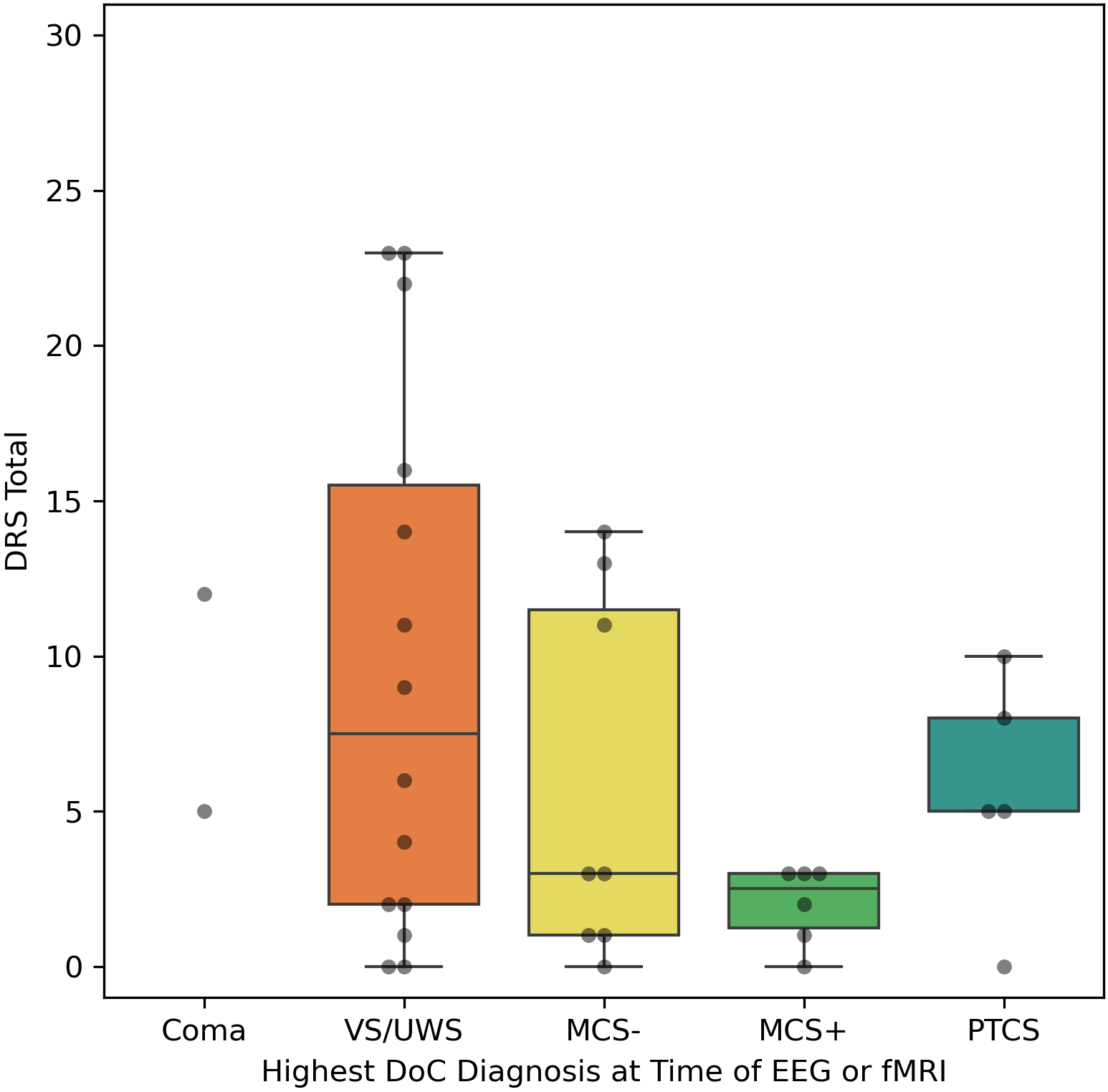

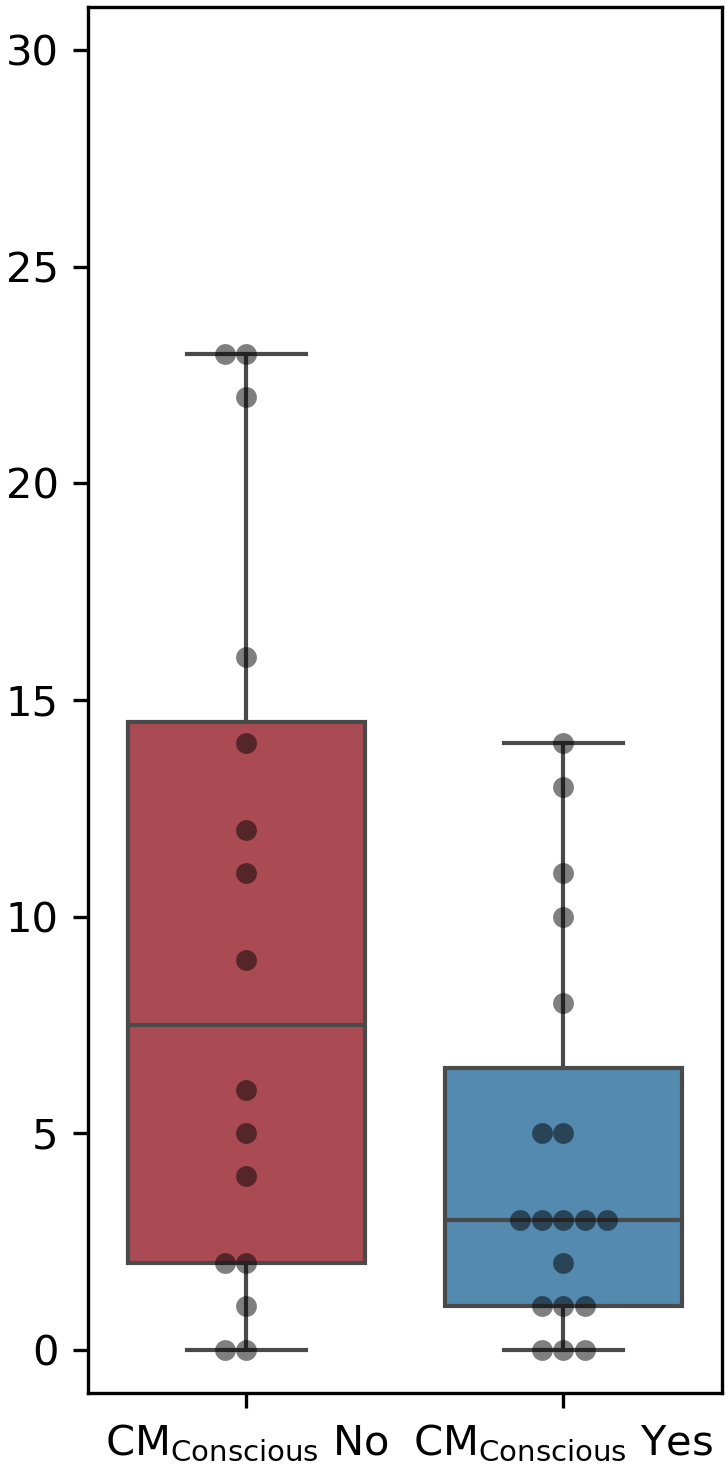

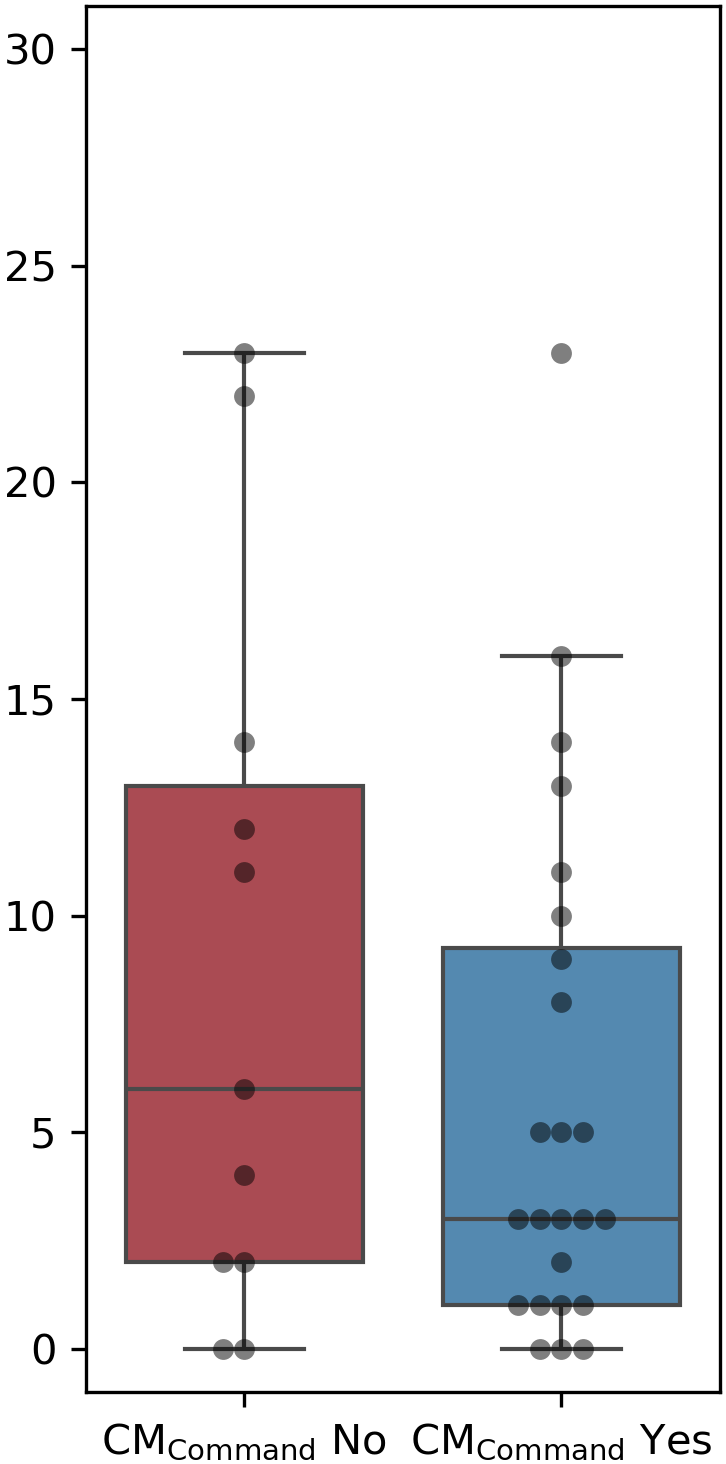

**Supplemental Figure 3**

B

**Supplementary Figure 3: DRS Outcome Across Behavioral Diagnosis and Composite Measures of Command-following and Consciousness in Participants who Survived**

In a subsample of participants who survived to six months, the 6-month DRS score is plotted for participants who had each behavioral diagnosis acutely, based on the Coma Recovery Scale-Revised (CRS-R, [A]) and for participants who had evidence of command-following (CM_Command_: CRS-R diagnosis of MCS+, or PTCS, or positive response to active-motor EEG, or positive response to active-motor fMRI, [B]), or evidence of consciousness (CM_Conscious_: CRS-R diagnosis of MCS-, or MCS+, or PTCS, or positive response to active-motor EEG, or positive response to active-motor fMRI [C]) on a composite measure. The behavioral diagnosis is based on the best CRS-R diagnosis obtained at EEG or fMRI. Medians are indicated by a solid line and means are indicated by a dotted line in each box plot. Abbreviations: CM_Conscious_ *composite measure of consciousness;* CM_Command_ *composite measure of command-following;* DoC *disorders of consciousness;* MCS-/MCS+ *minimally conscious state without/with language function*; VS *vegetative state;* PTCS *post traumatic confusional state*

C

A

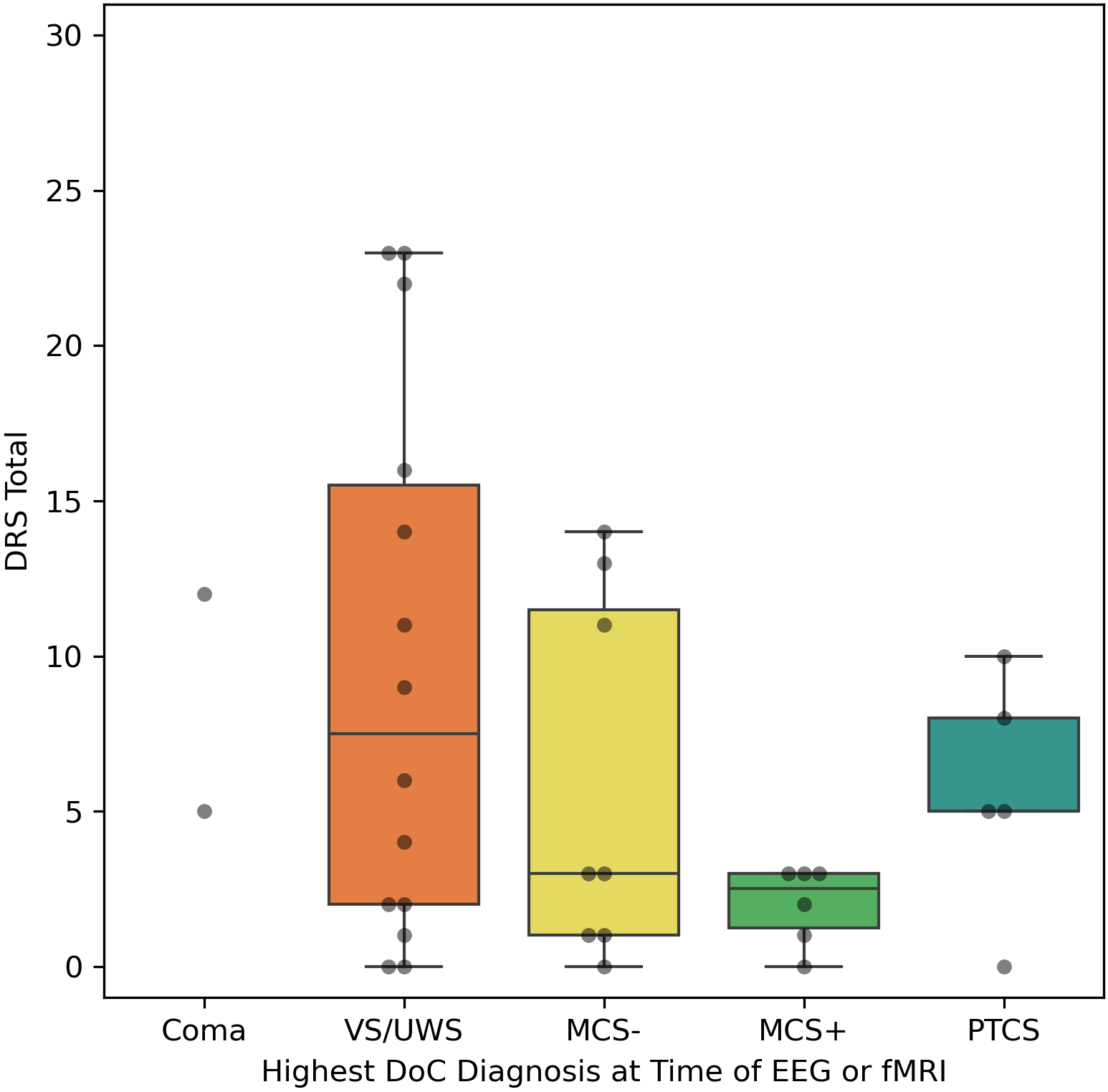

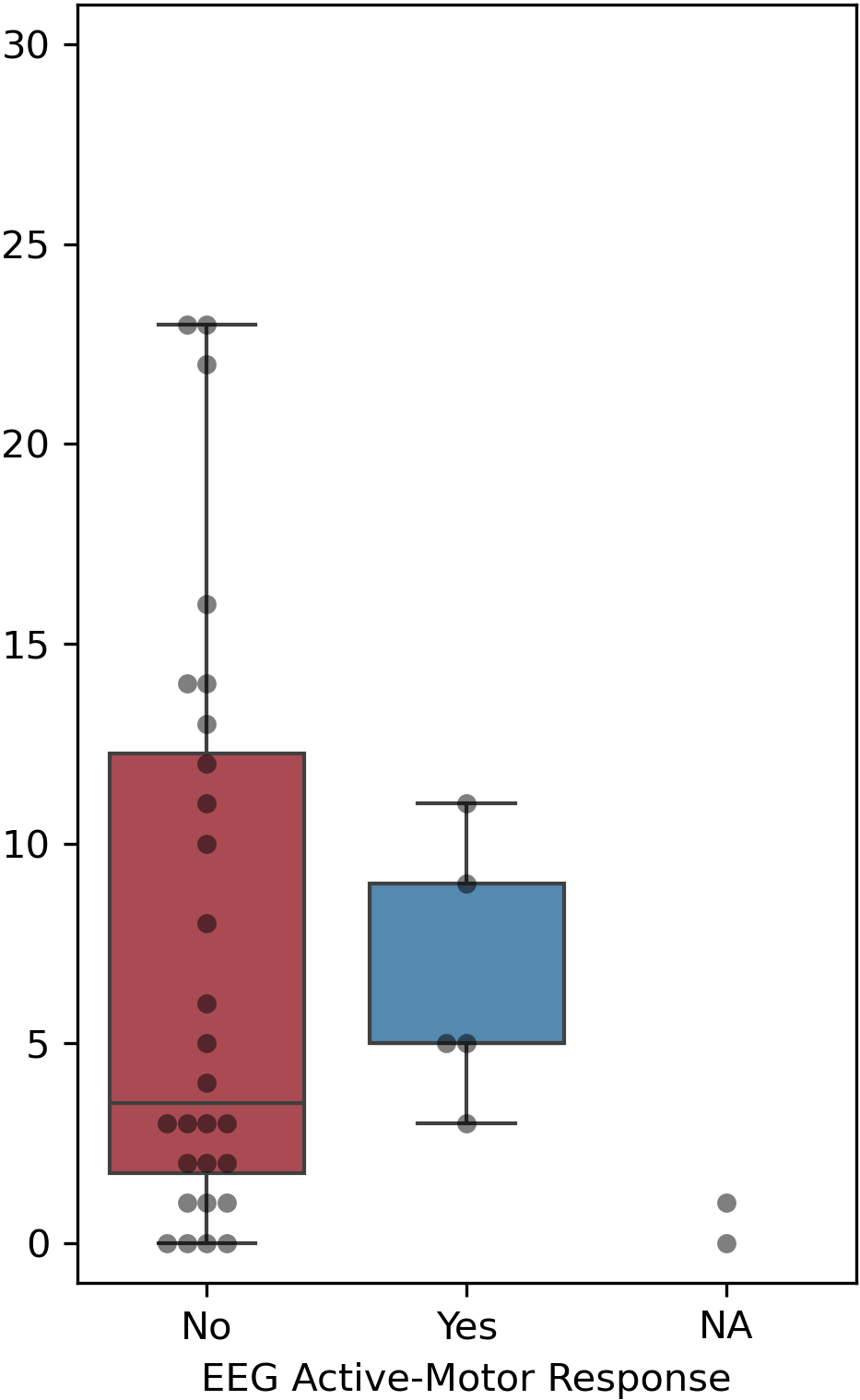

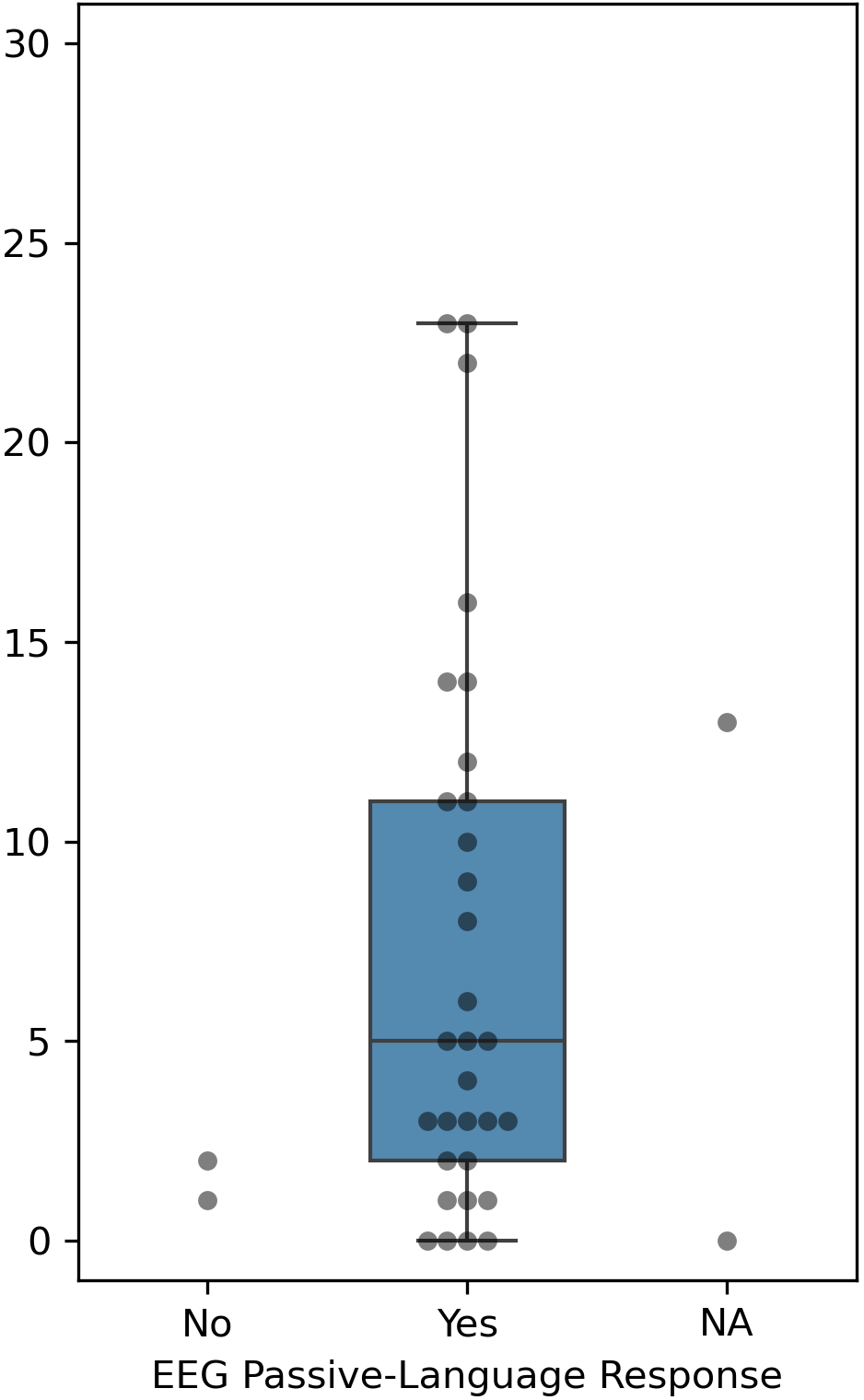

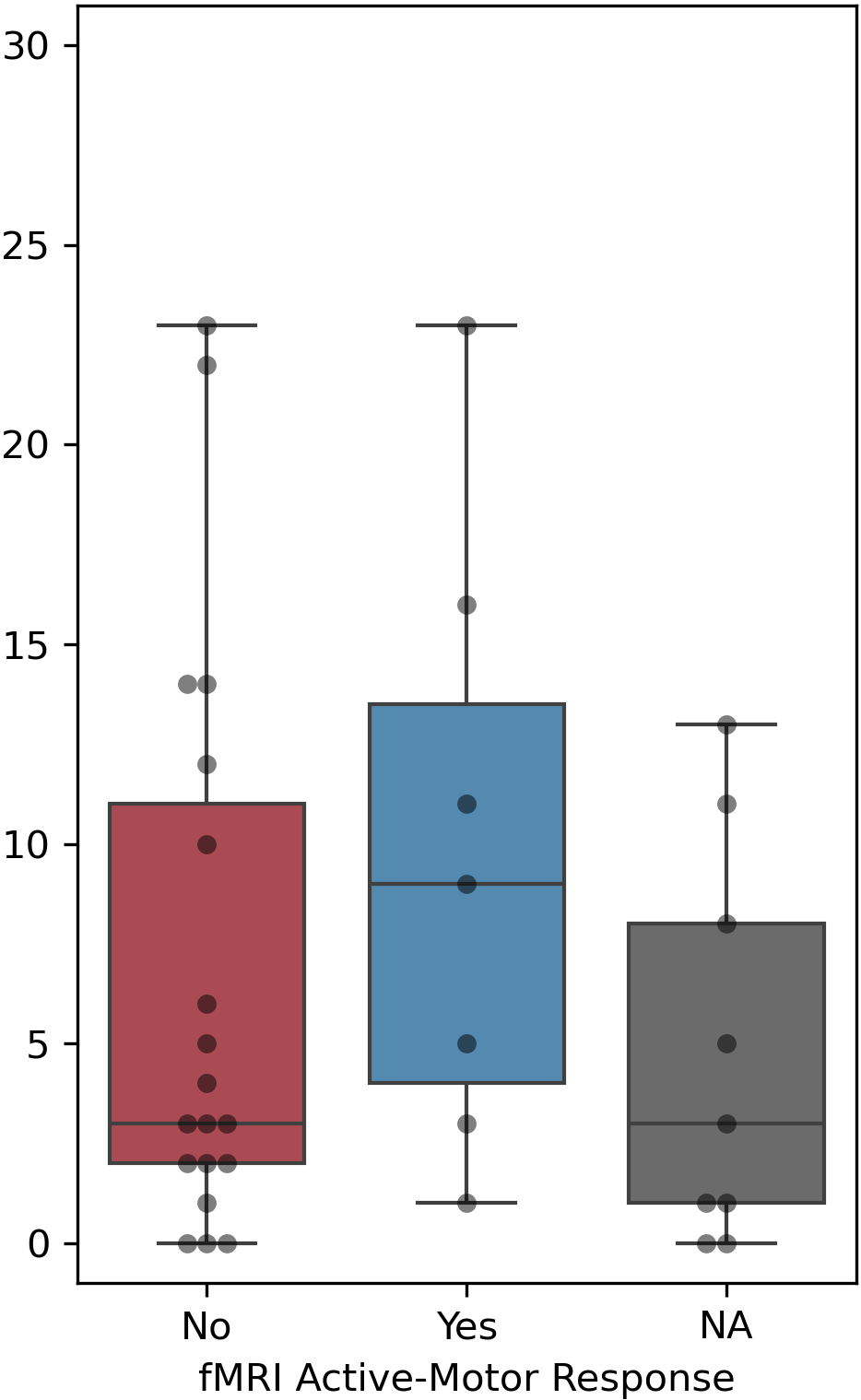

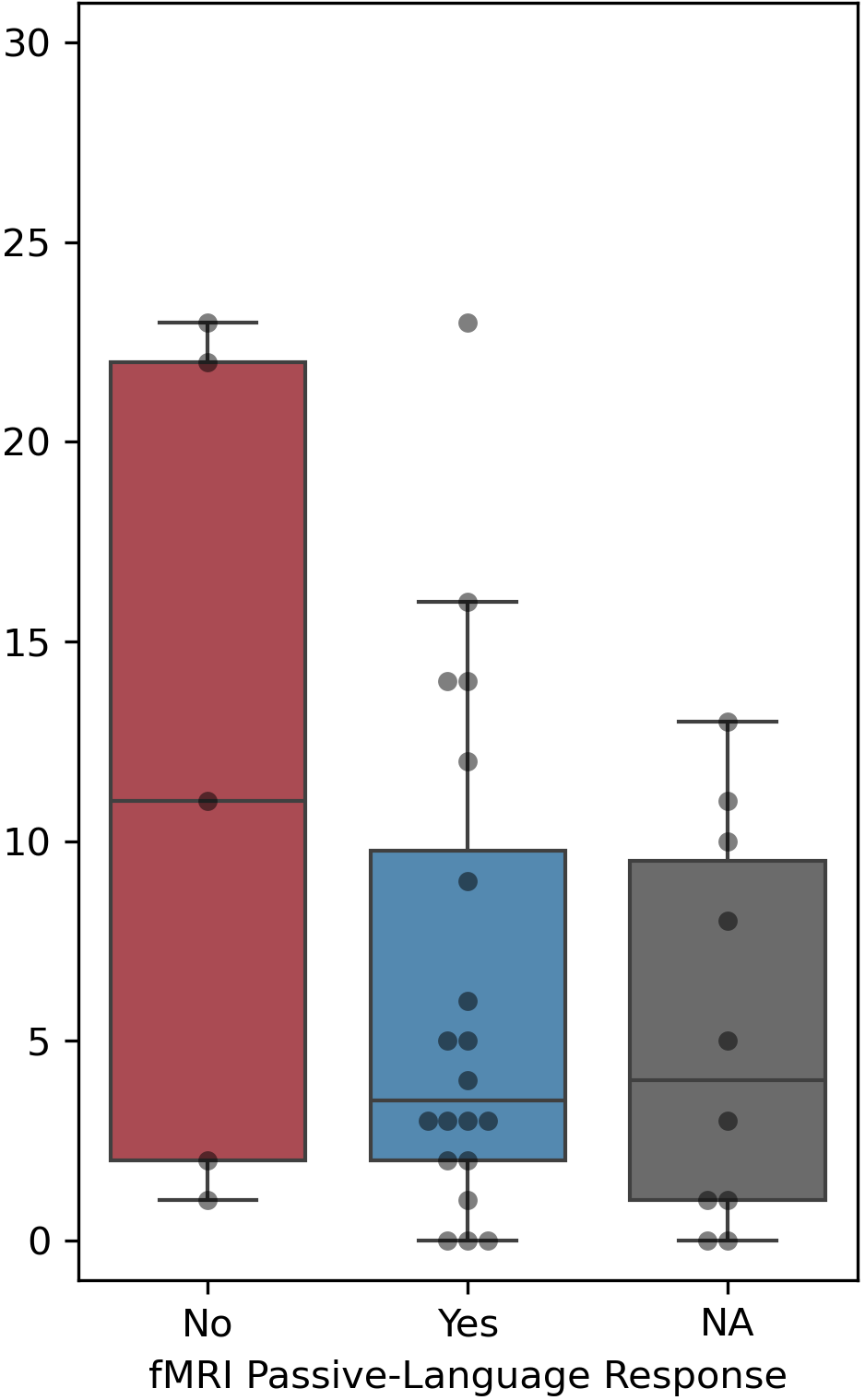

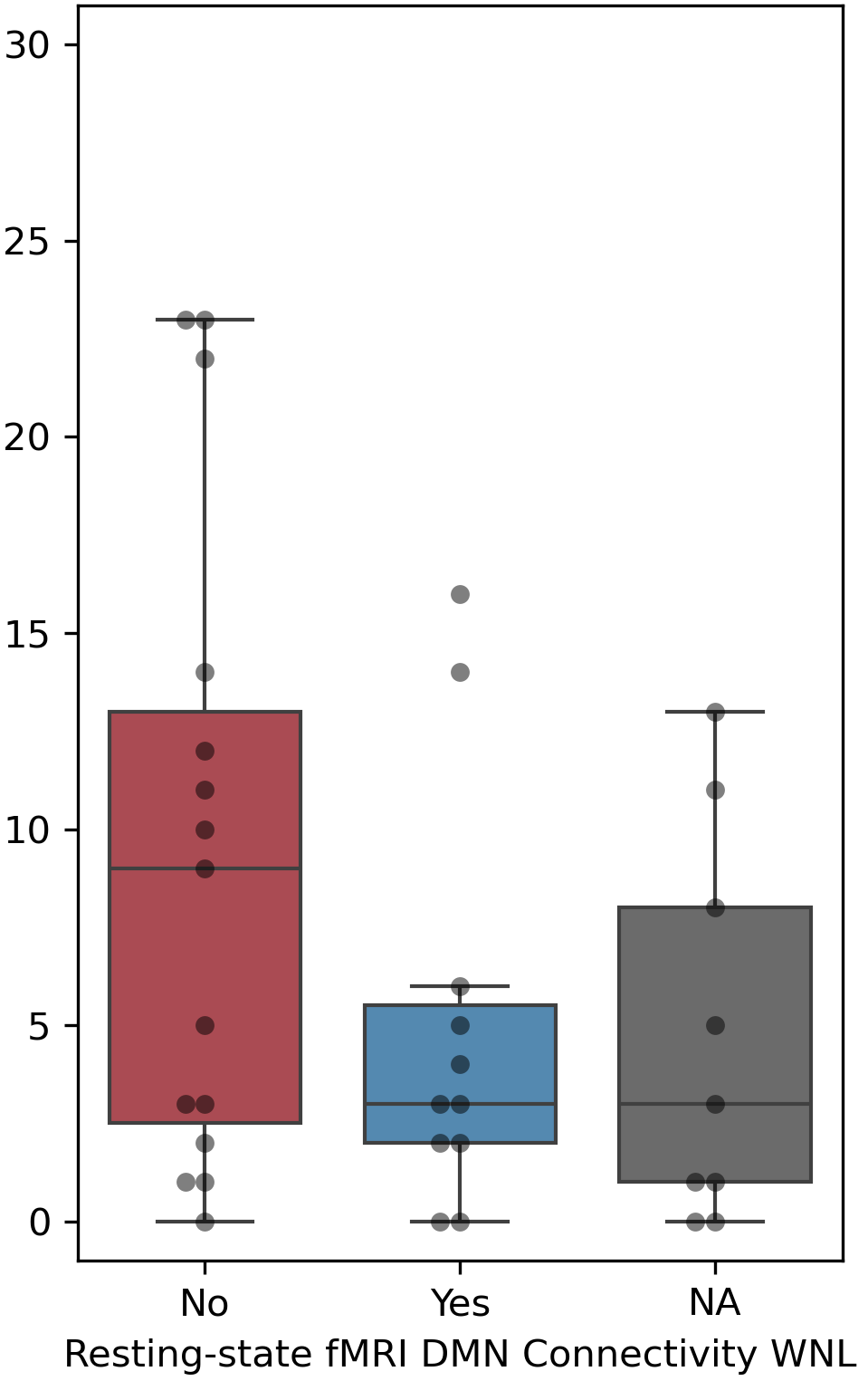

**Supplementary Figure 4**

E

C

A

D
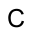

B
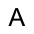

**Supplementary Figure 4: DRS Outcome Across Responses to EEG and fMRI in Participants who Survived**

In a subsample of participants who survived to six months, the 6-month DRS score is plotted based on the presence of a response to active-motor EEG (A), passive-language EEG (B), active-motor fMRI (C), passive-language fMRI (D), and intact DMN connectivity (E). Box plots are omitted when there are less than 6 data points in a group. NA indicates that the data were not acquired (see Main Manuscript, Table 1). Abbreviations: DoC *disorders of consciousness;* MCS-/MCS+ *minimally conscious state without/with language function*; NA *not acquired;* PTCS *post-traumatic confusional state;* VS/UWS *vegetative state/unresponsive wakefulness syndrome;* WNL *within normal limits (i.e., within 95% confidence interval of healthy control participants)*

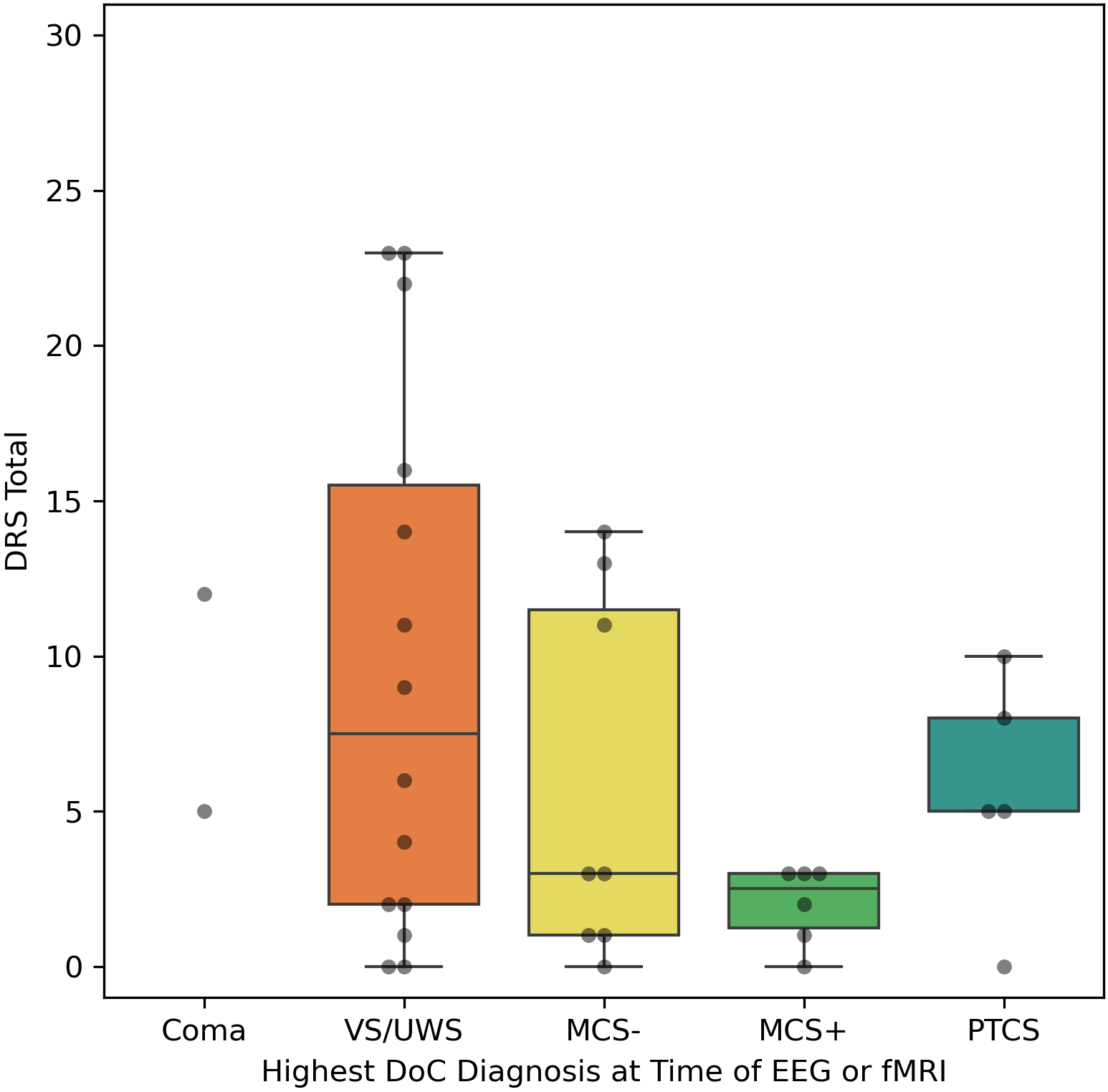

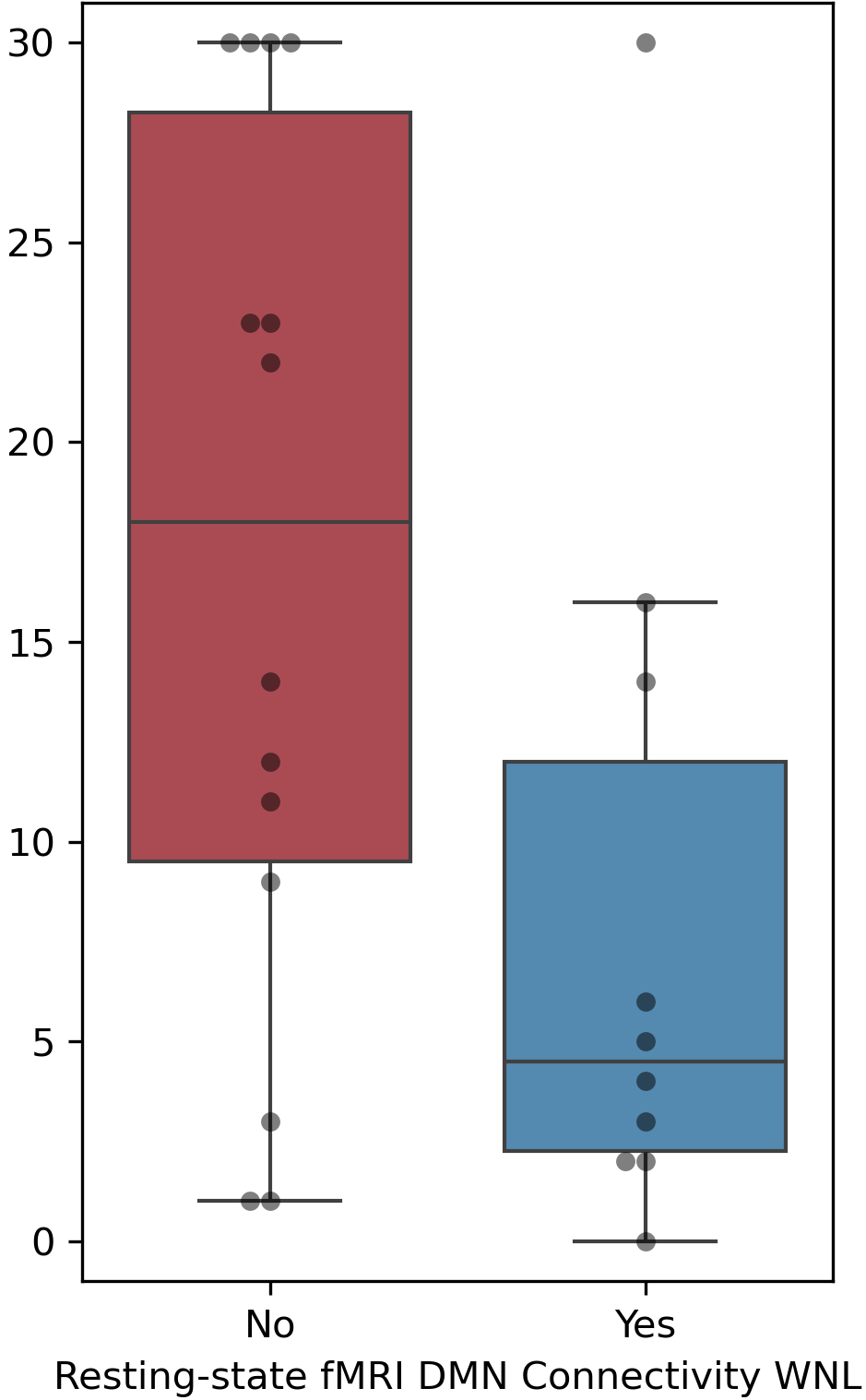

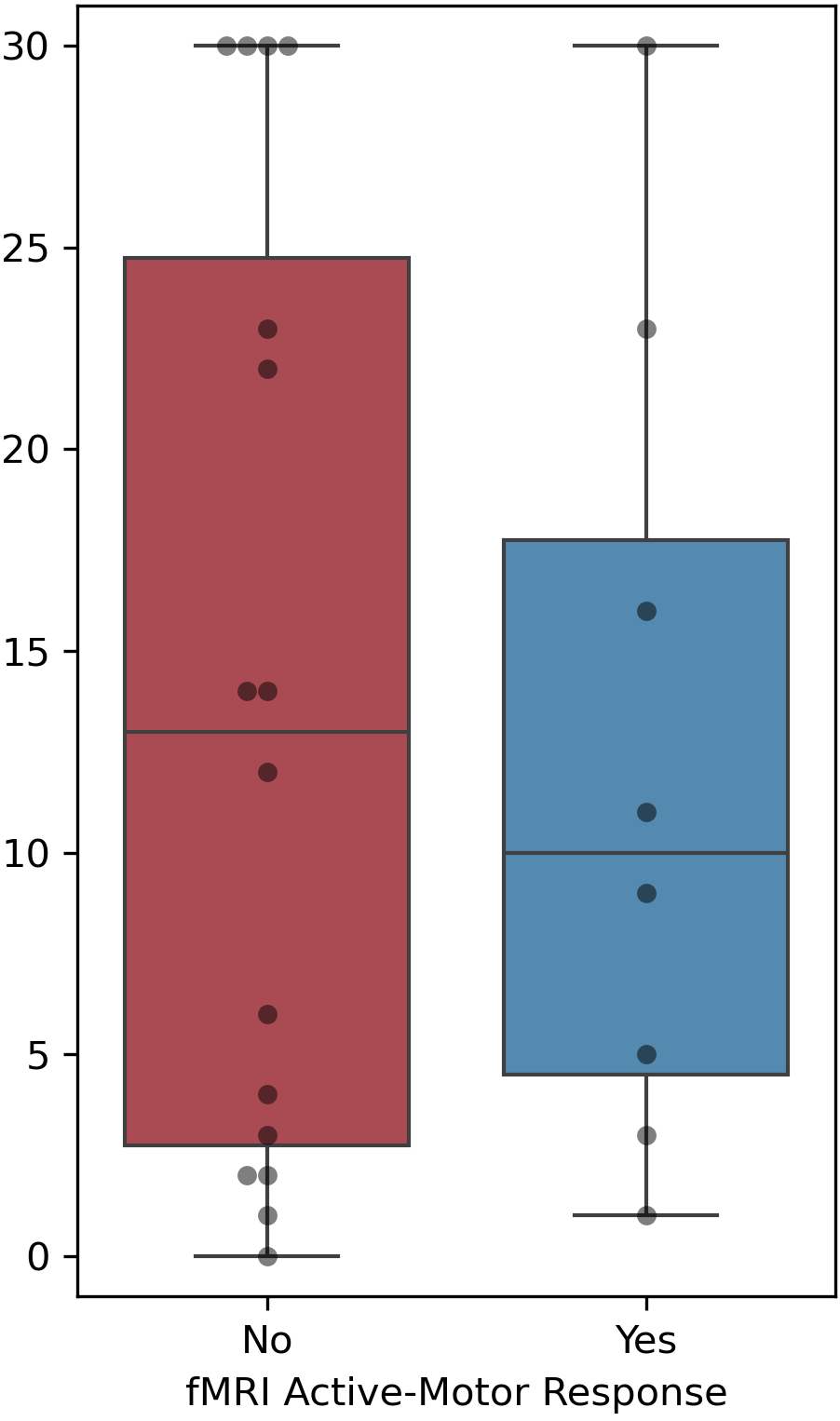

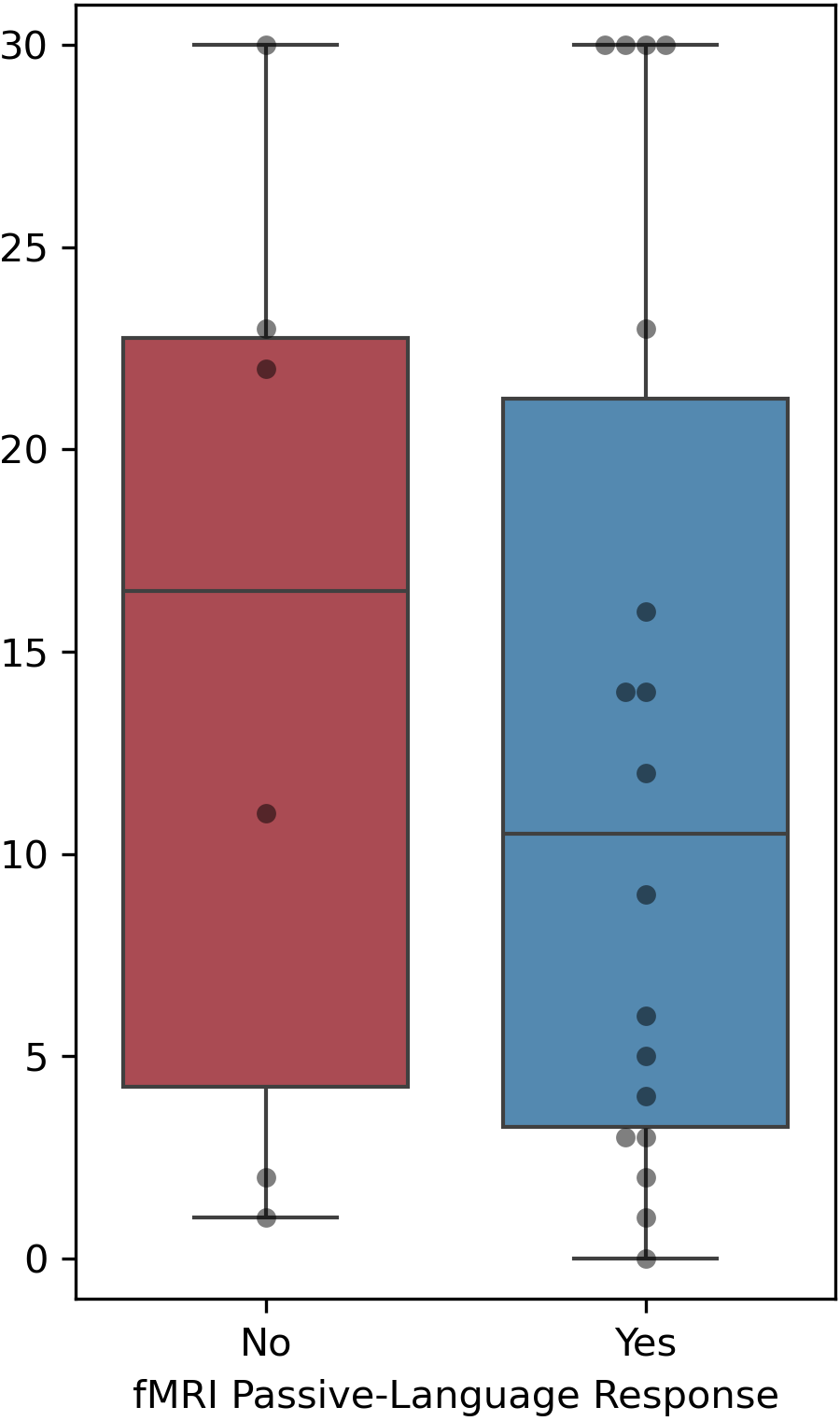

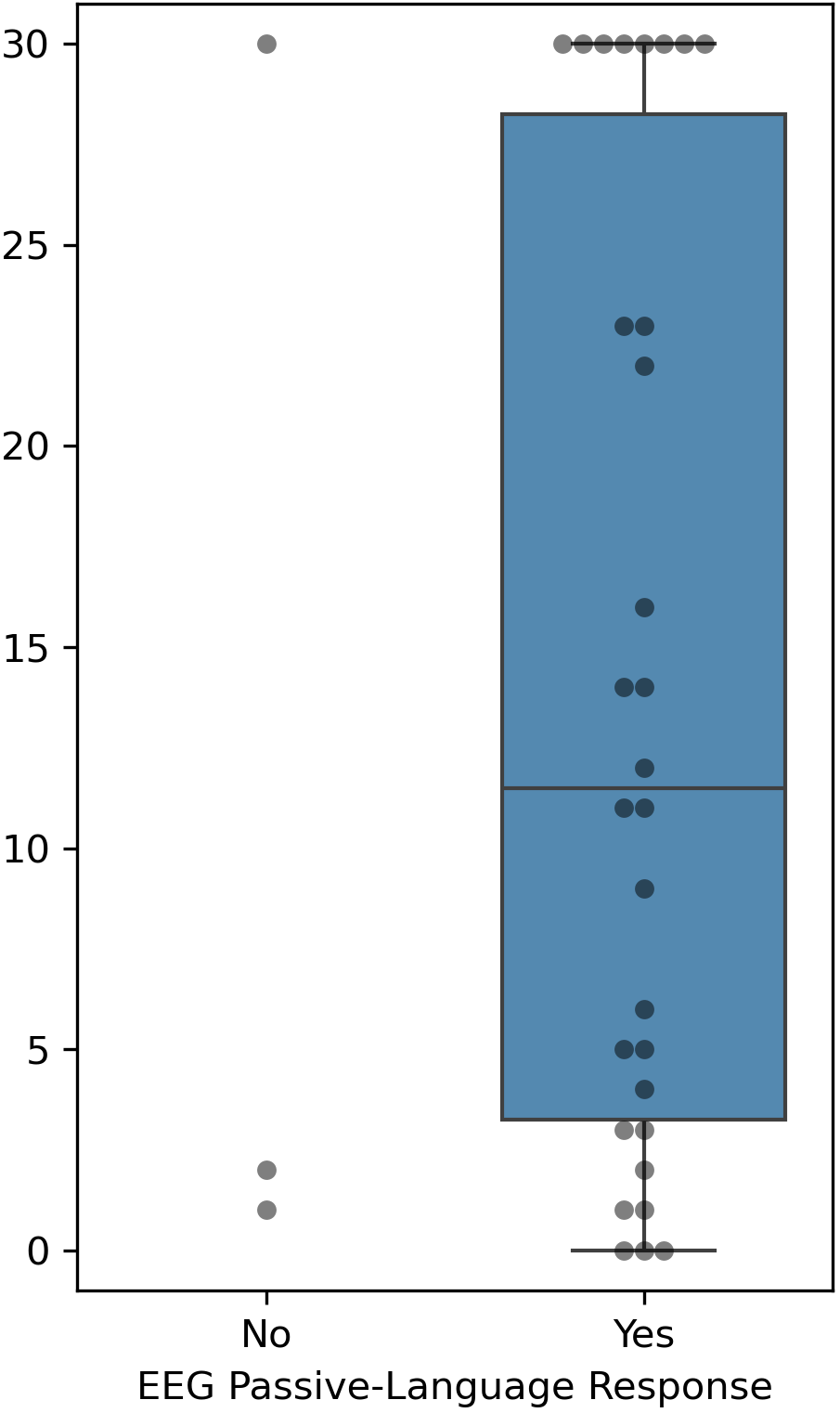

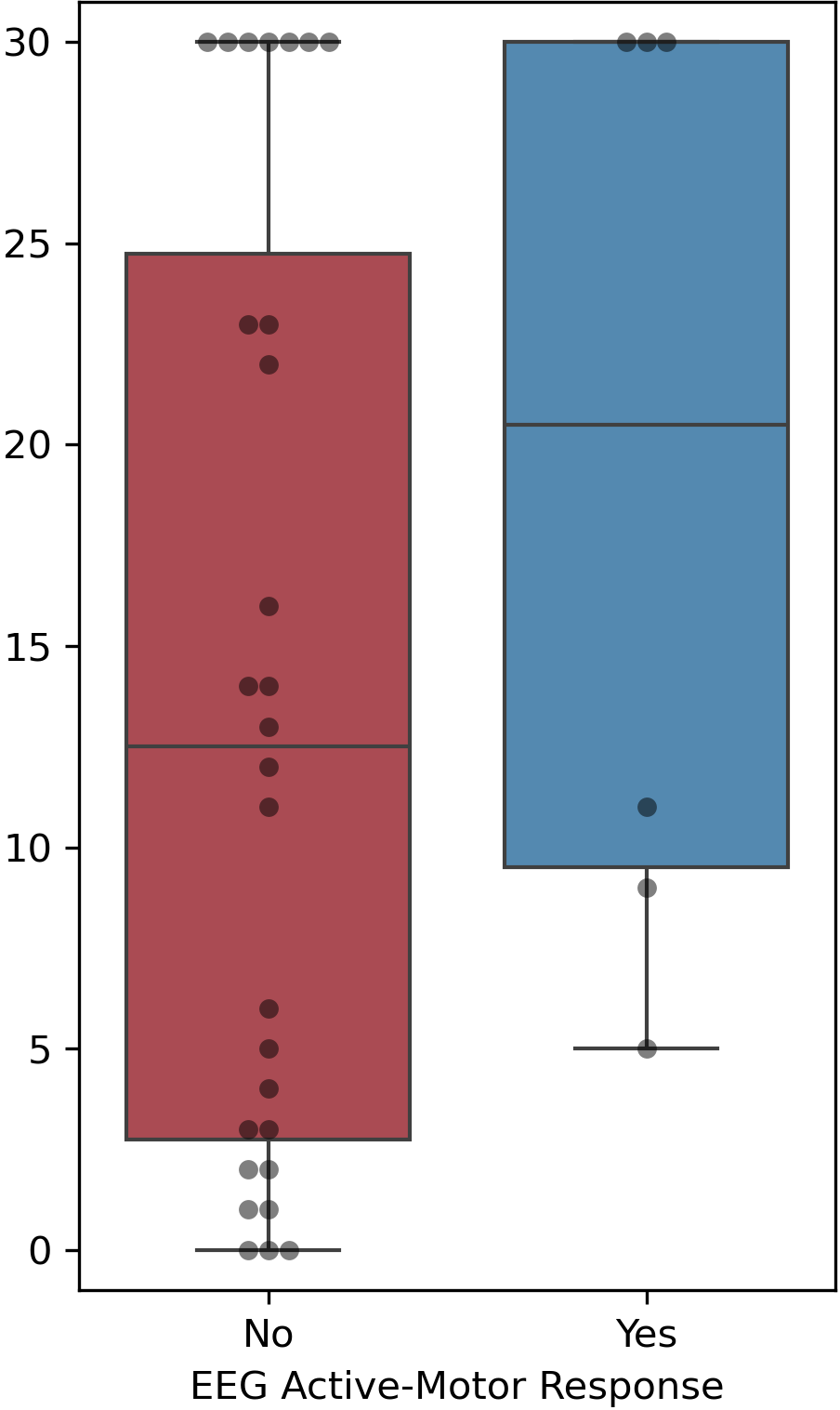

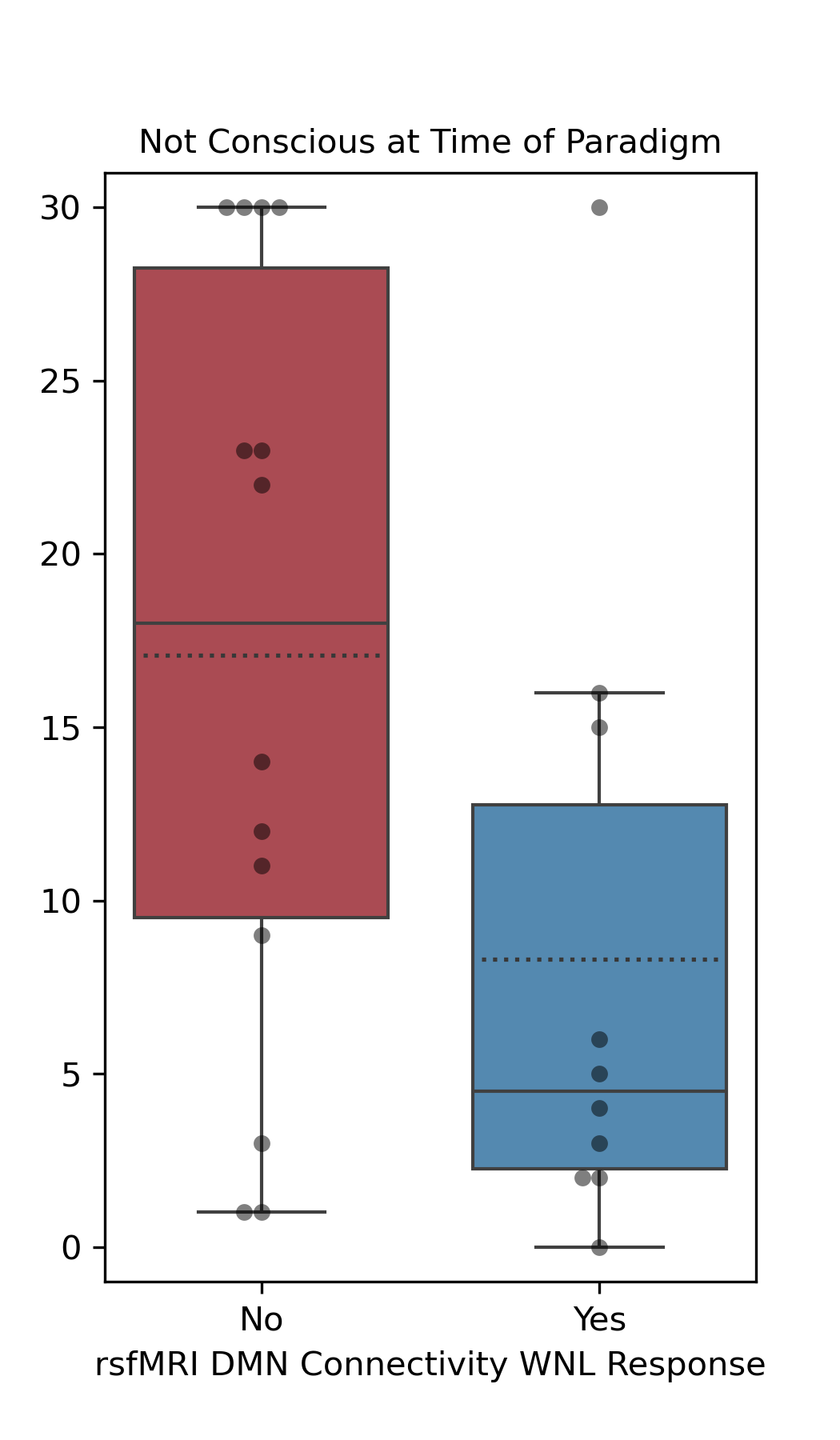

**Supplementary Figure 5**

A

B
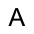

E

C

D
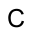

**Supplementary Figure 5: DRS Outcome Across Responses to EEG and fMRI in Participants with a CRS-R Diagnosis Coma, VS/UWS, MCS- Subsample**

In a subsample of participants with an acute CRS-R diagnosis of Coma, VS/UWS, MCS-, the 6-month DRS score is plotted based on the presence of a response to active-motor EEG (A), passive-language EEG (B), active-motor fMRI (C), passive-language fMRI (D), and intact DMN connectivity (E). Box plots are omitted when there are less than 6 data points in a group. NA indicates that the data were not acquired (see Main Manuscript, Table 1). Abbreviations: DoC *disorders of consciousness;* MCS-/MCS+ *minimally conscious state without/with language function*; NA *not acquired;* PTCS *post-traumatic confusional state;* VS/UWS *vegetative state/unresponsive wakefulness syndrome;* WNL *within normal limits (i.e., within 95% confidence interval of healthy control participants).*

**References**

1.	Giacino JT, Kalmar K, Whyte J. The JFK Coma Recovery Scale-Revised: measurement characteristics and diagnostic utility. Arch Phys Med Rehabil 2004;85:2020-9.

2.	Giacino JT, Ashwal S, Childs N, et al. The minimally conscious state: definition and diagnostic criteria. Neurology 2002;58:349-53.

3.	Bruno MA, Majerus S, Boly M, et al. Functional neuroanatomy underlying the clinical subcategorization of minimally conscious state patients. Journal of neurology 2012;259:1087-98.

4.	Schnakers C, Edlow BL, Chatelle C, Giacino J. Minimally conscious state. In: Laureys S, Gosseries O, Tononi G, eds. The Neurology of Consciousness. 2nd ed. San Diego, CA: Academic Press; 2015.

5.	Stuss DT, Binns MA, Carruth FG, et al. The acute period of recovery from traumatic brain injury: posttraumatic amnesia or posttraumatic confusional state? J Neurosurg 1999;90:635-43.

6.	Sherer M, Nakase-Thompson R, Yablon SA, Gontkovsky ST. Multidimensional assessment of acute confusion after traumatic brain injury. Arch Phys Med Rehabil 2005;86:896-904.

7.	Rappaport M, Hall KM, Hopkins K, Belleza T, Cope DN. Disability rating scale for severe head trauma: coma to community. Arch Phys Med Rehabil 1982;63:118-23.

8.	Snider SB, Kowalski RG, Hammond FM, et al. Comparison of common outcome measures for assessing independence in patients diagnosed with disorders of consciousness: a Traumatic Brain Injury Model Systems study. J Neurotrauma 2022;39:1222-30.

9.	Wilson JT, Pettigrew LE, Teasdale GM. Structured interviews for the Glasgow Outcome Scale and the extended Glasgow Outcome Scale: guidelines for their use. J Neurotrauma 1998;15:573-85.

10.	Curley WH, Bodien YG, Zhou DW, et al. Electrophysiological correlates of thalamocortical function in acute severe traumatic brain injury. Cortex 2022;152:136-52.

11.	Delorme A, Makeig S. EEGLAB: an open source toolbox for analysis of single-trial EEG dynamics including independent component analysis. J Neurosci Methods 2004;134:9-21.

12.	Bodien YG, Fecchio, M., Freeman, H. J., Sanders, W. R., Meydan, A., Lawrence, P., Kirsch, J., Fischer D., Cohen J., Rubin E., He J., Schaefer P. W., Hochberg L. R., Rapalino O., Cash S., Young M., Edlow, B. L. Clinical Implementation of Functional MRI and EEG to Detect Cognitive Motor Dissociation: Lessons Learned in an Acute Care Hospital. Neurology: Clinical Practice 2024;In Press.

13.	Edlow BL, Chatelle C, Spencer CA, et al. Early detection of consciousness in patients with acute severe traumatic brain injury. Brain 2017;140:2399-414.

14.	van der Kouwe AJ, Benner T, Salat DH, Fischl B. Brain morphometry with multiecho MPRAGE. Neuroimage 2008;40:559-69.

15.	Smith SM, Jenkinson M, Woolrich MW, et al. Advances in functional and structural MR image analysis and implementation as FSL. Neuroimage 2004;23 Suppl 1:S208-19.

16.	Whitfield-Gabrieli S, Nieto-Castanon A. Conn: a functional connectivity toolbox for correlated and anticorrelated brain networks. Brain Connect 2012;2:125-41.

17.	Kondziella D, Friberg CK, Frokjaer VG, Fabricius M, Møller K. Preserved consciousness in vegetative and minimal conscious states: systematic review and meta-analysis. Journal of Neurology, Neurosurgery & Psychiatry 2016;87:485-92.

18.	Makris N, Goldstein JM, Kennedy D, et al. Decreased volume of left and total anterior insular lobule in schizophrenia. Schizophrenia research 2006;83:155-71.

19.	Eickhoff SB, Stephan KE, Mohlberg H, et al. A new SPM toolbox for combining probabilistic cytoarchitectonic maps and functional imaging data. Neuroimage 2005;25:1325-35.

20.	Monti MM, Vanhaudenhuyse A, Coleman MR, et al. Willful modulation of brain activity in disorders of consciousness. N Engl J Med 2010;362:579-89.

21.	Fernandez-Espejo D, Junque C, Vendrell P, et al. Cerebral response to speech in vegetative and minimally conscious states after traumatic brain injury. Brain Inj 2008;22:882-90.

22.	Bardin JC, Fins JJ, Katz DI, et al. Dissociations between behavioral and functional magnetic resonance imaging-based evaluations of cognitive function after brain injury. Brain 2011;134:769-82.

23.	Coleman MR, Davis MH, Rodd JM, et al. Towards the routine use of brain imaging to aid the clinical diagnosis of disorders of consciousness. Brain 2009;132:2541-52.

24.	Nieto-Castanon A. Handbook of functional connectivity Magnetic Resonance Imaging methods in CONN2020.

25.	Worsley KJ, Marrett S, Neelin P, Vandal AC, Friston KJ, Evans AC. A unified statistical approach for determining significant signals in images of cerebral activation. Hum Brain Mapp 1996;4:58-73.
